# Superior antibody immunogenicity of a RH5 blood-stage malaria vaccine in Tanzanian infants as compared to adults

**DOI:** 10.1101/2023.04.17.23288686

**Authors:** Sarah E. Silk, Wilmina F. Kalinga, Ivanny M. Mtaka, Nasoro S. Lilolime, Maximillian Mpina, Florence Milando, Saumu Ahmed, Ababacar Diouf, Fatuma Mkwepu, Beatus Simon, Thabit Athumani, Mohammed Rashid, Latipha Mohammed, Omary Lweno, Ali M. Ali, Gloria Nyaulingo, Bakari Mwalimu, Sarah Mswata, Tunu G. Mwamlima, Jordan R. Barrett, Lawrence T. Wang, Yrene Themistocleous, Lloyd D. W. King, Susanne H. Hodgson, Ruth O. Payne, Carolyn M. Nielsen, Alison M. Lawrie, Fay L. Nugent, Jee-Sun Cho, Carole A. Long, Kazutoyo Miura, Simon J. Draper, Angela M. Minassian, Ally I. Olotu

## Abstract

**Background:** RH5 is the leading blood-stage candidate antigen for inclusion in a *Plasmodium falciparum* malaria vaccine, however, its safety profile and ability to induce functional immune responses in a malaria-endemic population are unknown. Characterising safety and immunogenicity is key to refine and progress next-generation RH5-based blood-stage malaria vaccines to field efficacy assessment.

**Methods:** A Phase 1b, single-center, dose-escalation, age de-escalation, double-blind, randomized, controlled trial was conducted in Bagamoyo, Tanzania. Healthy adults (18-35 years), young children (1-6 years) and infants (6-11 months) were recruited to receive a priming dose of viral-vectored ChAd63 RH5 (or rabies control vaccine) followed by a booster dose of MVA RH5 (or rabies control vaccine) 8 weeks later. The primary outcomes were the number of solicited and unsolicited adverse events following vaccination and the number of serious adverse events over the whole study period. Secondary outcomes included quantitative and qualitative measures of the anti-RH5 immune response. All participants receiving at least one dose of vaccine were included in the primary analyses.

**Findings:** Between 12^th^ April and 25^th^ October 2018 a total of 63 adults, children and infants were recruited and primed and 60 of these were boosted, all completing six months of follow-up post-priming vaccination. Vaccinations were well-tolerated with participants reporting predominantly mild reactogenicity, with profiles comparable between ChAd63 RH5, MVA RH5 and rabies vaccine groups, and across the age groups. No serious adverse events were reported during the study period. RH5-specific T cell, B cell and serum antibody responses were induced by vaccination. Higher anti-RH5 serum IgG responses were observed post-boost in the 1-6 year old children (median 93 µg/mL; range: 31-508 µg/mL) and infants (median 149 µg/mL; range: 29-352 µg/mL) as compared to adults (median 14 µg/mL; range: 9-15 µg/mL). These contracted over time post-boost, but the same hierarchy of responses across the age groups was maintained to end of follow-up at 16 weeks post-boost (day 168). Vaccine-induced anti-RH5 antibodies were functional showing growth inhibition activity (GIA) *in vitro* against *P. falciparum* blood-stage parasites. The highest levels were observed in the 6-11 month old infants, with 6/11 showing >60% GIA following dilution of total IgG to 2.5 mg/mL (median 61%; range: 41-78%).

**Interpretation:** The ChAd63-MVA RH5 vaccine regimen shows an acceptable safety and reactogenicity profile and encouraging immunogenicity in children and infants residing in a malaria-endemic area. The levels of functional GIA observed in the RH5 vaccinated 6-11 month old infants are the highest levels reported to-date following human vaccination. These data support onward clinical development of RH5-based blood-stage vaccines that aim to protect against clinical malaria in young African infants.

**Funding:** Medical Research Council, London, United Kingdom.

**Trial Registration:** ISRCTN registry: 47448832 and ClinicalTrials.gov: NCT03435874.

## Introduction

Over the last two decades, expanded access to malaria prevention tools has led to a major reduction in the global burden of this disease, however, progress has stalled with current data showing that cases and deaths are now rising once again, likely magnified by the SARS-CoV-2 pandemic ^1^. Notably, over 80% of deaths occur in children under the age of 5 in sub-Saharan Africa and a highly effective vaccine remains urgently needed. Encouragingly, subunit vaccine strategies targeting the invasive sporozoite stage of *Plasmodium falciparum* are now showing moderate levels of efficacy in field trials ^2, 3^, however, durability of protection remains a key challenge. These vaccines also necessitate sterilizing immunity, with only a single break-through sporozoite leading to the subsequent pathogenic blood-stage of infection. An alternative and complementary approach is to vaccinate against the blood-stage merozoite to inhibit erythrocyte invasion, thus, leading to control and/or clearance of blood-stage parasitemia, minimizing morbidity and mortality and reducing transmission ^4^. Indeed, combining a blood-stage vaccine component with existing anti-sporozoite vaccines is now widely regarded as the most promising strategy to achieve a high efficacy intervention.

While a number of factors have long-stalled progress in the design of vaccines that can impact the blood-stage of the parasite’s lifecycle ^4^, the identification of an essential, highly conserved, antibody-susceptible, protein complex used by *P. falciparum* merozoites to invade erythrocytes has propelled the field over the last 10 years ^5^. Vaccine development efforts are most advanced for one component of this invasion complex: reticulocyte-binding protein homolog 5 (<u>RH5</u>) ^6^. This vaccine target binds <u>basigin</u> on the <u>erythrocyte surface</u> ^7^, a receptor-ligand interaction that is critical for parasite invasion and its human host tropism ^8^. Significant *in vivo* protection with RH5-based vaccines was first demonstrated against a stringent blood-stage *P. falciparum* challenge in an *Aotus* monkey model ^9^. Here, protection was correlated with functional antibody activity measured using an *in vitro* assay of growth inhibition activity (GIA); an immune mechanism subsequently validated by passive transfer of anti-RH5 monoclonal antibody ^10^.

This paved the way for the first RH5-based vaccine to enter Phase 1a clinical testing in healthy UK adults in 2014 (VAC057; ClinicalTrials.gov: NCT02181088). This vaccine utilized recombinant replication-deficient chimpanzee adenovirus serotype 63 (ChAd63) and the attenuated orthopoxvirus modified vaccinia virus Ankara (MVA), delivered in an 8-week prime-boost regimen, to enable *in situ* expression of RH5 by virally infected cells (ChAd63-MVA RH5). The vaccine was well-tolerated and induced functional human antibodies that exhibited cross-strain *in vitro* GIA ^11^. However, although the levels of anti-RH5 serum <u>immunoglobulin G</u> (IgG) in the UK adult participants greatly exceeded the levels observed in African adults following years of natural malaria exposure, they fell below the protective immunological threshold predicted by the *Aotus* model. In parallel and in a series of other clinical trials, immunization with the same ChAd63-prime MVA-boost viral-vectored delivery platform recombinant for the liver-stage malaria antigen ME-TRAP, reported ten-fold higher antibody levels in West African infants as compared to UK adults and West African adults ^12^. Given deployment trials of the WHO pre-qualifed RTS,S/AS01 vaccine are focussed on infants starting from 5 months of age, and given that a ten-fold higher anti-RH5 IgG antibody response in African infants could potentially translate into protection against clinical malaria, we proceeded to further assess the ChAd63-MVA RH5 in this target population.

We therefore conducted a single-center, dose-escalation, age de-escalation, double-blind, randomized, controlled Phase 1b trial to explore the tolerability, safety and immunogenicity of an RH5-based vaccine in a malaria-endemic area for the first time, to inform crucial decisions on progress to field efficacy studies and/or iterative vaccine refinement.

## Results

### Study recruitment and vaccinations

In total 125 participants were screened and 63 of these were enrolled into the VAC070 Phase 1b trial (**Figure 1**). Vaccinations began on 12^th^ April 2018 and all follow-up visits were completed by 13^th^ February 2019. Within each group, participants were randomized to receive the ChAd63-MVA RH5 vaccine or rabies control vaccine in a ratio of 2:1. Participants and all investigatory staff (involved in evaluation of safety and immunogenicity endpoints) were blinded to vaccine allocation. All participants received their immunizations as scheduled and completed 6 months of follow-up post priming vaccination, apart from three individuals who withdrew from the study after the prime. Similar numbers of males and females were enrolled across the younger age groups (**Table S1**). Independent safety reviews were conducted by the Safety Monitoring Committee (SMC) between every age de-escalation and/or dose-escalation step in the protocol.

**Figure 1.**
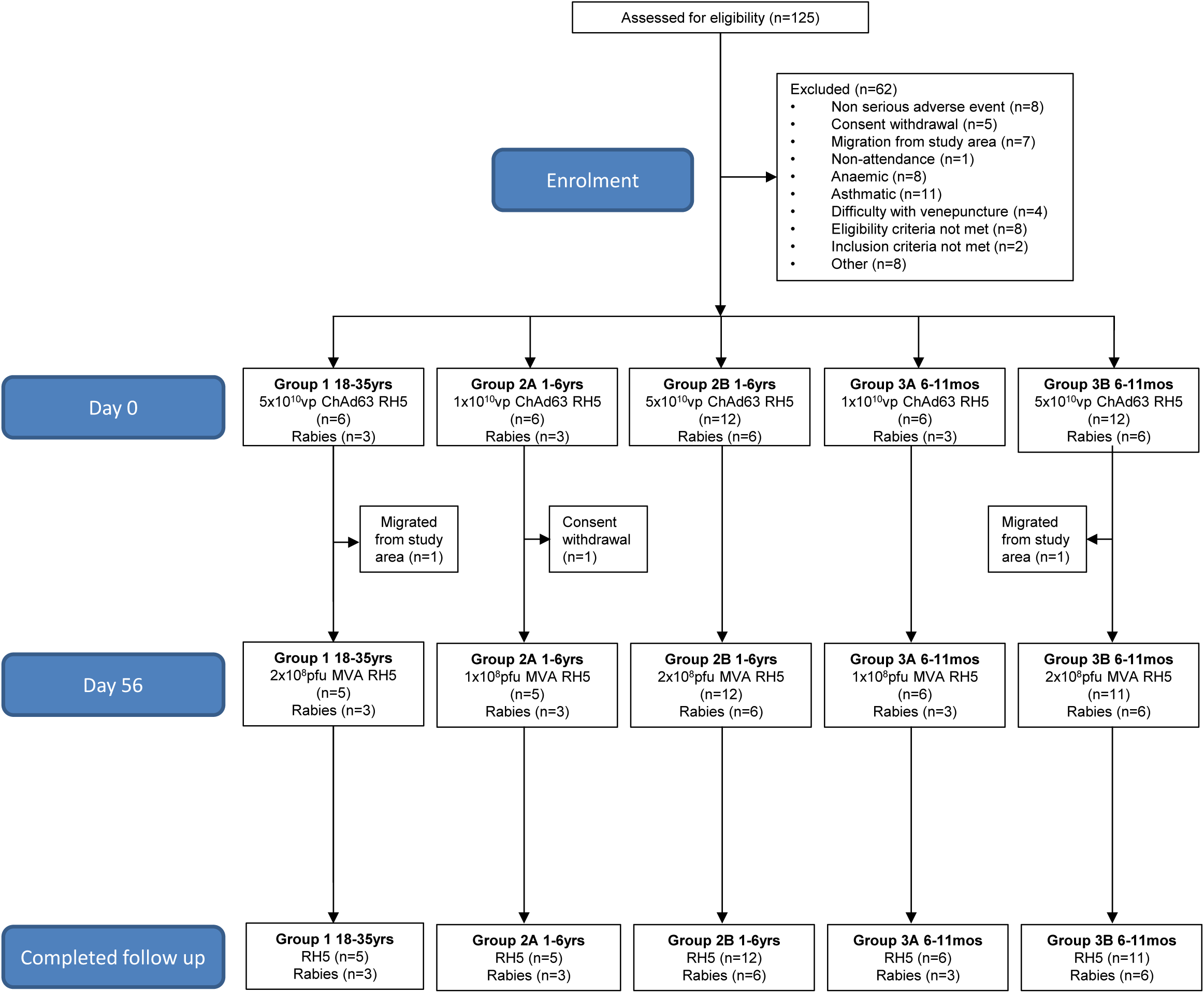
VAC070 flow chart of study design and participant recruitment. Screening into the VAC070 study began in March 2018 and all follow-up visits were completed by February 2019. All immunizations were administered IM into the left deltoid area.

All participants were recruited from the district of Bagamoyo, Tanzania. Malaria prevalence in the district ranges from 15.4% in the western part to at times as low as zero in Bagamoyo town; the population average prevalence was 13% in 2013 ^13^. The mean age of adult participants in Group 1 was 25.8 years (range 19.0 – 30.7 years), and they received the full dose of both vaccines via the intramuscular (IM) route: 5 × 10^10^ viral particles (vp) ChAd63 RH5 to prime on day 0 and 2 × 10^8^ plaque-forming units (pfu) MVA RH5 to boost on day 56. The mean age of children across Groups 2A and 2B was 3.6 years (range 1.2 – 5.6 years), and infants across Groups 3A and 3B was 9.4 months (range 5.9 – 11.9 months). Those in Groups 2A and 3A received lower “lead-in” doses of vaccine: 1 × 10^10^ vp ChAd63 RH5 and 1 × 10^8^ pfu MVA RH5, and those in Groups 2B and 3B received the full vaccine dose (all via the IM route).

A total of sixty-three adults, children and infants were recruited and primed and sixty of these were boosted (three withdrew post-prime). Thirty-nine participants were primed with ChAd63 RH5 and boosted with MVA RH5, twenty-one participants received two doses of rabies vaccine 8 weeks apart. All boosted participants completed six months of follow-up post-prime and analysis was by original assigned groups. All participants (RH5 and rabies vaccinated) were negative for malaria by blood film throughout the trial (tested at screening and monthly intervals post-vaccination). We also conducted retrospective analysis for malaria parasitemia by highly sensitive qPCR on blood samples taken from every participant at the day 0, day 63 and day 84 time-points. All samples tested negative, except for only one transient low level asymptomatic parasitemia of 450 parasites/mL blood in a single RH5 vaccinated Group 2B child at the day 63 time-point (7 days post-MVA RH5 boost).

## Outcomes and estimation

### Reactogenicity and safety

There were no serious adverse events (SAEs), AEs of special interest (AESIs) or unexpected reactions and no safety concerns during the course of the trial. The local and systemic reactogenicity profile of the full dose RH5 vaccines (**Figure 2**) was similar, if not reduced, as compared to that seen in healthy UK adults immunized with the identical vaccines ^11^. Following the prime with ChAd63 RH5, the majority of participants across all age groups experienced local pain at the injection site and systemic fever (>37.5°C) (**Figure 2A–C**); all of these adverse events (AEs) were mild in severity, with similar profiles seen in those that received the rabies control vaccine (**Figure S1A-C**). A very similar pattern was observed following the boost with MVA RH5 (**Figure 2D–F**), with some of the older participants also experiencing Grade 1 (mild) erythema, in contrast to the boost with the rabies vaccine where hardly any local or systemic AEs were observed (**Figure S1D-F**). Very similar AE profiles were observed in the Group 2A children and Group 3A infants who received the lower “lead-in” doses of the ChAd63-MVA RH5 vaccine or the rabies control vaccine (**Figure S2**). The majority of solicited AEs occurred within the first two days after vaccination and the median duration of each local or systemic AE was between 1 and 2 days following either vaccine. Of the unsolicited AEs recorded within 28 days of vaccination, none were considered related to any of the vaccines (**Table S2**). None were recorded in the adult group or following the ChAd63 RH5 prime. Those recorded following MVA RH5 or rabies vaccination were mostly mild in nature, with only a few graded moderate and none graded severe, and all resolved spontaneously. There were no Grade 3 unsolicited AEs. Viral upper respiratory tract infection (N=15) was the most common unsolicited AE among the infants 6-11 months of age with similar numbers observed in the MVA RH5 vaccine and rabies vaccine groups.

**Figure 2.**
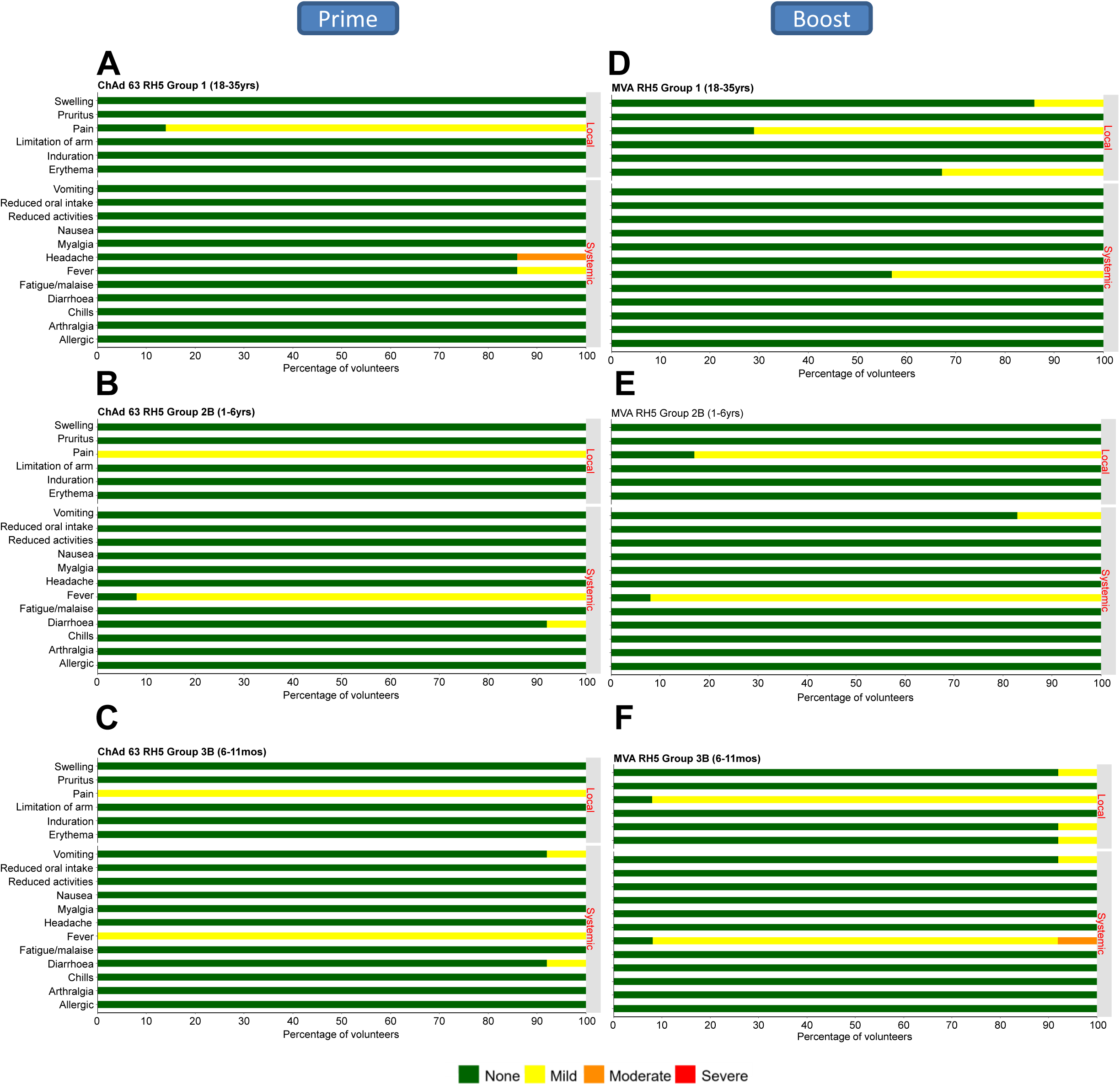
Solicited AEs following vaccination with full dose ChAd63 and MVA RH5. The solicited local and systemic AEs recorded for 7 days following ChAd63 RH5 prime and MVA RH5 boost are shown at the maximum severity reported by all participants within the VAC070 trial Groups 1, 2B and 3B. Participants aged (**A**) 18-35 years (n=6); (**B**) 1-6 years (n=12); and (**C**) 6-11 months (n=12) received the full priming dose of 5 × 10^10^ vp ChAd63 RH5 on day 0. The same participants aged (**D**) 18-35 years (n=5); (**E**) 1-6 years (n=12); and (**F**) 6-11 months (n=11) received the full boosting dose of 2 × 10^8^ pfu MVA RH5 on day 56. Rabies vaccine control data are shown in **Figure S1** and data for the lower lead-in dose of ChAd63-MVA RH5 (trial Groups 2A and 3A) are shown in **Figure S2**.

The most frequent laboratory abnormality identified in the 28-day post vaccination period was increased leukocytes, present in 9/42 vaccinees and 3/18 controls (*P*=0.74, 2-tailed Fisher’s exact test). These included four Grade 3 occurrences in the 6-11 month old infants (all in the ChAd63-MVA RH5 group). These elevations were associated with concurrent infections and resolved with treatment of the infection (**Table S3**). Other commonly observed laboratory abnormalities included mild and moderate anemia, mild increases in alanine aminotransferase (ALT) levels, mild decreases in leukocytes, and mild decreases in lymphocytes. The frequencies of these abnormalities did not differ between the vaccinees and controls (*P*=0.74, *P*=0.65, *P*=0.66 and *P*>0.99, respectively; 2-tailed Fisher’s exact test). Full blood count analyses at the end of trial follow-up were all normal.

## Immunogenicity

### IFN-γ T cell responses induced by ChAd63 and MVA RH5 are higher in adult than in children and infant participants

The kinetics and magnitude of the RH5-specific T cell response were assessed over time by *ex-vivo* IFN-γ ELISPOT following re-stimulation of PBMC with 20mer peptides overlapping by 10 amino acids (aa) spanning the entire RH5 insert present in the vaccines (**Figure 3A,B, S3A-D, Table S4**). Following ChAd63 RH5 prime, comparable responses were detectable in the Tanzanian adults and children on day 14 (median 271 [range 7 – 2723] and median 243 [range 60 – 1147] spot forming units (SFU) / million PBMC, respectively). These were approximately 3-fold lower than the responses previously observed in UK adults who received the identical dose of vaccine ^11^, but significantly higher than those seen in the Tanzanian infants (median 19 [range 0 – 745] SFU / million PBMC) who also mirrored the rabies vaccine control recipients (**Figure 3A**). Subsequently, administration of MVA RH5 boosted these responses in the Tanzanian adults, as measured four weeks later on day 84 with a median of 1159 [range 740 – 1935] SFU / million PBMC. These responses were now comparable to those previously seen post-boost in the UK adults ^11^, but significantly higher than those in the Tanzanian children (where no appreciable boost in the T cell response was observed) and the Tanzanian infants where a modest boost reached a median level of 125 [range 0 – 793] SFU / million PBMC (**Figure 3B**). As expected for T cell responses, these then contracted over time, but the same trend as day 84 was also observed 16 weeks post-boost at the end of study period (day 168), with responses in all three age groups maintained above baseline and above those observed in the rabies vaccine control recipients (**Figure S3D**). However, as expected, we also observed that peripheral lymphocyte counts decreased with age as measured by routine hematology tests at each time point (**Figure 3C**). Therefore, as reported in a previous pediatric study of a viral vectored malaria vaccine ^12^, we also analyzed the data by incorporating the lymphocyte count into the calculation of T cell responses, to facilitate a more physiologically relevant comparison of cellular immunity across the different age groups. Here, only the 6-11 month old infants still showed a significantly lower response following the ChAd63 RH5 prime (**Figure S3E**), whilst there was no significant difference across the age groups after the MVA RH5 boost (**Figure 3D**) and the end of follow up (**Figure S3F**). Finally, we also assessed the responses at day 14 and day 84 in the Tanzanian children and infants receiving the lower “lead in” doses of ChAd63 and MVA RH5. Here, these T cell responses were also comparable to those induced by the full dose of each vaccine (**Figure S3G,H**).

**Figure 3.**
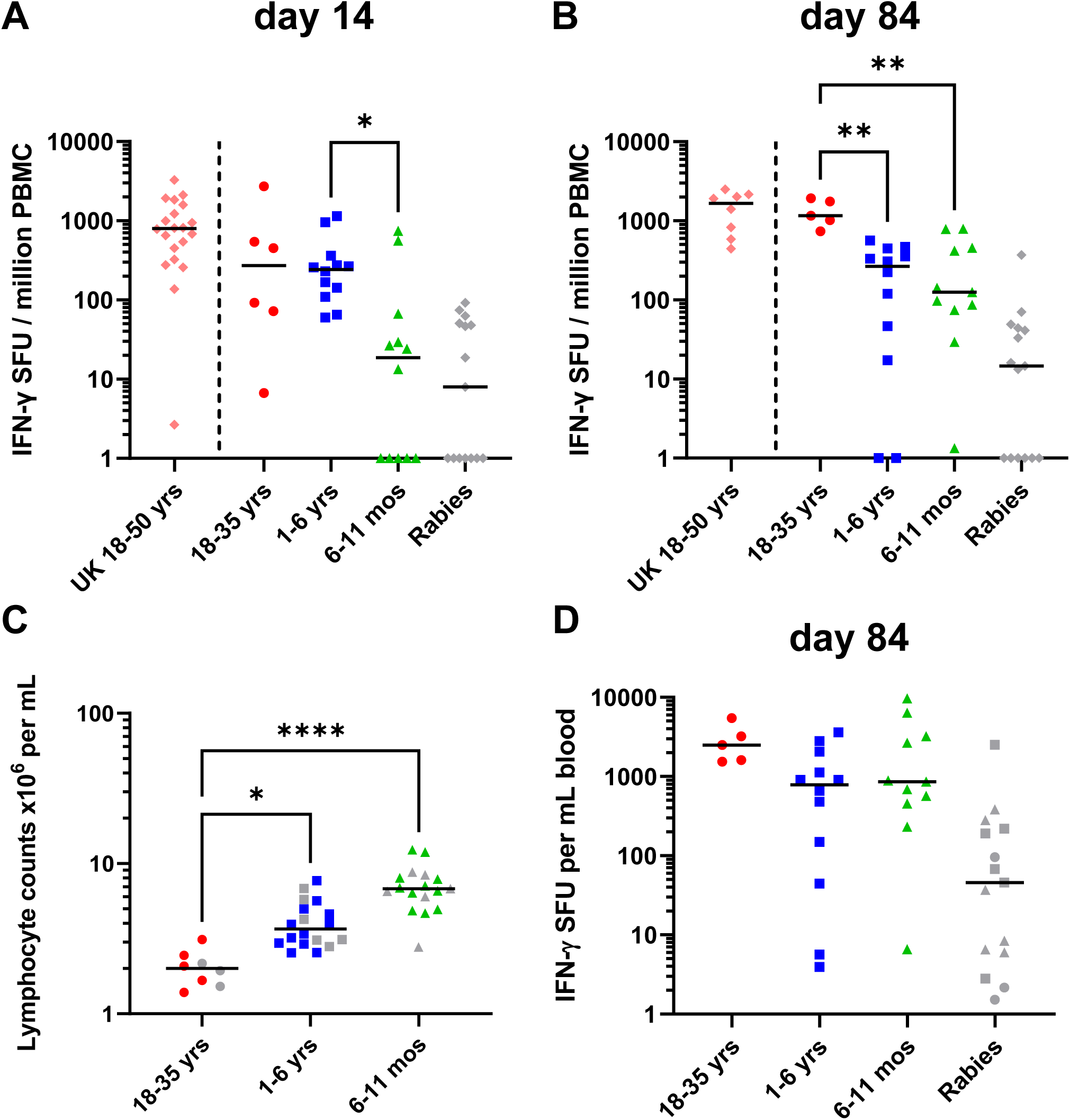
*Ex-vivo* IFN-γ T cell response to ChAd63-MVA RH5 vaccination. Median and individual *ex-vivo* IFN-γ enzyme-linked immunospot (ELISPOT) responses to RH5 per million peripheral blood mononuclear cells (PBMC) are shown for all groups receiving the full dose of ChAd63-MVA RH5 vaccines and rabies vaccine control. Responses are shown at (**A**) day 14, two weeks post-ChAd63 RH5 prime; and (**B**) day 84, four weeks post-MVA RH5 boost. Group 1 adults 18-35 years, N=5 or 6; Group 2B children 1-6 years, N=12; Group 3B infants 6-11 months, N=11 or 12; and rabies vaccine control, all groups and ages pooled, N=15. Historical data testing the identical dose and regimen of the ChAd63-MVA RH5 vaccine in healthy UK adults 18-50 years are shown for comparison only: day 14, N=20; and day 84, N=8 ^11^. (**C**) Lymphocyte counts per mL of blood for all vaccinated participants at day 84. Group 1 adults 18-35 years, N=8; Group 2B children 1-6 years, N=18; Group 3B infants 6-11 months, N=17. (**D**) Median and individual *ex-vivo* IFN-γ ELISPOT responses to RH5 per mL peripheral blood are shown for all groups receiving the full dose of ChAd63-MVA RH5 vaccines and rabies vaccine control at day 84. Groups as per panel (**B**). Analyses using Kruskal-Wallis test with Dunn’s multiple comparison test across Groups 1, 2B and 3B only; \**P* < 0.05, \*\**P* < 0.01, and \*\*\*\**P*<0.0001.

### Anti-RH5 serum IgG and B cell responses induced by ChAd63 and MVA RH5 are higher in children and infants than in adults

We next measured the kinetics and magnitude of the anti-RH5 serum IgG antibody response over time by ELISA against full-length RH5 (RH5_FL) recombinant protein (**Figure 4A,B, S4A-D**). Following ChAd63 RH5 prime, the magnitude of the antibody response showed a clear age-dependent hierarchy.

**Figure 4.**
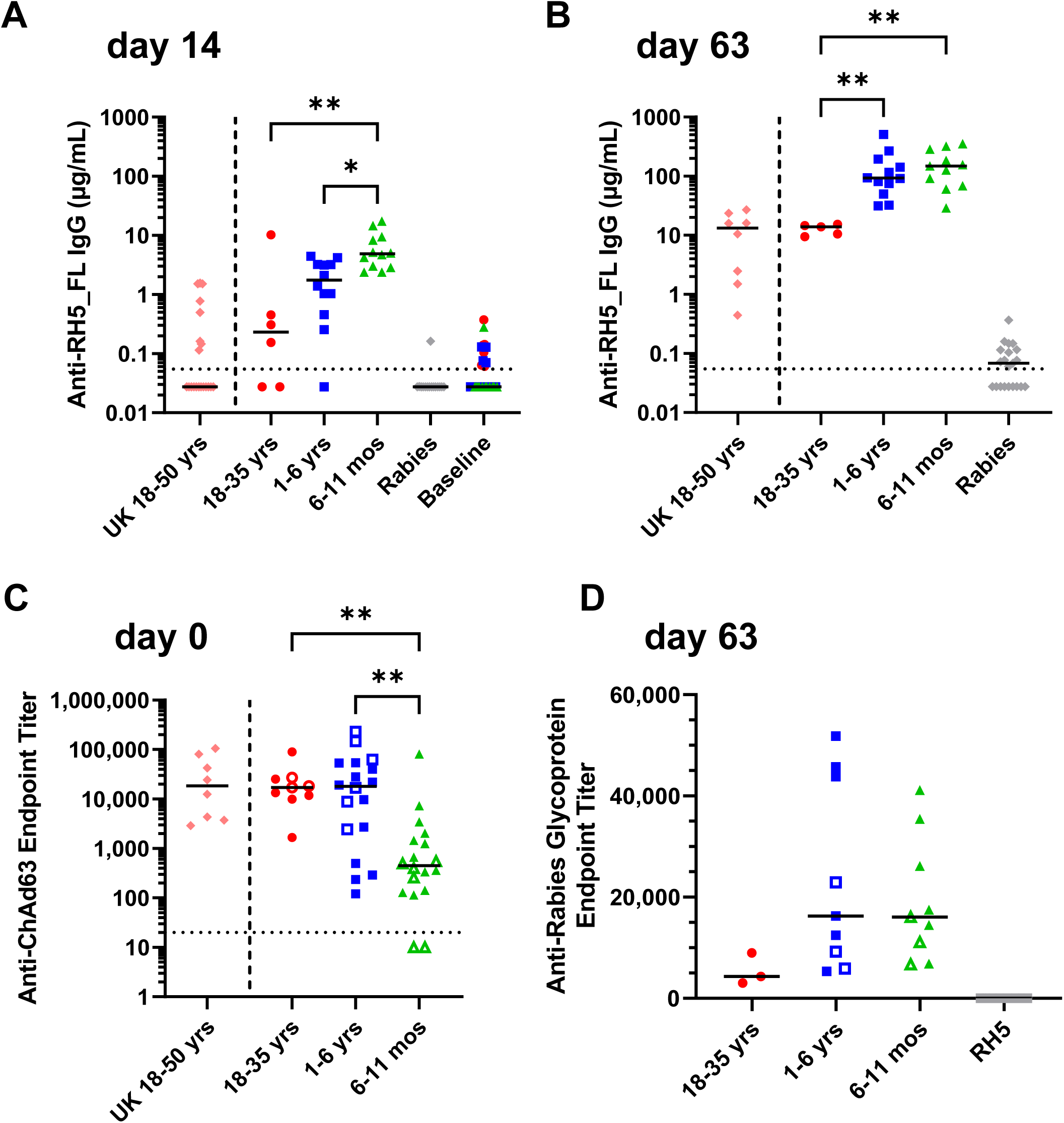
Serum antibody response to vaccination. Median and individual anti-RH5_FL serum total IgG responses as measured by ELISA are shown for all groups receiving the full dose of ChAd63-MVA RH5 vaccines and rabies vaccine control. Responses are shown at (**A**) day 14, two weeks post-ChAd63 RH5 prime; and (**B**) day 63, one week post-MVA RH5 boost. Group 1 adults 18-35 years, N=5 or 6; Group 2B children 1-6 years, N=12; Group 3B infants 6-11 months, N=11 or 12; and rabies vaccine control, all groups and ages pooled, N=15. Baseline = day 0 pre-vaccination time-point shown for Groups 1, 2B and 3B combined and color-coded. Historical data testing the identical dose and regimen of the ChAd63-MVA RH5 vaccine in healthy UK adults 18-50 years are shown for comparison only: day 14, N=20; and day 63, N=8 ^11^. The horizontal dotted line indicates the limit of detection of the assay. Responses to the “lead in” vaccine doses are shown in **Figure S4**. (**C**) Median and individual anti-ChAd63 serum IgG endpoint titers as measured by ELISA in pre-vaccination (day 0) samples. Solid symbols = participants who subsequently underwent ChAd63-MVA RH5 vaccination and open symbols = those who received rabies vaccine control. UK adults 18-50 years, N=8 ^11^; Group 1 adults 18-35 years, N=9; Group 2B children 1-6 years, N=18; Group 3B infants 6-11 months, N=18. (**D**) Median and individual anti-rabies glycoprotein serum total IgG responses as measured by ELISA in day 63 samples (after two vaccine doses) are shown for all individuals receiving rabies vaccine control: Group 1 adults 18-35 years, N=3; Group 2 children 1-6 years, N=9; Group 3 infants 6-11 months, N=9; open symbols = Groups 2A and 3A, closed symbols = Groups 2B and 3B; as well as the ChAd63-MVA RH5 vaccinees (N=28, Groups 1, 2B and 3B combined). Analyses shown used Kruskal-Wallis test with Dunn’s multiple comparison test across Groups 1, 2B and 3B only; \**P* < 0.05, \*\**P* < 0.01.

Here, on day 14, the lowest responses were detected in the Tanzanian adults, with higher responses observed in the children 1-6 years of age, and the highest in the infants 6-11 months of age; median levels of serum anti-RH5 IgG were 0.2, 1.8 and 4.9 µg/mL, respectively (in contrast to negligible median responses in all groups at baseline). Median responses in UK adults who previously received the identical dose of the ChAd63 RH5 vaccine ^11^ or in the rabies vaccine control recipients were also negligible at this time-point (**Figure 4A**). Subsequently, administration of MVA RH5 boosted these responses in all three age groups, but the same hierarchy was maintained as measured by ELISA one week later on day 63 (**Figure 4B**). Median responses of serum anti-RH5 IgG in the Tanzanian adults were 14 µg/mL, now highly comparable to those previously observed in UK adults who received the identical dose of the viral vaccines ^11^; whilst significantly higher responses were observed in the children (∼6-fold, median of 93 µg/mL) and infants (∼10-fold, median of 149 µg/mL). Antibody responses induced by the vaccines were also dose dependent; here, responses at day 14 and day 63 were significantly lower in the Tanzanian infants receiving the lower “lead in” doses of ChAd63 and MVA RH5 as compared to those receiving the full dose of each vaccine, with a similar trend observed in the children (**Figure S4E,F**). As expected for serum IgG responses, these contracted over time post-boost, but the same hierarchy of responses across the age groups was maintained out to 16 weeks post-boost at the end of study period (day 168) (**Figure S4A**).

Having observed such high anti-RH5 serum IgG responses in children and infants, we hypothesized this could be due to anti-vector immunity that may increase with age and thereby negatively affect the priming immunogenicity of adenovirus-vectored vaccines in adults. However, although the 6-11 month old Tanzanian infants did show the lowest anti-ChAd63 antibody responses at baseline (consistent with previous data on antibody responses to human adenovirus serotypes in this age group ^14^), the children aged 1-6 years and adults in our study were comparable (**Figure 4C**). Consequently, there was no significant correlation between existing anti-ChAd63 antibody responses at baseline and the day14 ChAd63 RH5 humoral immunogenicity (Spearman’s r_s_ = –0.35, *P* > 0.05, N=30). In line with this, we also measured the anti-rabies glycoprotein serum antibody responses in the control participants. Here we observed a similar age-dependent hierarchy, despite the small number of vaccinees, with responses in the children and infants on average ∼4-5-fold higher than in adults after the first and second immunizations (**Figure 4D, S4G,H**).

We therefore next measured the underlying B cell response. Previous studies have shown that antibody-secreting cells (ASC) can be detected in peripheral blood for a short time (around day 7) after MVA boost when using the ChAd63-MVA regimen ^11, 15, 16^. RH5-specific ASC responses were assessed by *ex-vivo* ELISPOT using fresh PBMC collected at the day 63 visit for participants. ASC responses were detectable above baseline in all age groups vaccinated with ChAd63-MVA RH5, with the highest responses trending to be measured in the Group 3B infants with a median of 60 RH5-specific ASC per million PBMC (**Figure 5A**). We subsequently assessed RH5-specific B cell responses further by flow cytometry (**Figure 5B, S5A**). As noted during the analysis of the T cell responses, clinical hematology data showed a clear age-dependent hierarchy in the lymphocyte counts per microliter of blood (**Figure S5B**).Within these total lymphocyte populations, flow cytometry analysis showed the % live B cells to be approximately 2.5-fold higher in the pediatric groups as compared to adults (**Figure S5C**). Combining these data showed that 6-11 month old infants have significantly more B cells per microliter of blood (approximately 5-fold on average) as compared to the adults 18-35 years of age, with children 1-6 years of age having intermediate levels (**Figure 5C**). Following the MVA-RH5 vaccine boost, children and infants showed comparable RH5-specific responses within the live CD19^+^ IgG^+^ B cell population. These frequencies were significantly higher in the infants (approximately 10-fold on average) as compared to the vaccinated adults (**Figure 5D**).

**Figure 5.**
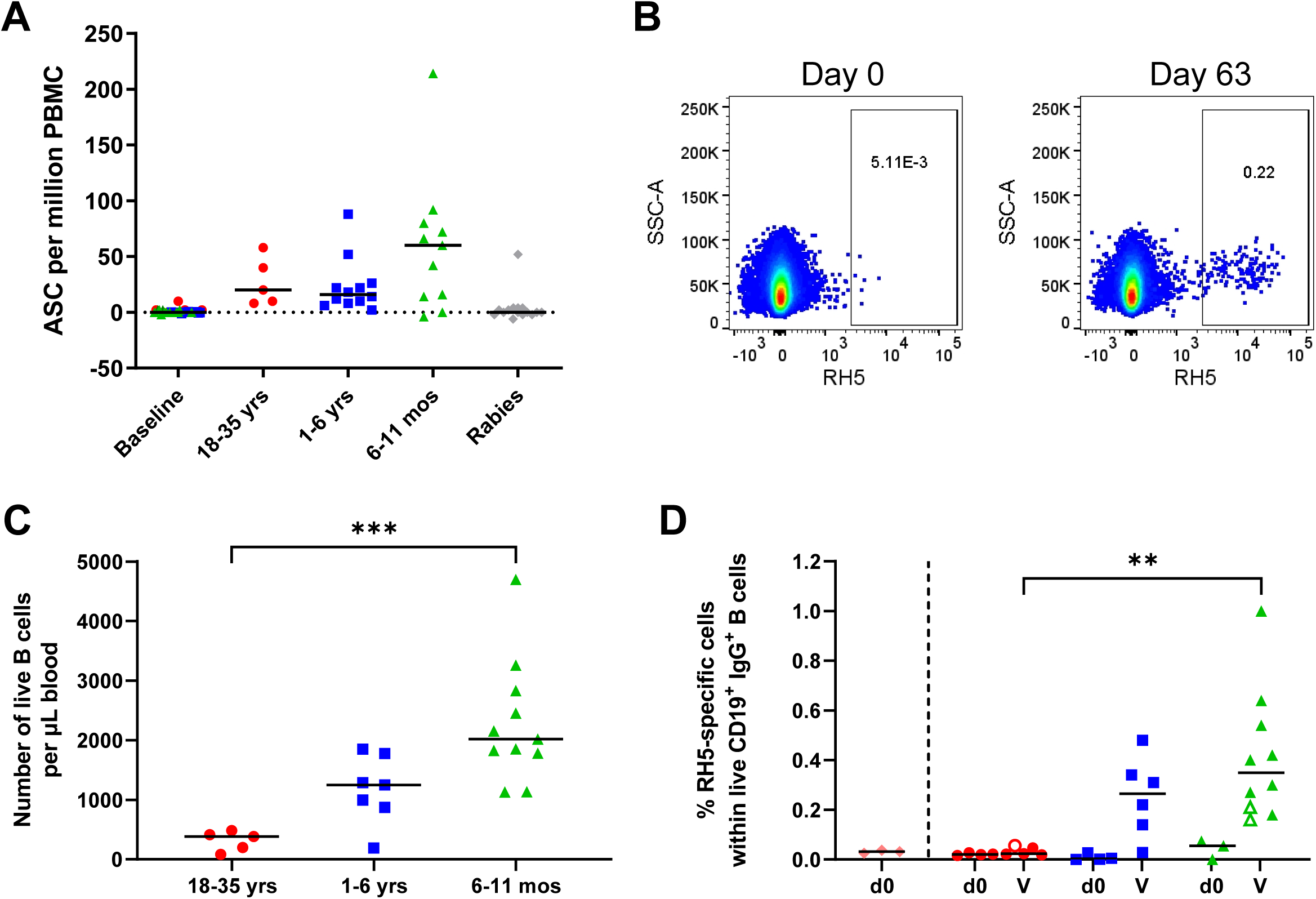
B cell response to ChAd63-RH5 vaccination. (**A**) RH5-specific antibody-secreting cell (ASC) responses were assessed by *ex-vivo* enzyme-linked immunospot (ELISPOT) using RH5_FL protein and fresh peripheral blood mononuclear cells (PBMC) from the day 63 time-point. Responses are reported as RH5-specific ASC per million PBMC used in the assay. Group 1 adults 18-35 years, N=5; Group 2B children 1-6 years, N=12; Group 3B infants 6-11 months, N=11; Baseline = Groups 1, 2B and 3B combined, N=30; Rabies = Groups 1, 2B and 3B combined, N=15. (**B**) Representative flow cytometry plots showing definition of RH5-specific cells within the live CD19^+^ IgD^−^ IgM^−^ IgA^−^ B cell population (see **Figure S5A** for full gating strategy). Representative plots are shown for matched day 0 (pre-vaccination) and day 63 (one week post-MVA RH5 boost vaccination) samples from a single vaccinee. (**C**) Number of live CD19^+^ B cells per microliter of blood, as calculated by combining data from lymphocyte counts per microliter of blood (**Figure S5B**) and percentage of live CD19^+^ B cells within lymphocytes as assessed by flow cytometry (**Figure S5C**). (**D**) Percentage of RH5-specific cells within the CD19^+^ IgD^−^ IgM^−^ IgA^−^ B cell population as assessed by flow cytometry. Same individuals and color coding for age groups as shown in (**C**) measured at baseline day 0 (d0) or at 7 days post-MVA RH5 boost vaccination (V, closed symbols); N=3 individuals were measured at 28 days post-MVA RH5 boost due to sample availability (open symbols). Data from baseline samples for N=3 healthy UK adult participants 18-50 years of age enrolled in a previous trial of the ChAd63-MVA RH5 vaccine ^11^ are shown for comparison only (pink diamonds to left of dashed line). Individual and median results are shown in each panel. Analyses used Kruskal-Wallis test with Dunn’s multiple comparison test across the three groups in (**C**) or the three vaccinated groups in (**D**); \*\**P* < 0.01, *** *P* < 0.001.

### Vaccine-induced antibodies show high-level functional GIA *in vitro*

Finally, serum samples were analyzed for functional anti-parasitic growth inhibition at the GIA Reference Center at the NIH. Here, the standardized GIA assay of human samples typically tests purified total IgG (normalized to a starting concentration of 10 mg/mL) against *P. falciparum* 3D7 clone parasites in the absence of complement ^17, 18^. However, we initially measured each serum sample’s physiological total serum IgG concentration by HPLC (**Figure S6A**). These data showed the median level in 6-11 month old infants to be 10.1 mg/mL, highly comparable to that previously seen in healthy UK adults ^17^. In contrast, the levels in 1-6 year old children (median 13.7 mg/mL) and 18-35 year old adults (median 14.8 mg/mL) were significantly higher. We therefore elected instead to initially screen all samples for GIA starting at their physiological total IgG concentration as opposed to normalizing to 10 mg/mL. At these higher starting levels of total IgG, some weak GIA was observed at baseline, especially in the adults (**Figure 6A**), in line with higher levels of prior malaria exposure in this age group as confirmed by an anti-parasite lysate ELISA (**Figure 6B**). However, following ChAd63-MVA RH5 vaccination, very large increases in GIA were observed in the Group 2B children and Group 3B infants (**Figure S6B**), reaching median levels of 89.2% and 98.5%, respectively, at their physiological IgG concentration at the day 63 time-point (**Figure 6A**). These increases were not observed in the RH5 vaccinated adults or any of the rabies vaccine control groups.

**Figure 6.**
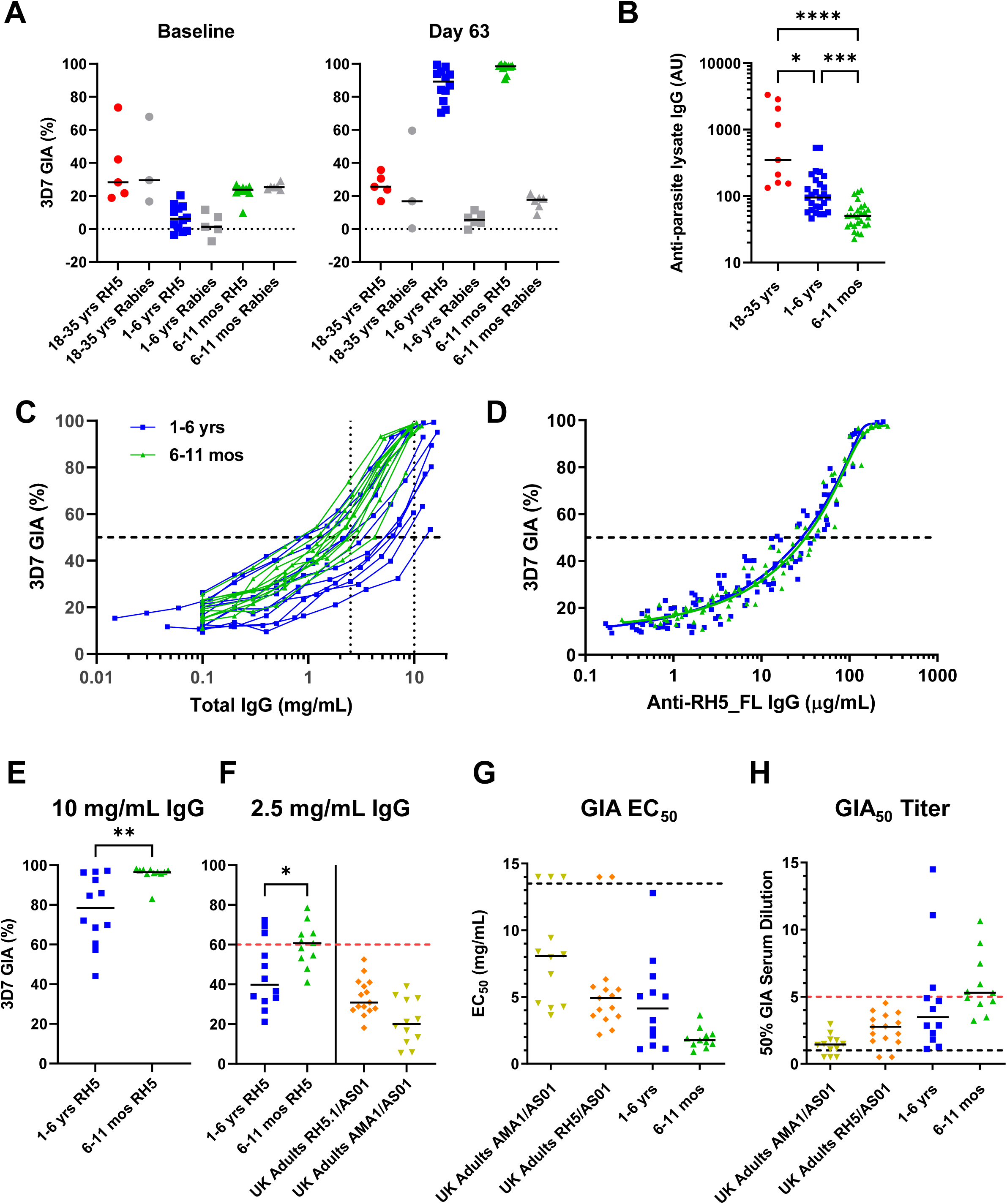
Functional GIA induced by ChAd63-MVA RH5 vaccination. (**A**) *In vitro* growth inhibition activity (GIA) of purified IgG was assessed against 3D7 clone *P. falciparum* parasites. Total IgG purified from serum at baseline (day 0) and the day 63 time-point was initially tested at each sample’s physiological total serum IgG concentration as measured by HPLC (**Figure S6A**). Data shown for Group 1 adults 18-35 years, N=5 vaccinees and N=3 control; Group 2B children 1-6 years, N=12 vaccinees and N=5 or 6 control (one sample was lost during analysis); and Group 3B infants 6-11 months, N=11 vaccinees and N=6 control. (**B**) Median and individual anti-parasite lysate serum IgG responses as measured by ELISA in arbitrary units (AU) in pre-vaccination (day 0) samples. Group 1 adults 18-35 years, N=9; Group 2A and 2B children 1-6 years, N=27; Group 3A and 3B infants 6-11 months, N=27. Analysis using Kruskal-Wallis test with Dunn’s multiple comparison test; \**P* < 0.05, \*\*\**P* < 0.001, \*\*\*\**P* < 0.0001. (**C**) Day 63 samples from Group 2B children 1-6 years (N=12) and Group 3B infants 6-11 months (N=11) vaccinated with ChAd63-MVA RH5 were subsequently titrated in the GIA assay using a 2-fold dilution series and starting at each sample’s physiological total serum IgG concentration. GIA responses were later interpolated for 2.5 and 10 mg/mL total IgG (dotted lines), and the GIA assay EC_50_ of each purified IgG in mg/mL was also interpolated from the data, i.e. the concentration of total IgG that, following titration, showed 50% GIA (dashed line). (**D**) Relationship between GIA data from the dilution series shown in (**C**) and concentration of anti-RH5_FL purified IgG used in the assay as measured by ELISA. Non-linear regression curves are shown for all samples combined in Group 2B (blue line, r^2^ = 0.92, N=108) and in Group 3B (green line, r^2^ = 0.95, N=81). The EC_50_ (concentration of anti-RH5_FL polyclonal IgG that gives 50% GIA, dashed line) was calculated. (**E**) GIA data interpolated from (**C**) at 10 mg/mL total IgG and (**F**) at 2.5 mg/mL total IgG. Analysis using Mann-Whitney test comparing Groups 2B and 3B only; \**P* < 0.05, \*\**P* < 0.01. Historical GIA assay data are also shown for comparison only in (**F**) for the RH5.1/AS01_B_^17^ (N=15) and AMA1 FMP2.1/AS01 ^18^ (N=12) protein-in-adjuvant vaccines previously tested in healthy UK adults. (**G**) Individual GIA assay EC_50_ of each purified IgG in mg/mL interpolated from the data in (**C**). Samples for which the GIA was <50% at the top concentration of total IgG tested were plotted as 14 mg/mL (above the dashed black line). (**H**) To relate the GIA assay results (using purified total IgG) back to the original sera, the concentration of IgG in each original serum sample was also measured by HPLC (**Figure S6A**). This enabled calculation of the “GIA_50_ Titer”, defined previously ^9^ as the dilution factor of each serum sample required to reach the concentration of purified IgG that gives 50% GIA (i.e. the GIA EC_50_). Samples for which the GIA_50_ titer could not be calculated (because they did not achieve ≥50% GIA using purified IgG) are plotted arbitrarily at 0.8. In (**G**) and (**H**), individual data and median results are shown for the same samples shown in (**F**). Red dashed lines indicate threshold level of GIA at 2.5 mg/mL total IgG in (**F**) and a GIA_50_ titer > 5 in (**H**) above which protection against a stringent *P. falciparum* blood-stage challenge has been reported in *Aotus* monkeys vaccinated with RH5 ^9^.

We next titrated individual total IgG samples from the high responding pediatric groups using a 2-fold dilution series in the GIA assay, starting at each individual sample’s physiological total IgG concentration (**Figure 6C**). Here, the observed GIA showed a very strong relationship to the concentration of RH5-specific IgG present in the total IgG used in the assay as measured by ELISA (**Figure 6D**), as seen previously for this antigen following viral vectored or protein-in-adjuvant immunization of healthy UK adults ^11, 17^. The concentration of RH5_FL-specific polyclonal IgG required to give 50% GIA (EC_50_) was highly comparable in both age groups: 34 μg/mL (95% confidence interval [CI], 25–48) in children and 39 μg/mL (95% CI 31–53) in the infants. This functional “quality” readout of vaccine-induced RH5-specific IgG was highly similar to that previously reported in RH5 vaccinated healthy UK adults ^11, 17^, which we also confirmed in an independent head-to-head repeat (**Figure S6C**).

Data from previous *Aotus* monkey *P. falciparum* challenge studies have suggested that levels of *in vitro* GIA >60% at 2.5 mg/mL purified total IgG are associated with a protective outcome following blood-stage vaccination ^9, 19^. We therefore next interpolated the levels of GIA at 10 mg/mL and 2.5 mg/mL total IgG for both age groups, which showed these were significantly higher in the 6-11 month old infants (**Figure 6E,F**). Notably, 6/11 vaccinated infants showed >60% GIA at 2.5 mg/mL total IgG (median 61%, range 41-78%). Finally, to relate the GIA assay results back to the original sera, and to account for the significant differences in the physiological concentrations of total IgG across the age groups (**Figure S6A**), we calculated the “GIA_50_ titer”, defined in previous studies ^9, 11, 17^ as the dilution factor of each serum sample required to reach the concentration of purified IgG that gives 50% GIA (i.e. the GIA EC_50_). Here, a GIA_50_ titer >5 was previously associated with protection against a stringent *P. falciparum* blood-stage challenge in *Aotus* monkeys vaccinated with RH5 ^9^. In this analysis of the GIA EC_50_ (**Figure 6G**) followed by GIA_50_ titer (**Figure 6H**), the Group 3B vaccinated 6-11 month old infants again showed the highest responses, with a median GIA_50_ titer = 5.3 (range 3.2-10.6). In summary, these GIA responses in the vaccinated 6-11 month old infants are the highest levels of GIA reported to-date following human vaccination, and were higher than levels of GIA achieved in healthy UK adults using highly immunogenic recombinant protein-in-adjuvant vaccine formulations targeting the RH5 or AMA1 antigens (**Figure 6F–H**) ^17, 18^_._

## Discussion

This dose-escalation, age de-escalation, double-blind, randomized, controlled Phase 1b trial reports the first data in a malaria-endemic population for a vaccine targeting the RH5 antigen from the blood-stage *P. falciparum* merozoite. We show in healthy Tanzanian adults, children and infants that a recombinant ChAd63-MVA heterologous prime-boost immunization regimen has a favorable safety profile, and can induce robust functional RH5-specific serum antibody responses in addition to B cell responses in the target infant age group. The local and systemic reactogenicity profile of the full dose RH5 vaccines in all three age groups was similar, if not reduced, as compared to that seen in healthy UK adults immunized with the identical vaccines ^11^. These data are consistent with previous Phase 1a/b malaria vaccine trials using the same viral vector delivery platforms at similar doses ^16, 20–30^.

The clinical safety of MVA as a recombinant vaccine vector for other infectious diseases and cancer is well documented ^31^, whilst current efforts are also using this virus to vaccinate against mpox ^32^. Our data with the ChAd63 vector also add support to existing data that suggest this simian adenovirus vector is safe for clinical use. However, a very rare but serious adverse reaction to a similar adenovirus vector (ChAdOx1) has been observed in the context of COVID-19 vaccines – vaccine-induced thrombosis with thrombocytopenia ^33^. The mechanism of this is not completely understood and so it is unclear whether this risk is likely to apply to another serotype of adenovirus, delivering a non-coronavirus antigen to a predominantly African target population. Reassuringly this phenomenon has not been observed in any trials to-date of the same serotype of adenovirus (ChAdOx1) delivering a malarial or other non-coronavirus antigen, nor has it been observed in trials of other serotypes ^34^. However, all these trials have been limited by their size (none reaching Phase 3 or more than hundreds of recipients) and so such a rare adverse event, even if it were real, may not have been detected. Nevertheless, since the time of undertaking this trial, a second Phase 1b trial has initiated in Tanzania using a soluble protein-in-adjuvant formulation, RH5.1/Matrix-M™ (ClinicalTrials.gov NCT04318002) ^17, 35^. Future efforts will thus focus on the clinical development of recombinant RH5 antigen-based vaccines formulated in adjuvant, rather than ChAd63-MVA viral vectors, in order to align blood-stage vaccine delivery with the existing anti-sporozoite vaccines RTS,S/AS01 ^36^ and R21/Matrix-M™ ^2^ and to enable future multi-stage malaria vaccine strategies.

The viral vectored vaccine platform was historically developed to induce T cell responses against the encoded transgene ^37^, and in this trial RH5-specific IFN-γ T cell responses were induced by ChAd63 RH5 in the Tanzanian adults; these subsequently peaked at median levels of >1000 SFU/million PBMC following the MVA RH5 boost. The magnitude and maintenance of these responses post-boost were largely comparable to those seen previously in healthy UK adults immunized with the identical vaccine regimen ^11^. In contrast, T cell priming by ChAd63 RH5 in the 6-11 month old infants, and boosting by MVA RH5 in both the infants and the 1-6 year old children appeared much weaker, leading to significantly lower responses (∼5-10-fold) post-boost as compared to adults when using the standard ELISPOT readout that reports per million PBMC. However, age-dependent variations are observed in the numbers of lymphocytes circulating per mL of blood, with much higher lymphocyte frequencies measured in young children and infants as compared to adults ^38^. Following incorporation of the lymphocyte count to report ELISPOT responses per mL blood, only the 6-11 month old infants still showed significantly lower responses post-prime, whilst far more comparable IFN-γ T cell responses were observed across the age groups post-boost, as reported in previous West African Phase 1b clinical trials using the same vectors recombinant for the pre-erythrocytic malaria antigen ME-TRAP ^12^. Nonetheless, the possible contribution of IFN-γ-secreting T cells to vaccine-induced blood-stage malaria immunity in humans remains unclear. The ChAd63-MVA viral vectors have routinely induced a mixed antigen-specific CD4^+^/CD8^+^ T cell response in humans, and it is highly likely that RH5-specific CD4^+^ T cells will provide key help to B cell responses ^39, 40^. Indeed, previous work in UK adults has shown the ChAd63-MVA RH5 vaccines can induce antigen-specific peripheral T follicular helper (Tfh) cell responses ^41^. Work therefore remains on-going to investigate the phenotypes of the Tfh cells induced in the Tanzanian population and their contribution to vaccine-induced humoral immune responses.

We also optimized the design and delivery of these recombinant ChAd63 and MVA viruses to induce strong antibody responses against blood-stage malaria antigens ^42–44^. Here, the ChAd63-MVA RH5 vaccines induced RH5-specific IgG antibody responses in the Tanzanian adults that peaked at ∼14 µg/mL; these levels of serum antibody were highly comparable to those seen previously in healthy UK adults immunized with the identical vaccine regimen ^11^. More encouragingly, the levels of anti-RH5 IgG induced by the same full-dose vaccines in the pediatric groups were significantly higher, reaching peak levels of ∼150 µg/mL in the 6-11 month old infants following the MVA RH5 boost. The ability to induce such high levels of anti-RH5 IgG in the target age group for a blood-stage malaria vaccine (i.e. young children and infants over 5 months of age) bodes well for future field efficacy testing of standalone blood-stage malaria vaccines as well as multi-stage malaria vaccine strategies whereby a blood-stage RH5 component could be combined with existing pre-erythrocytic vaccines such as RTS,S/AS01 ^36^ or R21/Matrix-M™ ^2^. The data also support the ongoing and wider use of the chimpanzee adenovirus vaccine platform to protect against outbreak or endemic viruses, such as Ebola, rabies and Rift Valley fever ^45–47^.

The induction of higher antibody levels in the pediatric groups, as compared to adults, is likely not specific to the ChAd63-MVA RH5 vaccine. Indeed, although numbers were small, we observed the same age-dependent hierarchy of antibody induction against the rabies virus glycoprotein in the rabies vaccine controls. Elsewhere, younger children (aged 6-11 years) also induced higher antibodies than older children (aged 12-17 years) vaccinated with the ChAdOx1-nCoV19 (AZD1222) Covid-19 vaccine ^48^, and trials of the pre-erythrocytic vaccine ChAd63-MVA ME-TRAP also reported ten-fold higher anti-TRAP antibody levels in West African infants as compared to UK adults and West African adults ^12^. Published immunogenicity data spanning adults and infants 5-17 months of age vaccinated with the RTS,S/AS01 ^36, 49^ and R21/Matrix-M™ ^2, 50^ malaria vaccines also suggest the same. However, responses in infants 6-12 weeks of age at first vaccination with RTS,S/AS01 showed lower levels of antibody that associated with reduced efficacy against clinical malaria ^36^. Consequently, given both anti-sporozoite and anti-merozoite malaria vaccine strategies necessitate very high levels of antibody to protect against parasite infection, current efforts remain focused on infants and young children over 5 months of age at the time of first vaccination.

Why the infants and young children vaccinated with ChAd63-MVA RH5 induced such high levels of antibody remains to be fully understood. Our results suggested that pre-existing anti-vector immunity is unlikely to be the reason explaining the observed improvement in anti-RH5 humoral immunogenicity in the younger age groups. Consistent with this, studies of the ChAdOx1-nCoV19 (AZD1222) Covid-19 vaccine given in a 2-dose homologous regimen reported similar findings across studies in adults and children, with no evidence that anti-ChAd immune responses measured after the first vaccine dose associated with the immunogenicity outcome measures of the second vaccine dose ^48, 51^. We therefore sought to analyse the underlying B cell responses. Here, we observed higher peripheral ASC responses in the infants, as well as higher absolute numbers of B cells per microliter of blood and a stronger RH5-specific IgG^+^ B cell response within the CD19^+^ population in the younger age groups. The significantly higher anti-RH5 serum IgG response induced by the ChAd63-MVA RH5 vaccine in children and infants, as compared to adults, is therefore strongly associated with greater B cell immunogenicity. Given current antibody-inducing vaccine strategies to protect against clinical malaria are focussed on infants 5-17 months of age, this observation warrants further investigation in the future.

Finally, we assessed functional GIA of the vaccine-induced anti-RH5 antibodies. Our previous work has identified the *in vitro* assay of GIA as a highly significant predictor of *P. falciparum in vivo* growth inhibition following blood-stage challenge of both vaccinated UK adults ^17^ and *Aotus* monkeys ^9, 19^. We have also confirmed this association as a mechanistic correlate in *Aotus* monkeys, i.e. one that can cause *in vivo* protection, via passive transfer of a GIA-positive RH5-specific IgG monoclonal antibody ^10^, with similar results observed in humanized mice ^52^. Importantly, full protection of *Aotus* monkeys required a serological threshold level of GIA, defined as i) a level of *in vitro* GIA >60% at 2.5 mg/mL purified total IgG ^9, 19^, or ii) a GIA_50_ titer >5, with this latter measure also taking into account any differences in the physiological concentrations of total IgG in the vaccinees ^9^. Previous trials in healthy UK adults did not exceed this threshold when using the ChAd63-MVA RH5 vaccine ^11^, or a more immunogenic soluble RH5 protein-in-adjuvant, RH5.1/AS01_B_ ^17^. In contrast, the 6-11 month old infants in this trial showed the highest yet reported levels of GIA in vaccinated humans, with over half of the vaccinees in this target age group exceeding the threshold. This related to the high quantity of anti-RH5 serum IgG induced by the ChAd63-MVA RH5 vaccine, given the functional quality (i.e. GIA per unit anti-RH5 antibody) was highly similar across the age groups tested. Moreover, the highly similar functional antibody quality data across the UK and Tanzanian vaccine trials are encouraging because they indicate no obvious interference from pre-existing and naturally-occurring anti-malarial antibody responses with the vaccine-induced anti-RH5 IgG, as occurred for historical vaccines targeting AMA1 ^53^.

The main limitations of the trial include relatively small numbers of participants and a limited follow-up period of four months post-MVA RH5 booster vaccination, such that the longer-term kinetic of the immune response has not been characterized. Improving the durability of protection against clinical malaria will be critical for next-generation malaria vaccine strategies. We also only tested a single heterologous prime-boost regimen in this trial, whereas a more extensive assessment of vaccine dose and regimen, including the use of delayed boosting, in Phase 1a/b trials could be key to optimizing immunogenicity and the longevity of vaccine-induced protection ^17, 54^. In this regard, since undertaking this trial, a second Phase 1b trial has initiated in Tanzania using a soluble protein-in-adjuvant formulation, RH5.1/Matrix-M™ delivered in a variety of dosing regimens (ClinicalTrials.gov NCT04318002). This trial will indicate whether even higher antibody and GIA levels can be achieved in the target 5-17 month old infant population with a delivery platform that is more immunogenic than ChAd63-MVA. Nonetheless, the data in the Phase 1b trial reported here confirm, for the first time, that substantial anti-RH5 immune responses can be achieved safely by vaccination in infants from a malaria-endemic area, in stark contrast to the poor immunogenicity seen to this antigen following natural *P. falciparum* infection ^6, 11^. These data also justify onward progression to Phase 2b field efficacy trials to determine whether growth inhibitory antibody levels of this magnitude can ultimately protect against clinical malaria.

## Methods

### Study Design

This was a randomized, controlled, age de-escalation, dose-escalation study called VAC070 that was conducted according to the principles of the current revision of the Declaration of Helsinki 2008 and in full conformity with the ICH guidelines for Good Clinical Practice (GCP). It was approved by the Oxford Tropical Research Ethics Committee in the UK (OxTREC, reference 29-17), the Ifakara Health Institute Institutional Review Board in Tanzania (reference: 20-2017), the National Institute for Medical Research in Tanzania, the National Health Research Ethics Sub-Committee (NatHREC) and the then Tanzania Food and Drugs Authority (now the Tanzania Medicines and Medical Devices Authority), reference: TFDA0017/CTR/0015/3. The Consolidated Standards of Reporting Trials (CONSORT) guideline was followed.

We report here the safety, reactogenicity and immunogenicity profile of heterologous prime-boost ChAd63-MVA RH5 vaccination up until 168 days post-enrolment. The study was conducted (and participants recruited and vaccinated) at Kingani Clinical Facility, Ifakara Health Institute, Bagamoyo branch, Tanzania. The VAC070 trial was registered on ClinicalTrials.gov (NCT03435874), the Pan-African Clinical Trials Registry (PACTR201710002722229) and ISRCTN (ISRCTN47448832).

### Participants

Healthy adults (18-35 years), young children (1-6 years) and infants (6-11 months) residing in Bagamoyo, Tanzania, with a negative malaria blood film at screening, were eligible for inclusion in the study and enrolled into three groups according to age. A full list of inclusion and exclusion criteria are listed in the study protocol which is included within the Supplementary Appendix. Each participant (or guardian) signed or thumb-printed an informed consent form at the in-person screening visit and consent was verified before each vaccination. A Safety Monitoring Committee (SMC) periodically reviewed the study progress and safety data according to a safety review schedule, critically timed to age de-escalations and dose escalations.

### Procedures

#### Randomization and Masking

A randomization list was generated by an independent statistician. This contained sequential codes (Treatment numbers) linking a study identification (ID) to a vaccine assignment. Study ID was assigned to participants in the order in which they were enrolled in the trial. Access to the randomization list was exclusively limited to the study pharmacist and the independent statistician(s). These individuals had no role in the evaluation of the study participants.

Participants were assigned to groups based on their age, and groups (characterized by participant age and dose of vaccine) were enrolled sequentially. Randomization into vaccine or control groups was performed according to a 2:1 ratio.

Data pertaining to ChAd63 RH5, MVA RH5 or rabies vaccine were collected in a double-blinded manner. Neither the vaccine recipient nor their parent(s)/guardian(s) or those members of the study team responsible for administering the vaccines or evaluating safety and immunogenicity endpoints were aware of individual vaccine allocation. Only those staff responsible for the storage and preparation of vaccines were unblinded as both vaccines were distinguishable by their packaging and labelling; these staff played no other role in the study and the vaccine preparation area was kept physically separate from the immunization area. The Local Independent Safety Monitor was provided with sealed code-break envelopes for each participant to facilitate unblinding for urgent clinical/ethical reasons. They also had access to a copy of the master randomization list in a sealed envelope, in case emergency unblinding was required.

Participants attended a two-part screening visit and those eligible returned for enrolment and were randomized to either a dose of ChAd63 RH5 or rabies control vaccine. Eight weeks later all participants were then randomized to either a dose of MVA RH5 or rabies vaccine. All vaccines were administered by intramuscular injection in the upper arm.

### Safety Analysis

Following each vaccination, each participant was visited at home on days 1, 3, 4, 5 and 6 by a community health worker for assessment and recording of any solicited and unsolicited AEs. At days 2, 7, 14 and 28 post-vaccination participants were seen at the clinical research facility. Observations (heart rate, temperature and blood pressure measurements) were taken at the clinic visits from the day of vaccination until the 28 day follow-up visit. Blood tests for exploratory immunology were taken at all visits except those occurring 2 days after each vaccination, 7 days after the first vaccination and 14 days after the second vaccination. Blood samples for safety (full blood count, alanine aminotransferase (ALT) and creatinine) were carried out at screening and on days 0, 7, 14, 28, 56, 63, 84 and 168 for all groups. Any solicited AEs occurring during the 7 days post-vaccination were defined as being at least possibly related to vaccination. The likely causality and grading of all other AEs were assessed as described in the protocol. All unsolicited AEs are reported (**Table S2**) but none were considered possibly, probably or definitely related to vaccination. The types of AEs were classified according to MedDRA (version 26.0).

### Outcomes

Primary outcome measures for vaccine safety included numbers of solicited and unsolicited AEs after each vaccination. The primary outcome analysis was conducted on the safety analysis population and included participants who received at least the first dose of vaccine in the study. The maximum severity for each solicited systemic AE across seven days after first and second vaccinations was derived for each participant and summarized by group. Analyses were conducted similarly for local reactogenicity. Serious adverse events (SAEs) were collected for the entire study period. The secondary outcome measures for humoral immunogenicity were the concentration of anti-RH5 serum antibodies by ELISA and their percentage GIA *in vitro* using purified IgG, and for cellular immunogenicity were T and B cell responses to RH5 as measured by ELISPOT and/or flow cytometry.

### Vaccines

The design, production and preclinical testing of the viral vector vaccines have been reported previously in detail ^6, 9^. Briefly, both recombinant viruses express the same 1503 bp coding sequence of RH5 from the 3D7 clone of *P. falciparum*, aa E26–Q526 (NCBI Accession #XM_001351508.1). ChAd63 RH5 was manufactured by Advent, Pomezia, Italy which is a daughter company of ReiThera. This production facility meets current Good Manufacturing Practice (cGMP) requirements of the US Food and Drug Administration (FDA) and the European Medicines Agency (EMA) to produce investigational vaccines to be used in human clinical studies. MVA RH5 was manufactured under cGMP conditions by IDT Biologika GmbH, Germany, as described in detail previously ^11^. Control participants received VERORAB, an inactivated rabies vaccine (Sanofi Pasteur).

## Peripheral Blood Mononuclear Cell (PBMC), Plasma and Serum Preparation

Blood samples were collected into lithium heparin-treated vacutainer blood collection systems (Becton Dickinson, UK). PBMC were isolated and used within 6 hours in fresh assays as previously described ^22^. Excess cells were frozen in foetal calf serum (FCS) containing 10% dimethyl sulfoxide (DMSO) and stored in liquid nitrogen. Plasma samples were stored at –80 °C. For serum preparation, untreated blood samples were stored at room temperature (RT) and then the clotted blood was centrifuged for 5 min (1000 *xg*). Serum was stored at –80 °C.

## Peptides

Peptides for *ex-vivo* IFN-γ ELISPOT were purchased from NEO Scientific (Cambridge, MA, USA). Sequences are reported in **Table S4**. In brief, the peptides (20 amino acids (aa) in length and overlapping by 10 aa) covered the entire RH5 sequence present in the RH5 vaccine. Peptides were reconstituted in 100% DMSO at 50-200 mg/mL and combined into various pools for the ELISPOT assay.

## *Ex-vivo* IFN-γ ELISPOT

Fresh PBMC were used in all assays using a previously described protocol ^11^. Spots were counted using an ELISPOT counter (Autoimmun Diagnostika (AID), Germany). Results are expressed as IFN-γ spot-forming units (SFU) per million PBMC. Background responses in unstimulated control wells were almost always less than 20 spots, and were subtracted from those measured in peptide-stimulated wells.

## Recombinant RH5 Protein

Recombinant full-length RH5 protein (also known as “RH5.1”) was used for all ELISA assays and B cell ELISPOT assays. The protein was produced and purified from a stably transfected *Drosophila* S2 cell line as previously described ^35^.

## RH5 ELISA

Anti-RH5 total IgG ELISAs were performed against full-length RH5 protein (RH5.1) using standardized methodology as previously described in detail for other RH5 vaccine trials ^11, 17^. The reciprocal of the test sample dilution giving an optical density at 405nm (OD_405_) of 1.0 in the standardized assay was used to assign an ELISA unit value of the standard. A standard curve and Gen5 ELISA software v3.04 (BioTek, UK) were used to convert the OD_405_ of individual test samples into arbitrary units (AU). These responses in AU are reported in µg/mL following generation of a conversion factor by calibration-free concentration analysis (CFCA) as reported previously ^11^.

## ChAd63 ELISA

Antibody responses to the ChAd63 vaccine vector were determined by endpoint ELISA as previously described ^42, 55, 56^. Briefly, a purified ChAd63 vector encoding an irrelevant antigen (ovalbumin) ^57, 58^ was adsorbed overnight at 4 °C to 96 well NUNC-Immuno Maxisorp plates (Thermo Fisher Scientific) at 3 × 10^8^ vp/mL ^59^. Test sera were diluted 1:100, added in duplicate and serially diluted 3-fold. Bound antibodies were detected using goat anti-human IgG conjugated to alkaline phosphatase (Sigma), developed using 4-Nitrophenyl phosphate disodium salt hexahydrate (Sigma) and OD_405_ determined on a BioTek Elx808 reader with Gen5 ELISA software. Endpoint titers were calculated as the x-axis intercept of the sample titration curve at OD_405_ = 0.15 (equivalent to blank test samples). A positive control sample from participants vaccinated with a different ChAd63 vectored vaccine ^27^ was included as an internal reference.

## Parasite Lysate ELISA

Serum antibody levels to parasite lysate were assessed by standardized ELISA methodology previously described ^60, 61^. Schizont extract from *P. falciparum* (3D7 clone) produced by the GIA Reference Laboratory, NIAID, NIH, was adsorbed overnight at 4 °C to 96 well NUNC-Immuno Maxisorp plates (Thermo Fisher Scientific) at equivalent to 5×10^2^ parasites per µL. Test sera were diluted in 1% milk and added in triplicate to plates following blocking with 5% milk in DPBS (Sigma). A reference standard and internal control from a pool of N=34 high malaria pre-exposed serum samples, from the BCTF-IHI Biobank, plus blank wells were included. Bound antibodies were detected using goat anti-human IgG conjugated to alkaline phosphatase (Sigma), developed using 4-Nitrophenyl phosphate disodium salt hexahydrate (Sigma) and absorbance (OD_405_) was determined on a BioTek Elx808 reader with Gen5 software. Antibody units were assigned using the reciprocal dilution of the standard giving an optical density of 1.0 at OD_405_. The standard curve and Gen5 software v3.04 (Agilent) were then used to convert the OD_405_ of test samples to arbitrary units (AU).

## Rabies ELISA

Binding antibody responses induced by the rabies control vaccine were determined by endpoint ELISA, with the same methodology as for the anti-ChAd63 ELISA, but using recombinant rabies glycoprotein ^46^ adsorbed to the plate at 2 µg/mL. Pooled sera from Group 3B participants vaccinated with the rabies vaccine was included as a development control.

## Parasite qPCR

Parasitemia was determined retrospectively by quantitative polymerase chain reaction (qPCR) performed on blood samples taken at baseline, 7 days and 28 days post-boost as previously described ^62^. Briefly, blood was collected in 2.0 mL tubes containing EDTA. DNA was extracted from whole blood using Quick-DNA miniprep plus kit (Zymo research, USA), and 2 µL each extracted DNA was used per assay well and run in triplicate. qPCR was conducted on a CFX96 real-time qPCR machine (Bio-Rad) and analyzed with CFX Manager Software (v2.2) with the following cycling conditions: polymerase activation at 95 °C for 1 min, 45 cycles of denaturation at 95 °C for 15 s and annealing and elongation at 57 °C for 45 s. Parasites per µL were calculated against a defined international standard for *P. falciparum* DNA Nucleic Acid Amplification techniques (WHO reference from NIBSC #04/176) ^63^ reconstituted in 0.5 mL sterile nuclease-free water to 5×10^8^ parasites per µL.

## Antibody-Secreting Cell (ASC) ELISPOT

*Ex-vivo* ASC ELISPOT assays were performed against RH5.1 protein as described in detail elsewhere ^11^ using fresh PBMC. Plates were counted using an AID ELISPOT plate reader. Results are reported as RH5-specific ASC per million PBMC used in the assay.

## B cell Flow Cytometry

Frequencies of live B cells within total lymphocytes and RH5-specific cells within IgG^+^ B cells (CD19^+^ IgD^−^ IgM^−^ IgA^−^) were measured by flow cytometry. In brief, cryopreserved PBMC were thawed and washed in IMDM (12440053, Gibco) supplemented with 10% FCS (F9665, Sigma Aldrich), 0.2% MycoZap (VZA-2031, Lonza), and 0.04% benzonase (71205-3, Merck). Thawed PBMC were stained first with a viability stain (Live/Dead Aqua; L34966, Invitrogen), followed by a panel comprising anti-human CD3-BV510 (317332, Biolegend); anti-human CD14-BV510 (301842, Biolegend); anti-human CD56-BV510 (318430, Biolegend); anti-human CD27-A488 (393204, Biolegend); anti-human IgM-PerCP-Cy5.5 (561285, BD Biosciences); anti-human CD19-ECD (IM2708U, Beckman Coulter); anti-human IgD-PE-Cy7 (561315, BD Biosciences); anti-human IgA-A647 (109-475-011, Jackson); anti-human CD38-APC-Cy7 (303534, Biolegend); and a RH5-PE probe (produced in-house as previously described ^41, 54^). Stained samples were washed and acquired on a FACSAria Fusion Flow Cytometer (BD Biosciences). See **Figure S5** for an example gating strategy of live B cells within single lymphocytes, and RH5-specific (IgG^+^) cells within the CD19^+^ IgD^−^ IgM^−^ IgA^−^ B cell population.

## Haematology

The lymphocyte count was obtained during Complete Blood Count (CBC) analysis using the automated haematology analyzer, Sysmex XS 800i, for assessment of whole blood. The analyzer uses a fluorescence flow cytometry method for analysis of white blood cells (WBC) and the five differentials by using the semiconductor laser beam. A small volume of blood (20 µL) is aspirated by the analyzer to measure the WBC differentials according to their size and structure.

## Serum IgG Concentration

Total serum IgG concentrations were determined using a Bio-Monolith Protein G column on an Agilent 1260 HPLC system (Agilent, Cheshire, UK). Separation was performed at 1 mL/min using PBS and 0.2 M Glycine pH 2.0 as mobile phases with detection at UV 280 nm. A calibration curve was produced using purified human IgG.

## Assay of Growth Inhibition Activity (GIA)

Standardized assays were performed by the GIA Reference Center, NIH, USA, using previously described methodology ^64^, with one modification. Here, each sample was tested in three independent replication assays using three different batches of red blood cells (RBC), and the median of these three results was used to generate the final dataset. Otherwise for each assay, in brief, protein G purified IgG samples were incubated with RBC infected with synchronized *P. falciparum* 3D7 clone parasites in a final volume of 40 µL for 40 h at 37 °C, and the final parasitemia in each well was quantified by biochemical determination of parasite lactate dehydrogenase. All purified IgG samples were tested at final test well concentrations (reported in mg/mL) as described in Results. For certain samples a dilution series was used to determine the concentration that gave 50% GIA (EC_50_).

## Statistical Analysis

Data were analyzed using GraphPad Prism version 9.5 for Windows (GraphPad Software Inc., California, USA). All tests used were two-tailed and are described in the text. To analyze the relationship between GIA and ELISA assay data, a Richard’s five-parameter dose-response curve was fitted, constrained to 0% GIA at the bottom and 100% GIA at the top. A value of *P* < 0.05 was considered significant.

## Author Contributions

Conceived and performed the experiments: SES, WFK, IMM, NSL, MM, FM, SA, AD, FM, BS, TA, MR, LM, OL, AMA, GN, BM, SM, TGM, JRB, LTW, YT, LDWK, SHH, ROP, CMN, CAL, KM, SJD, AMM, AIO. Analyzed the data: SES, WFK, IMM, NSL, TA, TGM, CMN, KM, SJD, AMM, AIO. Project Management: SA, AML, FLN, J-SC. Wrote the paper: SES, SJD, AMM.

## Data Availability

All data produced in the present study are available upon reasonable request to the authors

## Acknowledgments

The authors are grateful to Nick Edwards, Julie Furze, Mimi Hou, Jing Jin, Simon Kerridge, Penelope Lane, Daniel Marshall-Searson, Ian Poulton and Alex Spencer for assistance, and to Sandy Douglas and Adam Berg for provision of rabies glycoprotein (Jenner Institute, University of Oxford); Helena Parracho, Tanja Brenner and Eleanor Berrie (Clinical Biomanufacturing Facility, University of Oxford); Amy Duckett and Carly Banner for arranging contracts (University of Oxford); Robert Hedley and Vasiliki Tsioligka for assistance with flow cytometry (Sir William Dunn School of Pathology, University of Oxford); Loredana Siani, Stefania Di Marco (ReiThera SRL, formerly Okairòs SRL) and Annunziata Del Gaudio (Advent SRL) for manufacture of ChAd63 RH5; Marc Lievens, Danielle Morelle and GSK for review of the clinical trial design, supply of the ChAd63 vector and review of the draft manuscript, but the authors are solely responsible for the final content and data interpretation; the Independent Safety Monitoring Committee for their insightful safety oversight (Prof Brian Angus, Prof Karim Manji and Prof Alison Elliot); and all the study participants.

This work was funded in part by an African Research Leader Award to AIO from the UK Medical Research Council (MRC) [MR/P020593/1]. This award was jointly funded by the UK MRC and the UK Department for International Development (DFID) under the MRC/DFID Concordat agreement and is also part of the EDCTP2 programme supported by the European Union. This work was also supported in part by the National Institute for Health Research (NIHR) Oxford Biomedical Research Centre (BRC). The views expressed are those of the authors and not necessarily those of the NIHR or the Department of Health and Social Care. The GIA work was supported by the United States Agency for International Development (USAID) and the Intramural Program of the National Institute of Allergy and Infectious Diseases, National Institutes of Health. SHH holds an NIHR Academic Clinical Lectureship. CMN held a Sir Henry Wellcome Postdoctoral Fellowship [209200/Z/17/Z]. SJD is a Jenner Investigator and held a Wellcome Trust Senior Fellowship [106917/Z/15/Z].

## Data and Materials Availability

Requests for materials should be addressed to the corresponding authors.

