## Supplemental Information for "Superior antibody immunogenicity of a RH5 blood-stage malaria vaccine in Tanzanian infants as compared to adults"

#### Supplementary Figures

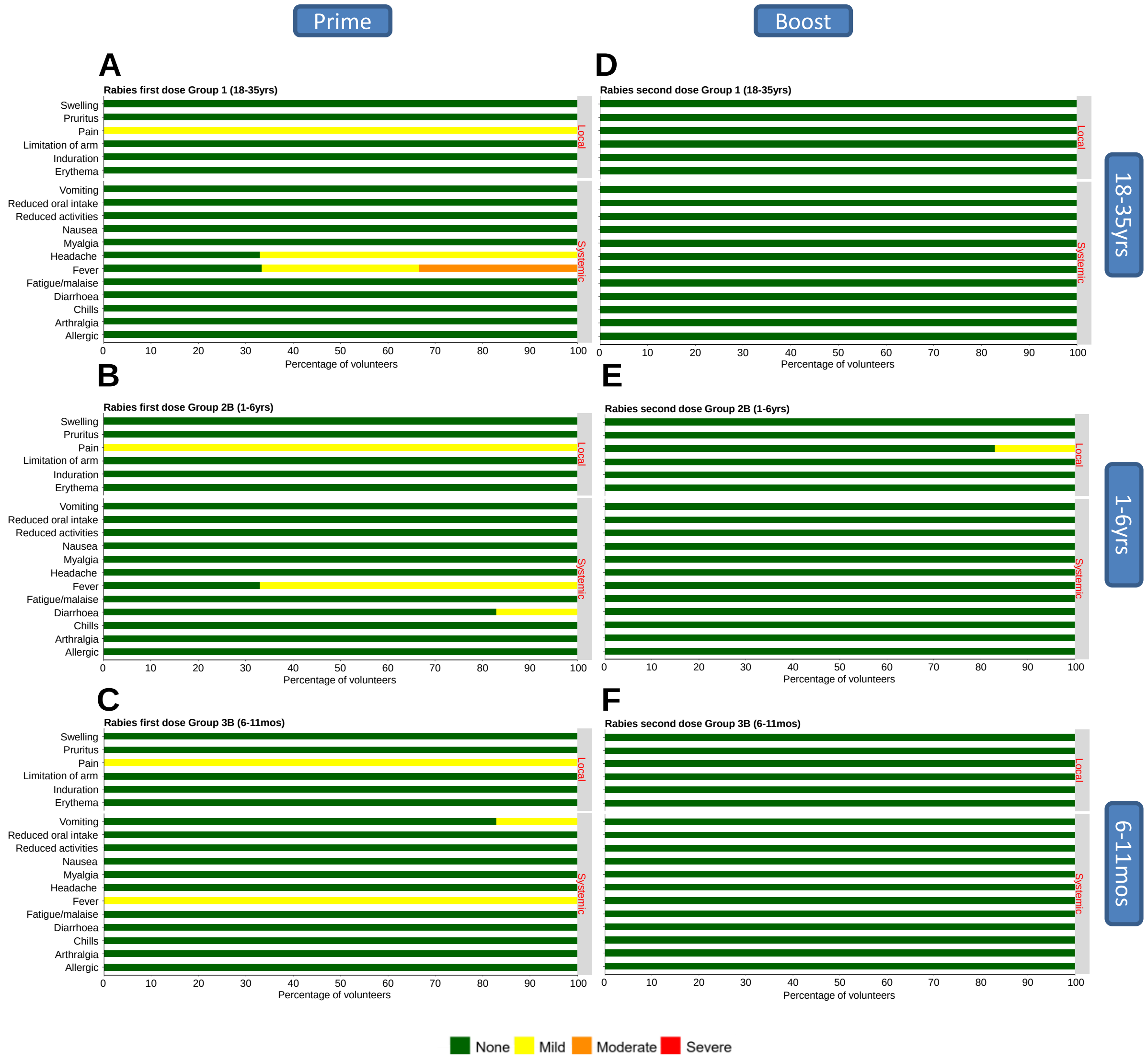

#### Supplementary Figure 1. Solicited AEs following vaccination with rabies vaccine control in Groups 1, 2B and 3B.

The solicited local and systemic AEs recorded for 7 days following prime and boost with rabies vaccine control are shown at the maximum severity reported by all volunteers within the VAC070 trial Groups 1, 2B and 3B. Volunteers aged (**A**) 18-35 years (n=3); (**B**) 1-6 years (n=6); and (**C**) 6-11 months (n=6) received rabies vaccine control on day 0, and the same volunteers aged (**D**) 18-35 years (n=3); (**E**) 1-6 years (n=6); and (**F**) 6-11 months (n=6) received a second dose of the same control on day 56.

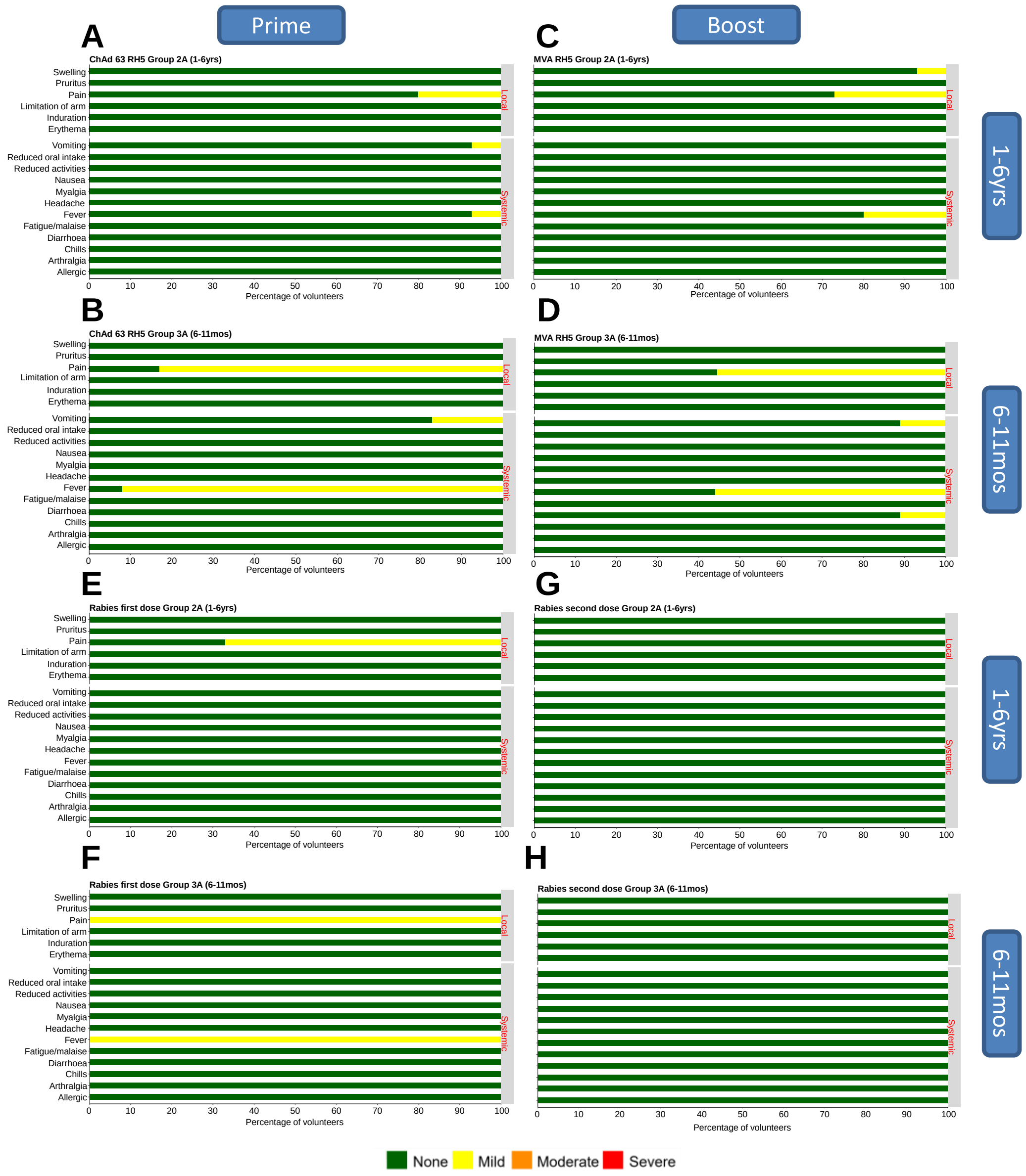

**Supplementary Figure 2. Solicited AEs following vaccination with a lower "lead in" dose of ChAd63 RH5 and MVA RH5 or rabies vaccine control in Groups 2A and 3A.** The solicited local and systemic AEs recorded for 7 days following the lower "lead in" dose of ChAd63 RH5 prime and MVA RH5 boost, or rabies vaccine control, are shown at the maximum severity reported by all volunteers within the VAC070 trial Groups 2A and 3A. Volunteers aged (**A**) 1-6 years (n=6); and (**B**) 6-11 months (n=6) received the lower "lead in" priming dose of 1 x $10^{10}$ vp ChAd63 RH5 on day 0. The same volunteers aged (**C**) 1-6 years (n=5); and (**D**) 6-11 months (n=6) received the lower "lead in" boosting dose of 1 x $10^{8}$ pfu MVA RH5 on day 56. Volunteers aged (**E**)

1-6 years (n=3); and (**F**) 6-11 months (n=3) received rabies vaccine control on day 0, and the same volunteers aged (**G**) 1-6 years (n=3); and (**H**) 6-11 months (n=3) received a second dose of the same control on day 56.

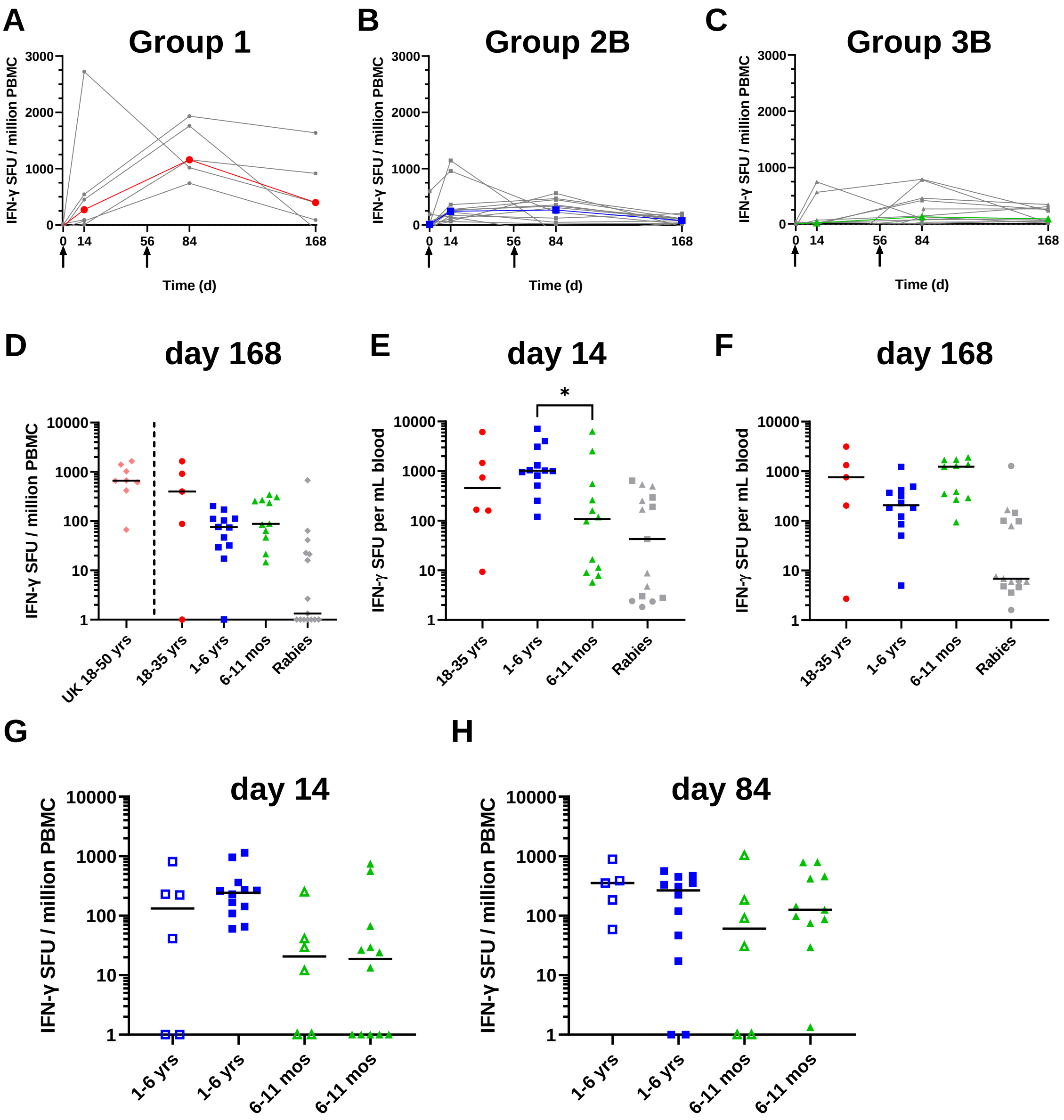

**Supplementary Figure 3. *Ex-vivo* IFN-γ ELISPOT data.**

Median (in color) and individual (in grey) *ex-vivo* IFN-γ ELISPOT responses to RH5 per million PBMC are shown over time (days 0, 14, 84 and 168) for all groups receiving the full dose of ChAd63-MVA RH5 vaccines. Responses are shown for (**A**) Group 1 adults 18-35 years, N=5 or 6; (**B**) Group 2B children 1-6 yrs, N=12; and (**C**) Group 3B infants 6-11 months, N= 11 or 12. (**D**) Median and individual *ex-vivo* IFN-γ ELISPOT responses to RH5 per million PBMC are shown for all groups receiving the full dose of ChAd63-MVA RH5 vaccines and rabies vaccine control at day 168, sixteen weeks post-MVA RH5 boost. Group 1 adults 18-35 years, N=5; Group 2B children 1-6 years, N=12; Group 3B infants 6-11 months, N=11; and rabies vaccine control, all groups and ages pooled, N=15. Historical data testing the identical dose and regimen of the ChAd63-MVA RH5 vaccine in healthy UK adults 18-50 years are shown for comparison only, N=8 (note in the UK trial these data were taken at day 140, the closest comparator time-point) [1]. (**E**) Median and individual *ex-vivo* IFN-γ ELISPOT responses to RH5 per mL peripheral blood are shown for all groups receiving the full dose of ChAd63-MVA RH5 vaccines and rabies vaccine control at day 14, and (**F**) day 168. Groups as per **Figure 3A** and panel (D), respectively. Analysis using Kruskal-Wallis test with Dunn's multiple comparison test across Groups 1, 2B and 3B only; $*P < 0.05$. (**G**) Median and individual *ex-vivo* IFN-γ ELISPOT responses to RH5 per million PBMC are also shown at day 14, two weeks post-ChAd63 RH5 prime, and (**H**) day 84, four weeks post-MVA RH5 boost in Group 2A children 1-6 years, N=5 or 6, and Group 3A infants 6-11 months, N=6, who received the lower "lead in" doses of ChAd63-MVA RH5 (open symbols) alongside those who received the full dose of vaccine in Groups 2B and 3B (closed symbols; data as per **Figure 3A,B**).

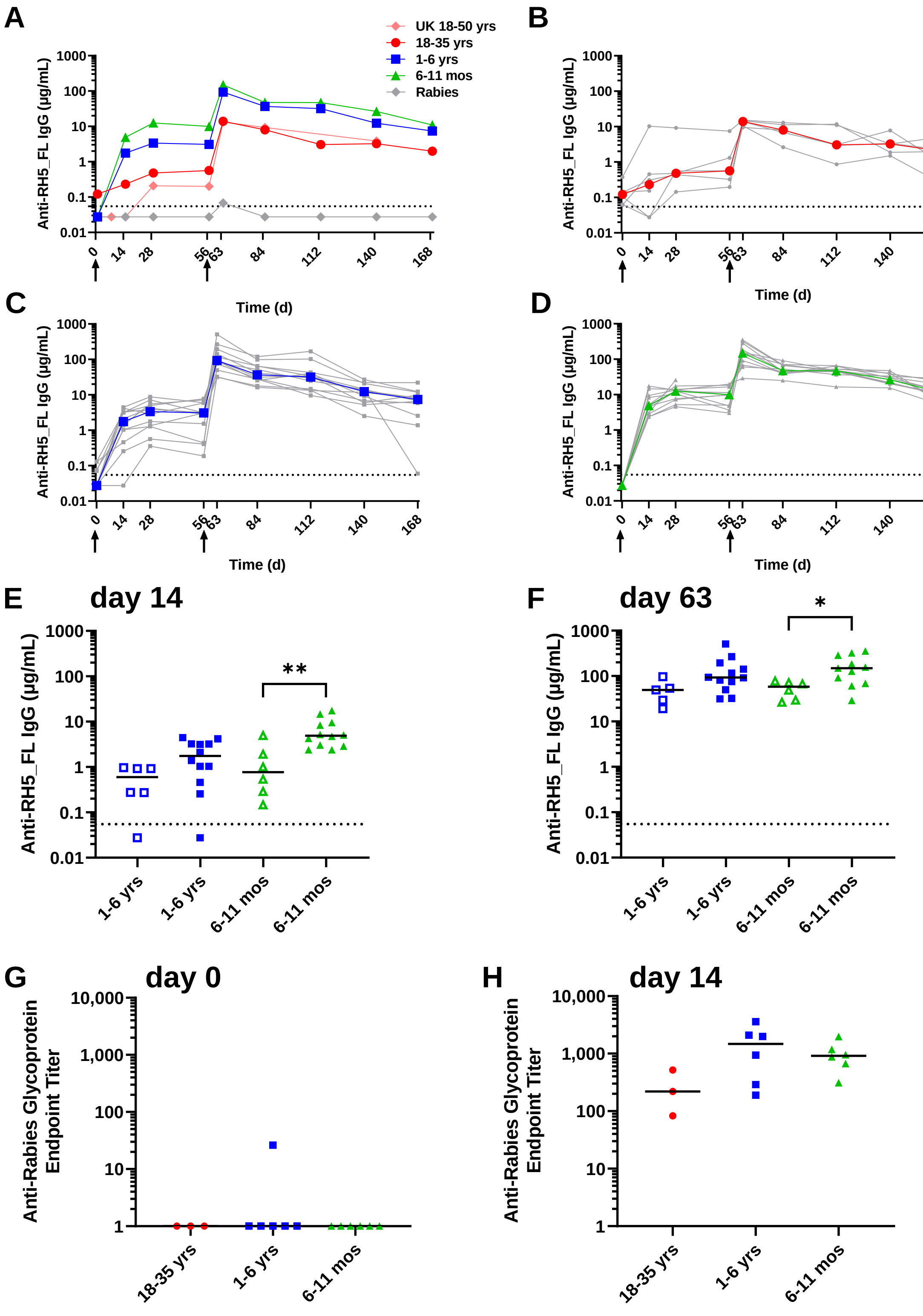

**Supplementary Figure 4. Serum antibody ELISA data.**

(**A**) Median anti-RH5_FL serum total IgG responses as measured by ELISA are shown over time for all groups receiving the full dose of ChAd63-MVA RH5 vaccines: Group 1 adults 18-35 years (red),

N=5 or 6; Group 2B children 1-6 yrs (blue), N=12; and Group 3B infants 6-11 months (green), N= 11 or 12; as well as rabies vaccine control (grey), all groups and ages pooled, N=15. Historical data testing the identical dose and regimen of the ChAd63-MVA RH5 vaccine in healthy UK adults 18-50 years (pink) are shown for comparison only: N=20 (up to day 56) and N=8 (onwards from day 63) [1]. (**B**) Median (in color) and individual (in grey) anti-RH5_FL serum total IgG responses as measured by ELISA are also shown over time for Group 1 adults 18-35 years, N=5 or 6, receiving the full dose of ChAd63-MVA RH5 vaccines, with the same also shown for (**C**) Group 2B children 1-6 yrs, N=12; and (**D**) Group 3B infants 6-11 months, N= 11 or 12. Arrows indicate vaccination time-points, and the horizontal dotted line indicates the limit of detection of the assay. Median and individual anti-RH5_FL serum total IgG responses as measured by ELISA are also shown at (**E**) day 14, two weeks post-ChAd63 RH5 prime, and (**F**) day 63, one week post-MVA RH5 boost in Group 2A children 1-6 years, N=5 or 6, and Group 3A infants 6-11 months, N=6, who received the lower "lead in" doses of ChAd63-MVA RH5 (open symbols) alongside those who received the full dose of vaccine in Groups 2B and 3B (closed symbols; data as per **Figure 4A,B**). Analysis using Kruskal-Wallis test with Dunn's multiple comparison test comparing only within Group 2 (2A vs 2B) and within Group 3 (3A vs 3B); *$P < 0.05$, **$P < 0.01$. (**G**) Median and individual anti-rabies glycoprotein serum total IgG responses as measured by ELISA in day 0 samples (pre-vaccination) and (**H**) day 14 samples (after one vaccination) are shown for individuals receiving rabies vaccine control: Group 1 adults 18-35 years, N=3; Group 2B children 1-6 years, N=6; Group 3B infants 6-11 months, N=6.

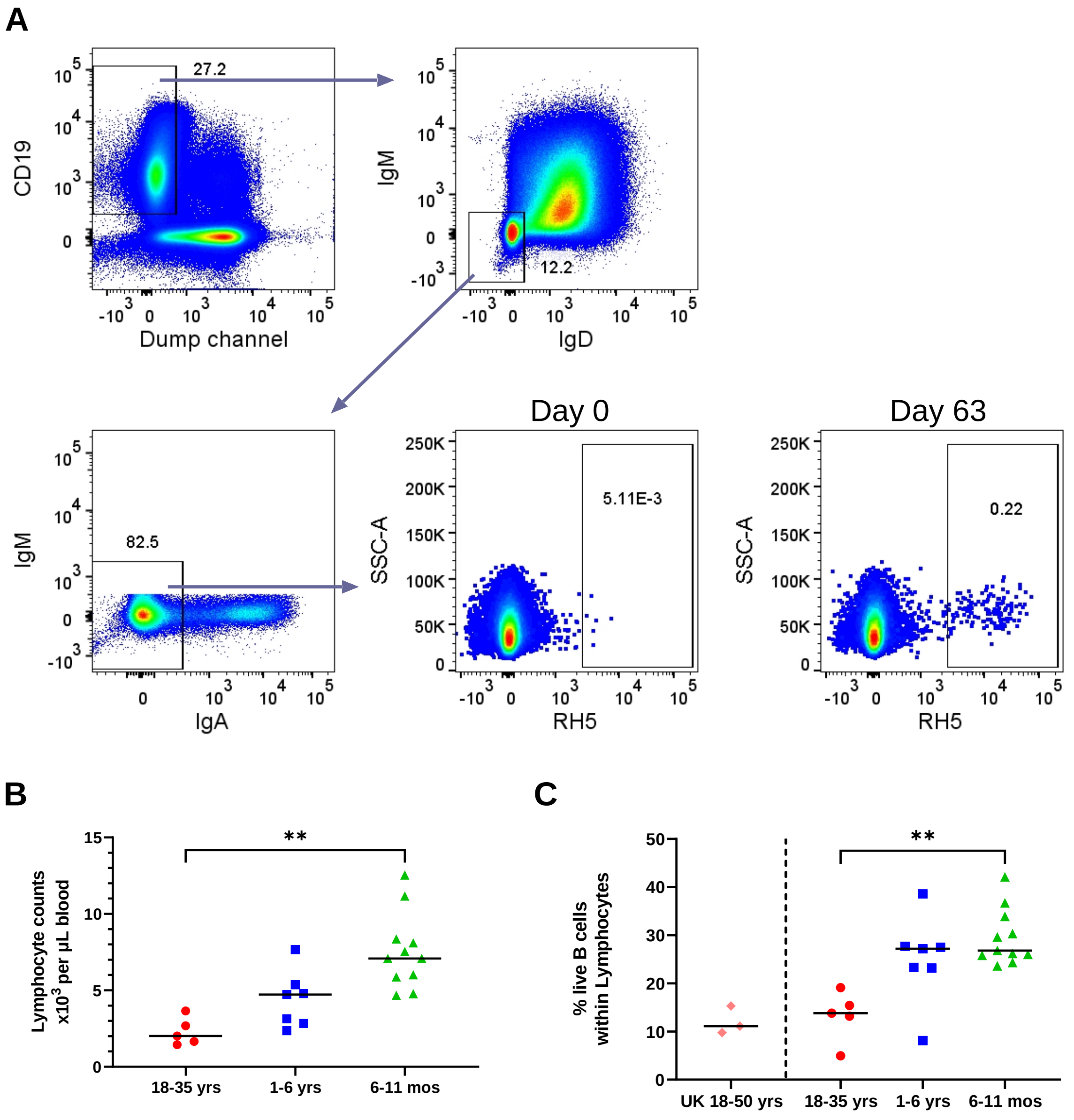

**Supplementary Figure 5. Analyses of RH5-specific B cells.**

(**A**) Gating strategy for identification of RH5-specific cells within the live CD19$^+$ IgD$^-$ IgM$^-$ IgA$^-$ B cell population. Representative RH5 probe staining is shown for matched Day 0 and Day 63 samples from a single vaccinee. Dump channel includes: anti-human CD3, anti-human CD14, anti-human CD56, and a cell viability stain. See Methods for further details. (**B**) Lymphocyte counts per microliter of blood for samples analyzed by flow cytometry. Group 1 adults 18-35 years, N=5; Group 2B children 1-6 years, N=7; Group 3B infants 6-11 months, N=11. (**C**) Percentage of live CD19$^+$ B cells within lymphocytes as assessed by flow cytometry. Same individuals as shown in (**B**). Data from baseline samples for three healthy UK adult volunteers 18-50 years of age enrolled in a previous trial of the ChAd63-MVA RH5 vaccine [1] are shown for comparison only. Individual and median results are shown in each panel. Analyses used Kruskal-Wallis test with Dunn's multiple comparison test across the three vaccinated groups; $^{**}P < 0.01$.

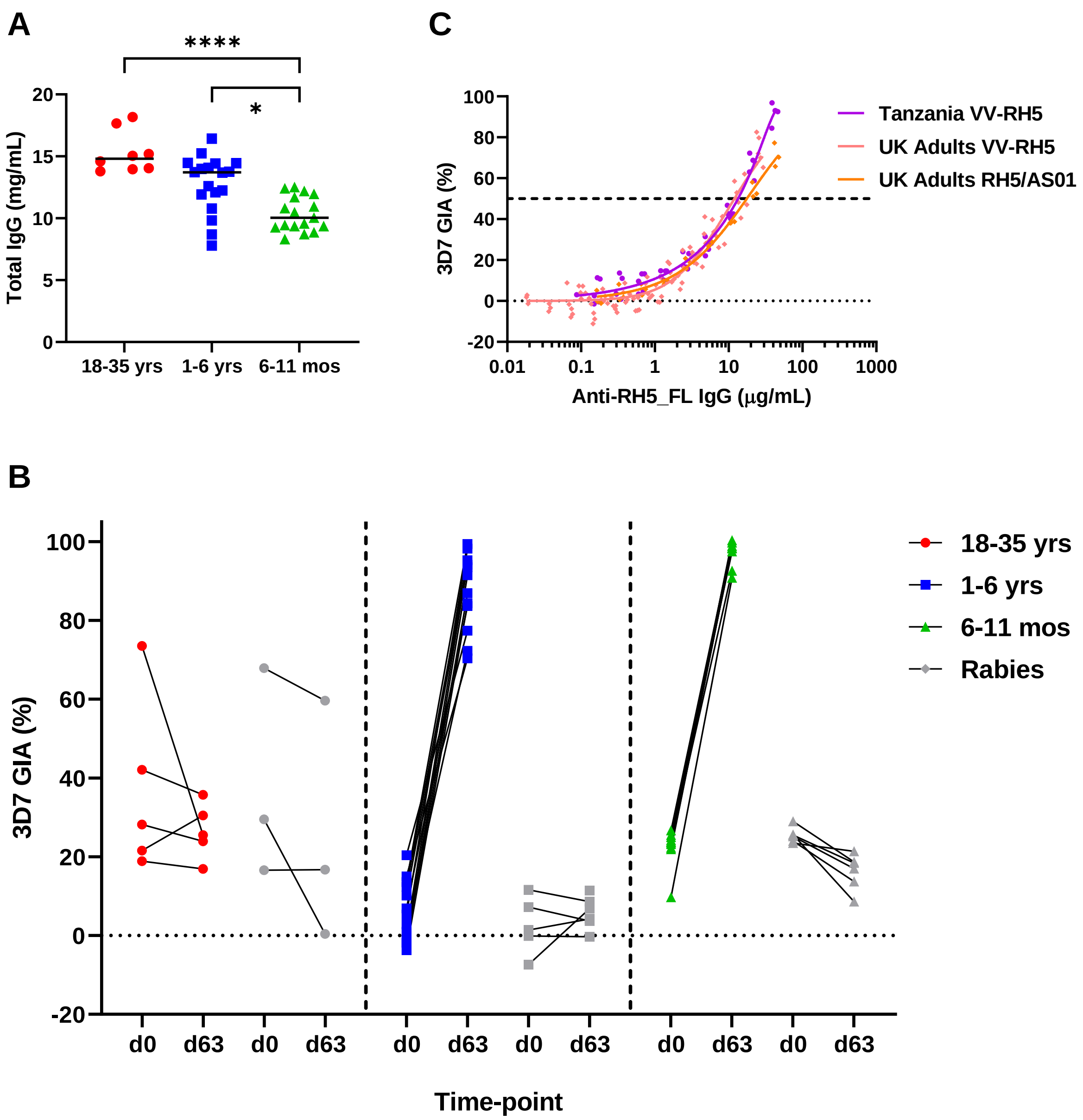

##### Supplementary Figure 6. Serum IgG concentration and GIA assay data.

(**A**) Total serum IgG concentrations in mg/mL at the day 63 time-point for the vaccinated volunteers in the VAC070 trial as measured by HPLC. Median and individual results are shown for all volunteers (RH5 vaccine and rabies vaccine control): Group 1 adults 18-35 years (red), N=8; Group 2B children 1-6 yrs (blue), N=18; and Group 3B infants 6-11 months (green), N= 17. (**B**) *In vitro* growth inhibition activity (GIA) of purified IgG was assessed against 3D7 clone *P. falciparum* parasites. Total IgG purified from serum at i) baseline (day 0) pre-vaccination, and ii) day 63, one week post-MVA RH5 boost vaccination, was initially tested at each sample's physiological total serum IgG concentration as measured in (**A**). Individual linked data shown for Group 1 adults 18-35 years, N=5 vaccinees and N=3 control; Group 2B children 1-6 years, N=12 vaccinees and N=5 or 6 control (one baseline sample was lost during analysis); and Group 3B infants 6-11 months, N=11 vaccinees and N=6 control. (**C**) Samples from three different RH5 vaccine clinical trials were compared for anti-RH5 IgG "functional quality" in an independent head-to-head experiment in the same GIA assay. Samples included those from i) the Tanzanian VAC070 trial (only children and infants) of the viral-vectored ChAd63-MVA RH5 (VV-RH5) vaccine reported here; ii) the VAC057 trial of the VV-RH5 vaccine tested previously in healthy UK adults [1]; and iii) the VAC063 trial of the RH5.1/$AS01_B$ protein-in-adjuvant vaccine tested previously in healthy UK adults [2]. Purified total IgG samples were i) titrated in the GIA assay and ii) tested by ELISA to quantify anti-RH5_FL IgG per mg total IgG. The relationship between the measured GIA and concentration of anti-RH5_FL IgG used in the assay are plotted. Non-linear regression curves are shown for all samples combined from VAC070 (purple line, $r^2$ = 0.97, N=45); VAC057 (pink line, $r^2$ = 0.92, N=90); and VAC063 (orange line, $r^2$ = 0.98, N=27). Dashed line indicates the $EC_{50}$ (concentration of anti-RH5_FL polyclonal IgG that gives 50% GIA).

#### Supplementary Tables

| Age Group | Demographic | ChAd63 RH5 / MVA RH5 | Rabies | Total |
|---|---|---|---|---|
| 6-11 mos | N = | 18 | 9 | 27 |
| | Age (mos) | 9.6 (5.9-11.9) | 9.1 (8.0-10.2) | 9.4 (5.9-11.9) |
| | Male | 10 (55.6%) | 4 (44.4%) | 14 (51.9%) |
| | Female | 8 (44.4%) | 5 (55.6%) | 13 (48.1%) |
| | Weight (kg) | 8.6 (7.0-10.5) | 8.6 (7.0-10.0) | 8.6 (7.0-10.5) |
| | Height (cm) | 71.1 (59.0-81.1) | 69.6 (66.0-72.2) | 70.6 (59.0-81.1) |
| | BMI ($kg/m^2$) | 17.2 (13.7-20.8) | 17.6 (14.5-20.4) | 17.3 (13.7-20.8) |
| | MUAC (cm) | 14.6 (12.0-16.6) | 14.4 (13.4-16.0) | 14.5 (12-16.6) |
| 1-6 yrs | N = | 18 | 9 | 27 |
| | Age (yrs) | 3.4 (1.2-5.6) | 4.0 (2.0-5.6) | 3.6 (1.2-5.6) |
| | Male | 8 (44.4%) | 3 (33.3%) | 11 (40.7%) |
| | Female | 10 (55.6%) | 6 (66.7%) | 16 (59.3%) |
| | Weight (kg) | 13.0 (9.0-17.0) | 13.7 (10.0-17.6) | 13.3 (9.0-17.6) |
| | Height (cm) | 94.6 (75.9-107.0) | 98.8 (83.6-111.0) | 96.0 (75.9-111.0) |
| | BMI ($kg/m^2$) | 14.5 (12.5-16.5) | 14.0 (12.7-15.9) | 14.4 (12.5-16.5) |
| | MUAC (cm) | 15.4 (13.5-17.2) | 15.2 (14.0-16.7) | 15.3 (13.5-17.2) |
| 18-35 yrs | N = | 6 | 3 | 9 |
| | Age (yrs) | 24.8 (19.0-30.2) | 27.7 (24.6-30.7) | 25.8 (19.0-30.7) |
| | Male | 5 (83.3%) | 2 (66.7%) | 7 (77.8%) |
| | Female | 1 (16.7%) | 1 (33.3%) | 2 (22.2%) |
| | Weight (kg) | 56.7 (52.0-60.0) | 59.0 (58.0-60.0) | 57.4 (52.0-60.0) |
| | Height (cm) | 164.9 (152.0-173.5) | 166.3 (159.0-176.0) | 165.4 (152.0-176.0) |
| | BMI ($kg/m^2$) | 21.0 (18.0-24.2) | 21.4 (19.4-23.3) | 21.1 (18.0-24.2) |

##### Supplementary Table 1. Demographics of VAC070 trial participants.

Baseline characteristics are presented as a frequency (%) or mean value with range. BMI = body mass index; MUAC = mid-upper arm circumference.

| Age Group | MedDRA system organ class | MedDRA preferred term | ChAd63-MVA RH5 | Rabies | Total |
|---|---|---|---|---|---|
| **6-11 mos** | Gastrointestinal disorders | Diarrhoea | 0 | 3 | 3 |
| | Respiratory, thoracic and mediastinal disorders | Viral upper respiratory tract infection | 7 | 4 | 11 |
| | Ear and labyrinth disorders | Otitis media | 1 | 0 | 1 |
| **1-6 yrs** | General disorders and administrative site conditions | Pyrexia | 1 | 0 | 1 |
| | Respiratory, thoracic and mediastinal disorders | Viral upper respiratory tract infection | 1 | 2 | 3 |
| | Skin and subcutaneous tissue disorders | Folliculitis | 0 | 1 | 1 |
| | Infections and infestations | Cutaneous larva migrans | 0 | 1 | 1 |
| **18-35 yrs** | Respiratory, thoracic and mediastinal disorders | Viral upper respiratory tract infection | 1 | 0 | 1 |
| | | Tonsillitis | 0 | 1 | 1 |
| | Nervous system disorders | Headache | 1 | 0 | 1 |

**Supplementary Table 2. Unsolicited AEs following vaccination with ChAd63-MVARH5, or rabies control vaccine.**

Unsolicited AEs were collected for 28 days after each dose of vaccine and are presented as frequency. None of the reported unsolicited AEs were assessed as possibly, probably or definitely related to vaccination.

| Lab Parameters | ChAd63-MVA RH5 | | | | Rabies | | | | Total | | | |
|---|---|---|---|---|---|---|---|---|---|---|---|---|
| | Number of abnormalities | Grade | | | Number of abnormalities | Grade | | | Number of abnormalities | Grade | | |
| | | 1 | 2 | 3 | | 1 | 2 | 3 | | 1 | 2 | 3 |
| **Infants 6-11 mos** | | | | | | | | | | | | |
| Increased WBC | **8** | 1 | 3 | 4 | **1** | 0 | 1 | 0 | **9** | 1 | 4 | 4 |
| Decreased WBC | **1** | 1 | 0 | 0 | **0** | 0 | 0 | 0 | **1** | 1 | 0 | 0 |
| Increased Lymphocytes | **0** | 0 | 0 | 0 | **0** | 0 | 0 | 0 | **0** | 0 | 0 | 0 |
| Decreased Lymphocytes | **0** | 0 | 0 | 0 | **0** | 0 | 0 | 0 | **0** | 0 | 0 | 0 |
| Decreased Haemoglobin | **7** | 6 | 1 | 0 | **2** | 2 | 0 | 0 | **9** | 8 | 1 | 0 |
| Decreased Platelets | **1** | 0 | 1 | 0 | **0** | 0 | 0 | 0 | **1** | 0 | 1 | 0 |
| Increased Neutrophil | **0** | 0 | 0 | 0 | **0** | 0 | 0 | 0 | **0** | 0 | 0 | 0 |
| Decreased Neutrophil | **0** | 0 | 0 | 0 | **0** | 0 | 0 | 0 | **0** | 0 | 0 | 0 |
| Increased Eosinophil | **0** | 0 | 0 | 0 | **0** | 0 | 0 | 0 | **0** | 0 | 0 | 0 |
| Increased Creatinine | **1** | 1 | 0 | 0 | **0** | 0 | 0 | 0 | **0** | 0 | 0 | 0 |
| Increased ALT | **3** | 3 | 0 | 0 | **1** | 1 | 0 | 0 | **4** | 4 | 0 | 0 |
| **Children 1-6 yrs** | | | | | | | | | | | | |
| Increased WBC | **1** | 1 | 0 | 0 | **1** | 1 | 0 | 0 | **2** | 2 | 0 | 0 |
| Decreased WBC | **1** | 1 | 0 | 0 | **1** | 1 | 0 | 0 | **2** | 2 | 0 | 0 |
| Increased Lymphocytes | **0** | 0 | 0 | 0 | **0** | 0 | 0 | 0 | **0** | 0 | 0 | 0 |
| Decreased Lymphocytes | **2** | 2 | 0 | 0 | **1** | 1 | 0 | 0 | **3** | 3 | 0 | 0 |
| Decreased Haemoglobin | **1** | 0 | 1 | 0 | **1** | 1 | 0 | 0 | **2** | 1 | 1 | 0 |
| Decreased Platelets | **0** | 0 | 0 | 0 | **0** | 0 | 0 | 0 | **0** | 0 | 0 | 0 |
| Increased Neutrophil | **0** | 0 | 0 | 0 | **1** | 1 | 0 | 0 | **1** | 1 | 0 | 0 |
| Decreased Neutrophil | **0** | 0 | 0 | 0 | **0** | 0 | 0 | 0 | **0** | 0 | 0 | 0 |
| Increased Eosinophil | **1** | 1 | 0 | 0 | **1** | 1 | 0 | 0 | **2** | 2 | 0 | 0 |
| Increased Creatinine | **0** | 0 | 0 | 0 | **0** | 0 | 0 | 0 | **0** | 0 | 0 | 0 |
| Increased ALT | **2** | 2 | 0 | 0 | **0** | 0 | 0 | 0 | **2** | 2 | 0 | 0 |
| **Adults 18-35 yrs** | | | | | | | | | | | | |
| Increased WBC | **0** | 0 | 0 | 0 | **1** | 1 | 0 | 0 | **1** | 1 | 0 | 0 |
| Decreased WBC | **2** | 2 | 0 | 0 | **0** | 0 | 0 | 0 | **2** | 2 | 0 | 0 |
| Increased Lymphocytes | **0** | 0 | 0 | 0 | **0** | 0 | 0 | 0 | **0** | 0 | 0 | 0 |
| Decreased Lymphocytes | **1** | 1 | 0 | 0 | **1** | 1 | 0 | 0 | **2** | 2 | 0 | 0 |
| Decreased Haemoglobin | **0** | 0 | 0 | 0 | **0** | 0 | 0 | 0 | **0** | 0 | 0 | 0 |
| Decreased Platelets | **0** | 0 | 0 | 0 | **1** | 1 | 0 | 0 | **1** | 1 | 0 | 0 |
| Increased Neutrophil | **0** | 0 | 0 | 0 | **0** | 0 | 0 | 0 | **0** | 0 | 0 | 0 |
| Decreased Neutrophil | **0** | 0 | 0 | 0 | **0** | 0 | 0 | 0 | **0** | 0 | 0 | 0 |
| Increased Eosinophil | **0** | 0 | 0 | 0 | **0** | 0 | 0 | 0 | **0** | 0 | 0 | 0 |
| Increased Creatinine | **0** | 0 | 0 | 0 | **1** | 1 | 0 | 0 | **1** | 1 | 0 | 0 |
| Increased ALT | **0** | 0 | 0 | 0 | **0** | 0 | 0 | 0 | **0** | 0 | 0 | 0 |

**Supplementary Table 3. Laboratory AEs related to vaccination with ChAd63-MVA RH5 or rabies vaccine control.**

Table shows laboratory AEs recorded within 28 days of each vaccine administration and considered possibly, probably or definitely related to vaccination. Total numbers vaccinated with ChAd63-MVA RH5: Group 1 adults 18-35 years, N=6; Group 2 children 1-6 years, N=18; Group 3 infants 6-11 months, N=18. Total numbers vaccinated with rabies control: Group 1 adults 18-35 years, N=3; Group 2 children 1-6 years, N=9; Group 3 infants 6-11 months, N=9.

WBC = white blood cells (leukocytes); ALT = alanine aminotransferase.

| Name | Sequence | Pool | | |
|---|---|---|---|---|
| RH5 1 | ENAIKKTKNQENNLALLPIK | P1 | Disordered N-terminus | |
| RH5 2 | ENNLALLPIKSTEEEKDDIK | P1 | | |
| RH5 3 | STEEEKDDIKNGKDIKKEID | P1 | | |
| RH5 4 | NGKDIKKEIDNDKENIKTNN | P1 | | |
| RH5 5 | NDKENIKTNNAKDHSTYIKS | P1 | | |
| RH5 6 | AKDHSTYIKSYLNTNVNDGL | P1 | | |
| RH5 7 | YLNTNVNDGLKYLFIPSHNS | P1 | | |
| RH5 8 | KYLFIPSHNSFIKKYSVFNQ | P1 | | |
| RH5 9 | FIKKYSVFNQINDGMLLNEK | P1 | | |
| RH5 10 | INDGMLLNEKNDVKNNEDYK | P1 | | |
| RH5 11 | NDVKNNEDYKNVDYKNVNFL | P1 | | |
| RH5 12 | NVDYKNVNFLQYHFKELSNY | P1 | | |
| RH5 13 | QYHFKELSNYNIANSIDILQ | P1 | | |
| RH5 14 | NIANSIDILQEKEGHLDFVI | P2 | Start of 45 kDa Fragment | |
| RH5 15 | EKEGHLDFVIIPHYTFLDYY | P2 | | |
| RH5 16 | IPHYTFLDYYKHLSYNSIYH | P2 | | |
| RH5 17 | KHLSYNSIYHKSSTYGKCIA | P2 | | |
| RH5 18 | KSSTYGKCIAVDAFIKKINE | P2 | | |
| RH5 19 | VDAFIKKINEAYDKVKSKCN | P2 | | |
| RH5 20 | AYDKVKSKCNDIKNDLIATI | P2 | | |
| RH5 21 | DIKNDLIATIKKLEHPYDIN | P2 | | |
| RH5 22 | KKLEHPYDINNKNDDSYRYD | P3 | Disordered Loop | |
| RH5 23 | NKNDDSYRYDISEEIDDKSE | P3 | | |
| RH5 24 | ISEEIDDKSEETDDETEEVE | P3 | | |
| RH5 25 | ETDDETEEVEDSIQDTDSNH | P3 | | |
| RH5 26 | DSIQDTDSNHAPSNKKKNDL | P3 | | |
| RH5 27 | APSNKKKNDLMNRAFKKMMD | P3 | | |
| RH5 28 | MNRAFKKMMDEYNTKKKKLI | P4 | Ordered Region 1 | |
| RH5 29 | EYNTKKKKLIKCIKNHENDF | P4 | | |
| RH5 30 | KCIKNHENDFNKICMDMKNY | P4 | | |
| RH5 31 | NKICMDMKNYGTNLFEQLSC | P4 | | |
| RH5 32 | GTNLFEQLSCYNNNFCNTNG | P4 | | |
| RH5 33 | YNNNFCNTNGIRYHYDEYIH | P4 | | |
| RH5 34 | IRYHYDEYIHKLILSVKSKN | P4 | | |
| RH5 35 | KLILSVKSKNLNKDLSDMTN | P4 | | |
| RH5 36 | LNKDLSDMTNILQQSELLLT | P5 | Ordered Region 2 | |
| RH5 37 | ILQQSELLLTNLNKKMGSYI | P5 | | |
| RH5 38 | NLNKKMGSYIYIDTIKFIHK | P5 | | |
| RH5 39 | YIDTIKFIHKEMKHIFNRIE | P5 | | |
| RH5 40 | EMKHIFNRIEYHTKIINDKT | P5 | | |
| RH5 41 | YHTKIINDKTKIIQDKIKLN | P5 | | |
| RH5 42 | KIIQDKIKLNIWRTFQKDEL | P5 | | |
| RH5 43 | IWRTFQKDELLKRILDMSNE | P5 | | |
| RH5 44 | LKRILDMSNEYSLFITSDHL | P6 | Ordered Region 3 | |
| RH5 45 | YSLFITSDHLRQMLYNTFYS | P6 | | |
| RH5 46 | RQMLYNTFYSKEKHLNNIFH | P6 | | |
| RH5 47 | KEKHLNNIFHHLIYVLQMKF | P6 | | |
| RH5 48 | HLIYVLQMKFNDVPIKMEYF | P6 | | Disordered C-terminus |
| RH5 49 | NDVPIKMEYFQTYKKNKPLT | P6 | | |
| RH5 50 | QTYKKNKPLTQ | P6 | | |

#### Supplementary Table 4. RH5 overlapping peptides for ELISPOT assays.

20mer peptides overlapping by 10 aa were used to measure T cell responses by *ex-vivo* IFN-γ ELISPOT assay against the whole of the RH5 insert present in the ChAd63 and MVA vaccines (except for the final peptide RH5-50 which is an 11mer overlapping by 10aa). Peptide sequences are shown, and peptide pools (P) 1-6 used in the ELISPOT assay indicated. These pools corresponded to regions of interest identified from RH5 structural and biochemical analysis [3,4]. Summed total responses across all the RH5 peptide pools in the ELISPOT assay are reported. Two potential or predicted sites of N-linked glycosylation (at sequons N-X-S/T) were mutated Asn (N) to Gln (Q) in the viral vaccine insert (at positions N38 and N214; green letters) [1]; whereas, in a protein version of this vaccine, called RH5.1, all four possible sequons (starting at N38, N214, N284 and N297) were mutated Thr (T) to Ala (A) (green, blue and red letters) [5]. The peptides used in this study corresponded to the RH5.1 protein vaccine sequence, and thus had small mismatches to the RH5 vaccine insert used in the ChAd63 and MVA viral vaccines. This could have potentially led to small under-estimations of the vaccine-induced T cell response if these mutations occurred in critical residues of T cell epitopes.

**Supplementary Appendix – VAC070 Protocol**

**Trial Title:** A Phase Ib age de-escalation dose-escalation randomised, double-blind, controlled study of the safety and immunogenicity of heterologous prime-boost with the candidate malaria vaccines ChAd63 RH5 and MVA RH5 administered intramuscularly according to a 0, 2-month vaccination schedule in healthy adults, young children and infants in Tanzania.

**Short title:** Safety and Immunogenicity of heterologous prime-boost with the candidate malaria vaccines ChAd63 RH5 and MVA RH5 in adults, young children and infants living in Tanzania.

| | |
|---|---|
| Study reference: | VAC070 |
| Protocol Version: | 1.2 |
| Date: | 23rd October 2017 |
| OXTREC Number | 29-17 |
| Sponsor: | University of Oxford |
| Funding body | MRC UK |
| Authors | Ally Olotu, Angela M Minassian, Simon J Draper |

#### Statement of Compliance

I have read this protocol, and I agree to abide by all provisions set forth herein. I agree to comply with the principles of the International Conference on Harmonization Tripartite Guideline on Good Clinical Practice (GCP).

#### Confidentiality Statement

This document contains confidential information that must not be disclosed to anyone other than the trial Sponsor, the Investigator Team, and members of the Ethics Committees and Regulatory Authorities. This information cannot be used for any purpose other than the evaluation or conduct of the clinical investigation without the prior written consent of the principal investigator.

#### Signature Page

| Role | Name | Signature | Date |
|---|---|---|---|
| Principal Investigator | Dr Ally Olotu | | |
| UK Senior Laboratory Investigator | Prof Simon J Draper | | |
| Chief Investigator | Dr Angela M Minassian | | |

### 1. TABLE OF CONTENTS

2. Figures ... 7
3. Tables ... 8
4. KEY ROLES AND GENERAL INFORMATION ... 9
5. SYNOPSIS ... 12
6. ABBREVIATIONS ... 14
7. BACKGROUND AND RATIONALE ... 16
7.1. Malaria epidemiology ... 16
7.2. Malaria vaccines ... 16
7.3. Blood-stage malaria vaccines ... 17
7.4. *Plasmodium falciparum* reticulocyte-binding protein homolog 5 ... 18
7.5. Use of viral vectors in a malaria vaccine platform ... 19
7.6. RH5 candidate malaria vaccine; key safety and immunogenicity data ... 20
Pre-clinical data ... 20
VAC057: Safety and immunogenicity of ChAd63 MVA RH5 in malaria naïve adults ... 21
Safety data ... 21
Humoral immunity data ... 22
Cellular immunity data ... 25
7.7. Experience with virus vectors used with RH5 ... 26
7.8. Rationale ... 26
7.9. Rationale for the use of Rabies vaccine as a control vaccine ... 27
8. OBJECTIVES AND OUTCOME MEASURES ... 28
8.1. Objectives ... 28
8.2. Outcome Measures ... 28
8.3. Time point(s) of evaluation ... 28
9. TRIAL DESIGN ... 29
9.1. Summary of trial design ... 29
9.2. Trial Centre ... 29
9.3. Study Population and Malaria Epidemiology ... 30
9.4. HIV Prevalence ... 31
9.5. Vaccination Services ... 31
9.6. Rationale for Trial Design ... 32
Administration Schedules ... 32
Route & Dose ... 32

16.7. Study results feedback ... 75
17. ETHICAL AND REGULATORY CONSIDERATIONS ... 76
17.1. Declaration of Helsinki ... 76
17.2. Guidelines for Good Clinical Practice ... 76
17.3. Approvals ... 76
17.4. Reporting ... 76
17.5. Participant Confidentiality ... 76
17.6. Potential Benefits to participants ... 77
17.7. Potential Risks and Burden to Participants ... 77
Vaccination: ... 77
Phlebotomy: ... 78
17.8. Incentives ... 78
17.9. Future use of stored samples ... 78
18. FINANCE AND INSURANCE ... 80
18.1. Funding ... 80
18.2. Insurance ... 80
19. PUBLICATION POLICY ... 81
20. REFERENCES ... 82
21. APPENDIX A: SCHEDULE OF PROCEDURES ... 90
22. APPENDIX B: SAE REPORTING FLOW CHART ... 93
23. APPENDIX C: AMENDMENT HISTORY ... 94

#### 2. Figures

#### 3. Tables

#### 4. KEY ROLES AND GENERAL INFORMATION

| | |
|---|---|
| **Trial Centre** | Ifakara Health Institute Clinical Trial Facility,<br>Bagamoyo Research and Training Centre<br>P.O. Box 74<br>Bagamoyo, Tanzania |
| **Principal Investigator (IHI):** | Ally Olotu MD DPhil<br>Ifakara Health Institute<br>P.O. Box 74<br>Bagamoyo, Tanzania<br>Tel: +255 718 927 104<br> |
| **UK Senior Laboratory Investigator (UOXF):** | Prof Simon J Draper<br>Jenner Institute<br>University of Oxford<br>Old Road Campus Research Building<br>Roosevelt Drive<br>Headington Oxford, OX3 7DQ<br>United Kingdom. |
| **Co-investigators:** | *Ifakara Health Institute, Bagamoyo, Tanzania*<br>1. Salim Abdulla MD PhD,<br>2. Nahya Salim MD, MMED, PhD<br>3. Said Jongo, MD, MMED<br>4. Omary Lweno MD, Msc<br>5. Maximillian Mpina, PhD |
| **Chief Investigator UOXF** | Dr Angela M Minassian<br>Jenner Institute<br>Centre for Clinical Vaccinology & Tropical Medicine<br>University of Oxford<br>Churchill Hospital<br>Old Road<br>Oxford, OX3 7LJ<br>United Kingdom |
| **Project Manager (Tanzania)** | *Ifakara Health Institute*<br>Saumu Ahmed MD, PGDip,<br>Tel: 255 784 358 670<br> |
| **Sponsor:** | University of Oxford<br>Named Contact: Dr Rebecca Bryant |

| | |
|---|---|
| | Research Services<br>University Offices<br>Wellington Square<br>Oxford, OX1 2JD<br>United Kingdom<br>Phone number: +44 (0)1865 282585<br> |
| **Funder:** | Medical Research Council<br>14th floor<br>One Kemble Street<br>London<br>WC2B 4AN<br>United Kingdom |
| **Local Safety Monitor** | Prof Karim Manji, M.D., M.Med., M.P.H.<br>Department of Pediatrics and Child Health<br>Muhimbili University of Health and Allied Sciences<br>Dar es Salaam, Tanzania<br> |
| **Independent Statistician** | Ali Ali, Msc PhD<br>Ifakara Health Institute, Tanzania |
| **Ethics Committees** | Ifakara Health Institute IRB<br>P.O. Box 78373<br>Dar es Salaam, Tanzania<br>Tel: +255 (0) 22 2774714,<br>Fax: + 255 (0) 22 2771714<br><br><br>National Health Research Ethics Sub-Committee (NatHREC)<br>National Institute for Medical Research<br>P.O. Box 9653<br>Dar es Salaam, Tanzania<br>Tel: +255 22 2121400<br>Fax: 255 22 2121360<br><br><br>Oxford Tropical Research Ethics Committee (OxTREC)<br>University of Oxford<br>Research Services |

| | |
|---|---|
| | University Offices<br>Wellington Square<br>Oxford, OX1 2JD<br>United Kingdom<br>Tel: +44 (0)1865 282585<br> |
| **Regulatory Authority** | Tanzania Food and Drug Authority<br>P.O. Box: 77150<br>Dar es Salaam, Tanzania.<br>Tel: +255 22 2450512 / 2450751 / 2452108<br>Fax: +255 22 2450793<br> |
| **Laboratories** | 1. Bagamoyo Research and Training Centre Laboratory, Ifakara Health Institute, Bagamoyo Tanzania.<br>2. Jenner Institute laboratories, Centre for Clinical Vaccinology & Tropical Medicine, University of Oxford, Churchill Hospital, Old Road, Oxford, OX3 7LJ, United Kingdom.<br>3. NIH/NIAID Laboratory of Malaria and Vector Research, Malaria Immunology Section, GIA Reference Centre, 12735 Twinbrook Parkway, Twinbrook III, Room 3W-13, Rockville, MD 20852, USA. |

#### 5. SYNOPSIS

<table>
<tr><td>Trial Title</td><td colspan="2">A Phase Ib age de-escalation dose-escalation randomised, double-blind, controlled study of the safety and immunogenicity of heterologous prime-boost with the candidate malaria vaccines ChAd63-RH5 and MVA-RH5, administered intramuscularly according to a 0, 2-month vaccination schedule, in healthy adults, young children and infants in Tanzania.</td></tr>
<tr><td>Study reference</td><td colspan="2">VAC070</td></tr>
<tr><td>Clinical Phase</td><td colspan="2">Phase Ib</td></tr>
<tr><td>Trial Design</td><td colspan="2">Age de-escalation dose-escalation randomised double-blind controlled trial. The study will consist of 5 groups as shown below.<br>
<table>
<tr><th>Groups</th><th>Age at enrollment</th><th># participants</th><th>Vaccination at day 0</th><th>Vaccination at day 56</th></tr>
<tr><td rowspan="2">Group 1</td><td rowspan="2">18-35 years</td><td>6</td><td>$5x10^{10}$vp ChAd63 RH5</td><td>$2x10^{8}$pfu MVA RH5</td></tr>
<tr><td>3</td><td>Rabies</td><td>Rabies</td></tr>
<tr><td rowspan="2">Group 2a</td><td rowspan="2">1-6 years</td><td>6</td><td>$1x10^{10}$vp ChAd63 RH5</td><td>$1x10^{8}$pfu MVA RH5</td></tr>
<tr><td>3</td><td>Rabies</td><td>Rabies</td></tr>
<tr><td rowspan="2">Group 2b</td><td rowspan="2">1-6 years</td><td>12</td><td>$5x10^{10}$vp ChAd63 RH5</td><td>$2x10^{8}$pfu MVA RH5</td></tr>
<tr><td>6</td><td>Rabies</td><td>Rabies</td></tr>
<tr><td rowspan="2">Group 3a</td><td rowspan="2">6-11 months</td><td>6</td><td>$1x10^{10}$vp ChAd63 RH5</td><td>$1x10^{8}$pfu MVA RH5</td></tr>
<tr><td>3</td><td>Rabies</td><td>Rabies</td></tr>
<tr><td rowspan="2">Group 3b</td><td rowspan="2">6-11 months</td><td>12</td><td>$5x10^{10}$vp ChAd63 RH5</td><td>$2x10^{8}$ pfu MVA RH5</td></tr>
<tr><td>6</td><td>Rabies</td><td>Rabies</td></tr>
</table>
</td></tr>
<tr><td>Study population</td><td colspan="2">Healthy adults (18-35 years), young children (1-6 years) and infants (6-11 months) residing in Bagamoyo, Tanzania. A total of 63 participants will be enrolled.</td></tr>
<tr><td>Follow up duration</td><td colspan="2">All participants will be followed for 16 weeks after the MVA RH5 vaccination. The duration of the entire study will be 6 months per participant from the time of first vaccination.</td></tr>
<tr><td>Planned Trial Period</td><td colspan="2">1.5 years after the start of recruitment</td></tr>
<tr><td></td><td>Objectives</td><td>Outcome Measures</td></tr>
<tr><td>Primary</td><td>To determine safety and tolerability of ChAd63-RH5 and MVA-RH5 given intramuscularly at a 2 month interval in adults (18-35 years), children (1-6 years) and infants (6-11 months) residing in a malaria endemic country.</td><td>• Solicited symptoms after vaccination.<br>• Unsolicited symptoms after each vaccination.<br>• Serious adverse events during the study period.</td></tr>
</table>

| | | |
|---|---|---|
| Secondary | To evaluate the magnitude of antibody and cellular immune responses to RH5 in adults, children and infants residing in a malaria endemic country.<br>To evaluate the quality of antibody and cellular immune responses to RH5 in adults, children and infants residing in a malaria endemic country.<br>To evaluate the longevity of antibody and cellular immune responses to RH5 in adults, children and infants residing in a malaria endemic country. | • Anti-RH5 antibody concentration by ELISA.<br>• Growth inhibition activity of sera from vaccinees on a panel of *P. falciparum* parasites.<br>• Avidity of anti-RH5 antibodies by ELISA and surface plasmon resonance (SPR) and/or other assays (to be defined). Cellular immune responses to RH5 by ELISpot assay and/or Intracellular Cytokine Staining (ICS) and/or other assays (to be defined). |
| Investigational Medicinal Product(s) | ChAd63 RH5<br>MVA RH5<br>Rabies vaccine | |
| Route of Administration | All vaccines will be given by intramuscular injection to the left deltoid area. | |

#### 6. ABBREVIATIONS

| | |
|---|---|
| ALT | Alanine Aminotransferase |
| ASC | Antibody Secreting Cells |
| BCG | Bacillus Calmette–Guérin |
| BDH | Bagamoyo District Hospital |
| BMI | Body Mass Index |
| CBF | Clinical Biomanufacturing Facility |
| CHW | Community Health Worker |
| CMI | Cell Mediated Immunity |
| CRF | Case Report form |
| CRO | Clinical Research Organization |
| CSP | Circumsporozoite Protein |
| CTA | Clinical Trial Agreement |
| DNA | Deoxyribonucleic acid |
| DPT | Diphtheria, Pertussis and Tetanus. |
| EDC | Electronic Data Capture |
| EPI | Expanded Program of Immunization |
| FVO | Falciparum Vietnam Oak-Knoll |
| GCP | Good Clinical Practice |
| GIA | Growth Inhibition Assay |
| GPI | Glycosylphosphatidylinositol |
| GSK | Glaxosmithkline |
| HBV | Hepatitis B Virus |
| HIV | Human Immunodeficient Virus |
| HRA | Health Research Authority |
| ICF | Informed Consent Form |
| ICS | Informed Consent Sheet |
| IDT | Impfstoffwerke DessauTornau |
| IFN | Interferon |
| IHI | Ifakara Health Institute |
| IPT | Intermittent Presumptive Treatment |
| IRB | Institutional Review Board |
| IRS | Indoor Residual Spraying |
| ISM | Independent Safety Monitor |
| IVD | Immunization and Vaccine Development |
| CTF | Clinical Trial Facility |
| MRC | Medical Research Council |
| MVA | Modified Vaccinia Ankara |

| | |
|---|---|
| NatHREC | National Health Research Ethics Sub-Committee (NatHREC) |
| OPV | Oral Polio Virus |
| OXTREC | Oxford Tropical Research Ethics Committee |
| PBMC | Peripheral blood mononuclear cells |
| PIS | Participant Information Sheet |
| RBC | Red Blood Cells |
| RSV | Respiratory Syncytial Virus |
| SAE | Serious Adverse Event |
| SAR | Serious Adverse Reaction |
| SDV | Source Data Verification |
| SMC | Safety Monitoring Committee |
| SOP | Standard Operating Procedures |
| SPR | Surface Plasmon Resonance |
| TFDA | Tanzania Food and Drug Authority |
| TMF | Trial Master File |
| TSG | Oxford University Hospitals NHS Foundation Trust / University of Oxford Trials Safety Group |
| UOXF | University of Oxford |
| WHO | World Health Organization |

### 7. BACKGROUND AND RATIONALE

#### 7.1. Malaria epidemiology

Malaria still remains a disease of public health significance affecting millions across the globe. In 2015, it was estimated 212 million new cases of clinical malaria were diagnosed worldwide resulting in more than 430,000 malaria deaths [1]. This is likely to be an underestimation since the estimates are highly influenced by the surveillance methods used [2]. The highest malaria burden is in children below the age of five years among whom over 303,000 and 292,000 deaths were reported worldwide and in Africa respectively in 2015. In addition to the immediate morbidity and mortality risk, malaria may inflict long-term effects with negative consequences on health and quality of life. For example, children who suffer from severe malaria are at increased risk of epilepsy [3, 4], chronic neurological and cognitive impairment [5]. Furthermore, malaria causes significant economic hardships and disproportionately affects the poor [6]. In endemic countries, it is estimated that more than 1% loss in gross domestic product is attributable to malaria [7, 8].

Malaria control interventions such as insecticide treated bed-nets (ITNs), treatment with artemisinin based combination therapies (ACTs), intermittent presumptive treatment (IPT) and indoor residual spraying (IRS) have made a significant impact on the burden of malaria. There have been encouraging reports on a decline of malaria from several parts of Africa [9-12] but this has not been consistent everywhere with some areas reporting sustained or even an increase in the burden of malaria [13, 14].

The progress made in controlling malaria is however being threatened by development of resistance in the parasite and in the vector. The above interventions also do not appear sufficient to eliminate malaria in high transmission settings in Africa [15].

#### 7.2. Malaria vaccines

Immunisation against communicable diseases is one of the most cost-effective public health interventions [16, 17]. The global health success of eradicating smallpox was the result of a widespread deployment of smallpox vaccine [18]. Furthermore, significant inroads have been made in the fight against other major childhood infectious diseases such as polio, *Haemophilus influenzae* and measles through immunisation [19, 20]. The success of vaccines is based not only on their ability to offer protection to the individual recipients, but also to the community by reducing the transmission through herd immunity [21, 22].

There have been many strategies utilized in the development of an effective malaria vaccine including development of peptides [23, 24], recombinant proteins and fusion proteins [25-28] targeting specific stages of the *Plasmodium falciparum* (*Pf*) life cycle, non-replicating whole sporozoites [29], DNA vaccines [30] and vectored and prime-boost vaccines [31].

The most clinically advanced malaria vaccine candidate, RTS,S/AS01, has shown modest efficacy in children and infants in Phase II and III field trials [32, 33]. The efficacy wanes over time and there is potential for rebound in areas with high malaria transmission [34], providing strong rationale for the future inclusion of an effective blood-stage component to prevent clinical disease as pre-erythrocytic anti-infection immunity wanes.

Thrombospondin-related adhesion protein (TRAP) fused to a multi-epitope (ME) is another pre-erythrocytic vaccine candidate that has shown promising results and is currently being evaluated in the field [35, 36]. The antigen is delivered using a viral vector prime-boost strategy. Early studies using folwpox strain 9 or plasmid DNA as priming vector followed by MVA were unsuccessful in providing adequate protection in malaria-exposed individuals [37]. Chimpanzee adenovirus serotype 63 (ChAd63) as a priming vector has provided better immunogenicity and significant protection, as has been demonstrated in both malaria naïve and malaria exposed adults [38-40]. The trials to investigate the efficacy of ME-TRAP in children and infants are ongoing.

Whole sporozoite-based approaches provide short term homologous protection in malaria-naïve adults up to 14 months after the last dose [29, 41]. The vaccine induces high levels of tissue resident $CD8^+$ T cell responses and moderate levels of antibodies to Pf circumsporozoite protein (CSP), both of which correlate with protection. In malaria exposed individuals who receive comparable doses to malaria naïve adults, efficacy against malaria infection is moderate both by experimental and natural challenge [42]. Studies are still ongoing to further optimize the dose and schedule including evaluating the potential of non-attenuated whole sporozoites given under anti-malarial chemoprophylaxis in malaria exposed individuals.

Transmission-blocking vaccines would be ideal for elimination, however, most of these vaccine candidates are still in early stages of development [43, 44].

##### 7.3. Blood-stage malaria vaccines

Blood-stage malaria vaccines are essential for prevention of morbidity and mortality from malaria. The importance of blood-stage naturally acquired immune responses was demonstrated by experiments using passive transfer of immunoglobulins [45, 46]. However the naturally acquired immunity develops slowly over time, does not completely prevent infection and wanes if exposure is removed [47]. In 2002 it was shown that blood-stage immunity can be induced experimentally through immunization with ultra-low doses of infected erythrocytes [48], although the effect of antimalarial used to clear parasitaemia in this study could not be completely ruled out as confounding factor.

Most of the research on blood-stage vaccines has focused on the antigens which are critical in the host-parasite interactions and expressed on the merozoites. Although the safety and immunogenicity of many blood-stage candidate vaccines have been promising, they have shown no clinical protection [28, 49] or only strain-specific protection [27] in the field. Substantial levels of polymorphism among the most widely studied blood-stage antigens and redundant erythrocyte invasion pathways present a major challenge for the development of blood-stage vaccines [50]. In addition rapid erythrocyte invasion means extremely high concentrations of functional antibody are required to neutralize the parasite to prevent erythrocyte invasion [51].

Thus while the development of blood-stage malaria vaccines is faced with challenges, they remain a critical element of the malaria vaccine portfolio. With the decline in malaria transmission and subsequent waning of naturally acquired immunity, populations living in a malaria endemic country will become increasingly at risk of a malaria epidemic once the interventions are interrupted [52], making a blood-stage malaria vaccine an essential component of any integrated malaria control programme. Furthermore blood-stage malaria vaccines need to be part of the multi-stage vaccine that would protect against rebound malaria when the immunity to the pre-erythrocytic component of the vaccine wanes over time.

##### 7.4. ***Plasmodium falciparum*** **reticulocyte-binding protein homolog 5**

*Pf* reticulocyte-binding protein homolog 5 (PfRH5) is a new generation and highly promising blood-stage malaria vaccine antigen [53]. The PfRH5 is part of the family of a protein complex consisting of PfRH1, PfRH2a, PfRH2b, PfRH3 and PfRH4. PfRH proteins are located in the apical organelles of the merozoite and are released onto the surface during invasion of erythrocytes. In contrast to other protein members, PfRH5 is a much shorter protein (~60kDa for PfRH5 vs 200-375kDa for other PfRH proteins) and does not have a transmembrane and cytosolic region at the C-terminus [54].

PfRH5 forms a complex with another two antigens, namely PfRH5-interacting protein (PfRipr) [55] and the cysteine-rich protective antigen (PfCyRPA) [56] to facilitate interaction with the erythrocyte. It has recently been shown that the N-terminal region of RH5 (RH5Nt) binds glycosylphosphatidylinositol (GPI)-anchored merozoite protein

P113, providing a platform for RH5 to attach to the merozoite surface [57]. It is hypothesized that CyRPA recruits Ripr to the complex resulting in the release of the RH5-CyRPA-Ripr complex. The RH5-CyRPA-Ripr complex is thought to play a role in the formation of a pore between the merozoite and the RBC that allows $Ca^{2+}$ ingress into the host cell prior to establishment of the tight junction.

PfRH5 binds basigin on the surface of erythrocytes forming a critical non-redundant interaction during erythrocyte invasion [58]. Immunization with full-length PfRH5 immunogens induces functional antibodies to PfRH5, unlike fragments of antigens produced from *E. coli* [54, 59]. Because of the technical challenge of generating full-length PfRH5 protein (RH5_FL), the earliest promising results were achieved using viral vectored immunization [60], whereby antigen is expressed *in situ* from virally infected muscle cells [61]. Recently, production of full-length PfRH5 protein has been successful from numerous heterologous expression systems including mammalian HEK293 cells [62], *E. coli* [63, 64], baculovirus-infected insect cells [65, 66], wheatgerm [67],and *Drosophila* S2 stable cell lines [68].

PfRH5 has several advantages compared to other investigational blood-stage malaria vaccine candidates, namely PfMSP1 and PfAMA1. PfRH5 is highly conserved across parasite lines, thus leading to strain-transcending neutralizing antibodies post-vaccination [60, 69]. Furthermore the high degree of PfRH5 sequence conservation is associated with functional constraints linked to basigin binding. Substitution of minimal amino acids in PfRH5 results in loss of basigin binding, suggesting the antigen may not easily escape vaccine-induced immune pressure [70-72].

Antibodies to PfRH5 can also block invasion of erythrocytes with high potency, quantitatively requiring less antibody levels to achieve maximal blockage compared to other antigens [73]. In addition, antibodies to PfRH5 can cross-inhibit all field isolates and laboratory strains of *Pf* tested to date [58, 60, 69]. Naturally-acquired antibodies to PfRH5 are several fold lower compared to other blood-stage antigens such as PfMSP1 and PfAMA1 indicating that PfRH5 is not a dominant target of naturally acquired immunity [65, 74]. However antibodies to PfRH5 have been shown to inhibit parasite growth *in vitro* and predict protection from malaria in the field [65, 75].

##### 7.5. Use of viral vectors in a malaria vaccine platform

Viral vectors have been evaluated as an antigen delivery platform for vaccines in HIV, Ebola and Tuberculosis [76] This approach uses modified live viruses as vectors that carry antigens or antigen-encoding genes for the antigen of interest leading to the *in vivo* expression of these antigens when subjects are immunized. They have an advantage over traditional vaccine delivery platforms in that they stimulate a broad range of immune responses including both humoral and cell mediated immunity [61, 77]. Viral vectors have been thoroughly investigated in animal models and many of them are now being evaluated in humans.

In the case of RH5, a recombinant replication-deficient adenovirus of simian serotype is used to prime the immune response, followed by a booster vaccination 8 weeks later with an attenuated poxvirus recombinant for the same antigen [61].

These vectors, when used to deliver antigens from *P. falciparum*, have now been shown to be safe and immunogenic for T cells and antibodies in healthy European and American adult volunteers [40, 78-81], as well as African adults, children and infants [82, 83]. More recently, similar adenovirus-poxvirus vectored vaccine technologies have been used to immunize humans against numerous other pathogens including *P. vivax* malaria [84], Ebola virus [85], hepatitis C virus (HCV) [86], respiratory syncytial virus (RSV) [87] and human immunodeficiency virus-1 (HIV-1) [88].

A viral-vector prime-boost strategy has been preferred for recombinant antigens which are difficult to produce using heterologous expression platforms [77, 89, 90]. RH5_FL is one of such antigens, which for years has proved difficult to produce using conventional recombinant technology until a viral vector platform was utilized leading to promising data that led to a clinical trial in humans [60].

##### 7.6. RH5 candidate malaria vaccine; key safety and immunogenicity data

An RH5 candidate malaria vaccine is being developed for the routine immunization of infants and children living in malaria-endemic areas as part of the Expanded Program of Immunization (EPI). In addition it is being developed as an essential component of a multi-stage malaria vaccine aimed to completely block malaria transmission. The clinical development plan for RH5 involves investigation of different approaches to deploy the antigen, including protein-in-adjuvant, viral vectors and novel vaccine adjuvants. To date, an RH5 candidate malaria vaccine has been administered to malaria-naïve adult subjects as well as non-human primates.

*Pre-clinical data*

A pre-clinical study using viral vectors or PfRH5 protein produced from mammalian HEK293 cells showed that PfRH5-based vaccine vaccines can protect *Aotus* monkeys against a virulent vaccine-heterologous *Pf* challenge [89]. Protection was strongly correlated with anti-PfRH5 serum IgG antibody concentration and *in vitro* functional growth inhibition activity (GIA) using purified IgG [89], confirming the utility of this assay to predict *in vivo* protection and for future vaccine candidate down-selection.

#### *VAC057: Safety and immunogenicity of ChAd63 MVA RH5 in malaria naïve adults*

The first administration of RH5 in humans utilized a vectored prime-boost approach and was conducted among malaria naïve adults at the Centre for Clinical Vaccinology and Tropical Medicine at the University of Oxford (UOXF) and the NIHR WTCRF in Southampton.

Twenty four healthy, malaria-naïve males and non-pregnant females aged 18-50 were recruited into the study to assess the safety and immunogenicity of ChAd63 RH5/MVA RH5. Eligible participants were randomized to Groups 1, 2A, 2B and 2C to receive a lead-in dose of ChAd63 RH5 ($5 \times 10^9$ viral particles, vp), or the target dose of ChAd63 RH5 ($5 \times 10^{10}$ vp) alone or followed by MVA RH5 ($1 \times 10^8$ plaque-forming units, pfu) 8 weeks later, or ChAd63 RH5 ($5 \times 10^{10}$ vp ) followed by MVA RH5 ($2 \times 10^8$ pfu ) 8 weeks later in a 1:1:2:2 ratio (Figure 1). Vaccinations were given through intramuscular injection in the deltoid area.

| Group | Number of volunteers | Dose of ChAd63 RH5 | Dose of MVA RH5 |
|---|---|---|---|
| 1 | 4 | $5 \times 10^9$ vp IM | |
| 2A | 4 | $5 \times 10^{10}$ vp IM | |
| 2B | 8 | $5 \times 10^{10}$ vp IM | $1 \times 10^8$ pfu IM |
| 2C | 8 | $5 \times 10^{10}$ vp IM | $2 \times 10^8$ pfu IM |

*Figure 1 VAC057 vaccination groups receiving ChAd63-MVA RH5*

#### Safety data

There were no serious adverse events (SAEs) or unexpected reactions and no safety concerns during the course of the trial. The reactogenicity of the vaccines was similar to that seen in previous malaria vaccine trials using the same viral vectors at similar doses in healthy adults [78, 84, 91], with the higher doses of both vaccines associated with an increased number and higher severity of reported adverse events (AEs) (Figure 2). The majority of AEs following ChAd63 RH5 were mild, but moderate AEs were reported by some participants in both groups, and two participants who received the full dose reported severe AEs on the day of vaccination which resolved within 24 hours. All moderate or severe solicited systemic AEs following MVA RH5 occurred in participants who had received the higher dose of vaccine. The majority of solicited AEs occurred within the first 2 days after vaccination and the median duration of each systemic AE was between 1 and 2 days following either vaccine. There were no severe laboratory AEs following ChAd63/MVA RH5 vaccination. One participant had a moderately raised ALT at day 7 following ChAd63 RH5 which had resolved fully by day 28. One participant had moderate thrombocytopenia and mild leukopenia at day 28 following ChAd63 RH5 but had commenced post-exposure prophylaxis for HIV exposure the day before these bloods were taken, so causality is unclear. All other laboratory AEs were mild and had resolved fully by day 84 except for one participant who had a persistent mild anaemia. This had been present at screening and had not worsened over the course of the study so was considered unrelated to the vaccine.

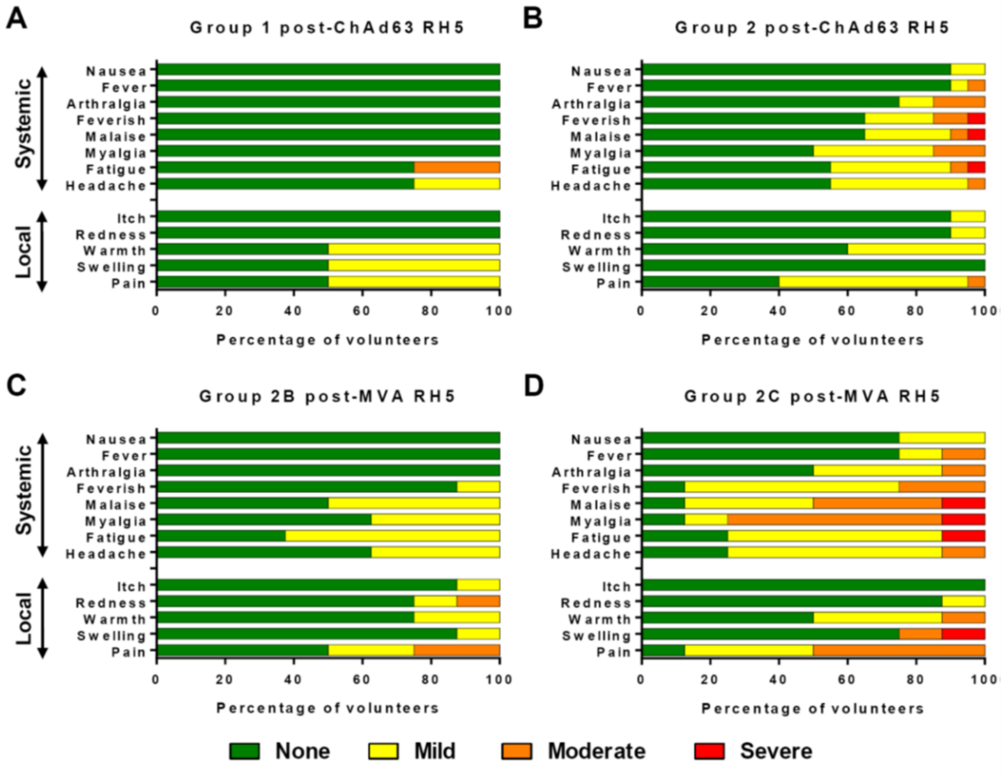

*Figure 2 ChAd63/MVA RH5 safety data in malaria naive adults*

#### *Humoral immunity data*

Priming with 5 x $10^{10}$ vp ChAd63 RH5 followed by MVA RH5 boost induced significant antigen-specific IgG responses in all participants (Groups 2B and 2C) as shown in Figure 33. The vaccine-induced serum antibody responses against RH5_FL appear to be mainly IgG1 with moderate levels of IgG3 and little or no IgG2 or IgG4 subtypes. The antibody responses peaked on day 84 (four weeks after the booster dose of MVA RH5) and later declined over time remaining significantly above the pre-vaccination level on day 140. Anti-RH5 antibody levels post-boost were significantly higher in comparison to those found in individuals exposed to natural malaria (Figure 4).

The avidity of anti-RH5_FL IgG increased over time both in those who did and did not receive the MVA RH5 booster, suggesting IgG affinity maturation over time as opposed to a direct consequence of the MVA RH5 boost.

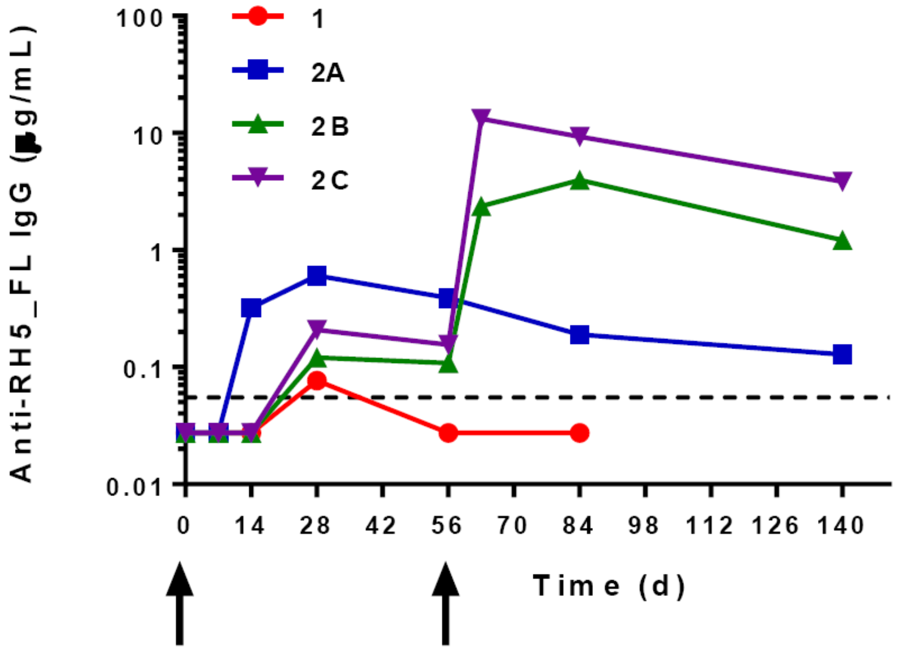

*Figure 3 Anti-RH5 antibodies levels following immunization with ChAd63/MVA RH5*

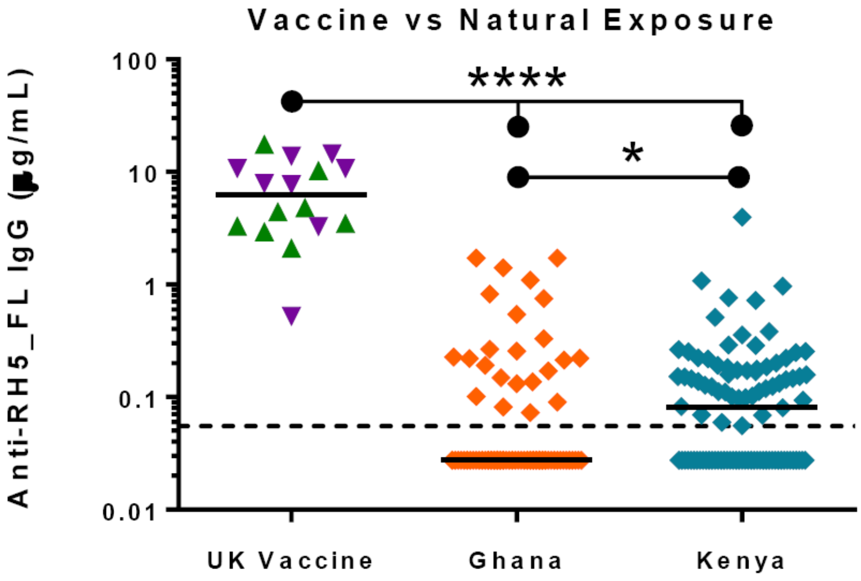

*Figure 4: Comparison between anti-RH5 antibodies in semi-immune African and naïve UK participants*

The sera taken 4 weeks after the MVA RH5 following a primary ChAd63 RH5 dose had a median *in vitro* GIA of 36.0 % (range 19.7-61.6 %) and 50.6 % (range 7.2-68.1 %) using 10 mg/mL purified IgG in those who received lower and higher doses respectively (Figure 5). GIA was inversely correlated with the IgG concentration as previously seen with the merozoite surface protein 1 (MSP1) and apical membrane antigen 1 (AMA1) antigens [92-94] and was not enhanced by inclusion of complement.

The concentration of RH5_FL-specific polyclonal IgG required to give 50% GIA ($EC_{50}$) was only 8.2 µg/mL, substantially lower than AMA1 and MSP1 candidate vaccines [92-94]. Similarly purified IgG from RH5 vaccinees had a higher overall level of GIA against 3D7 clone parasites using 10 mg/mL compared to those induced from vaccination with ChAd63-MVA expressing AMA1 or MSP1 [95]. However, the levels of GIA using 10 and 2.5 mg/mL purified IgG against 3D7 clone parasites were comparable to those reported previously in healthy UK adults immunized with an AMA1 recombinant protein vaccine delivered in the proprietary adjuvant system AS01B from GSK [93]. This could be due to 10-fold higher immunogenicity of AMA1 administered with potent AS01 adjuvant compared to ChAd63-MVA [93].

Purified IgG from ChAd63-MVA RH5 vaccinees were able to neutralize parasites from a panel of eight laboratory-adapted parasite lines (7G8, Dd2, FVO, GB4, MCamp) and short-term culture-adapted parasite isolates (from Cambodian patients with malaria (Cp845, Cp806) [73] or from an Australian resident who contracted malaria in Ghana (HMP002) [96], that between them include RH5 sequences that encompass the five most common RH5 single nucleotide polymorphisms (SNPs)).

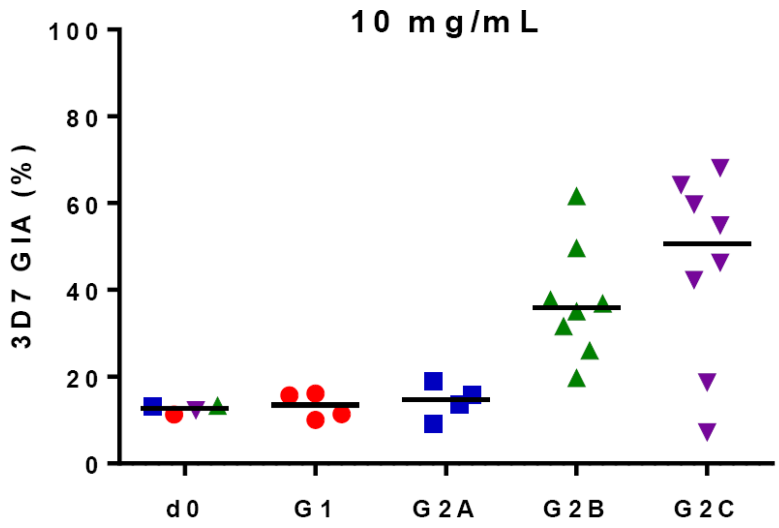

*Figure 5: GIA measured following ChAd63/MVA RH5 vaccination*

#### Cellular immunity data

Vaccination with ChAd63-MVA RH5 induced antigen-specific T cell responses in all participants which were boosted by vaccination with MVA RH5 8-weeks later (Figure 6).

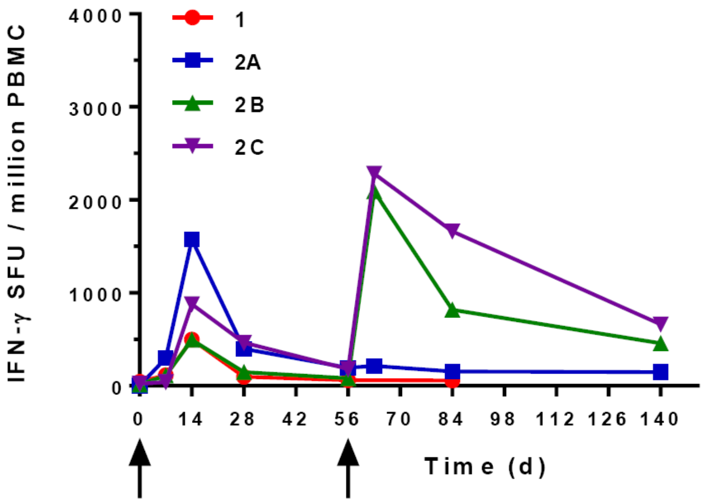

*Figure 6: Kinetics and magnitude of the RH5-specific T cell response by ex-vivo IFN-γ ELISPOT in malaria naïve adults*

#### 7.7. Experience with virus vectors used with RH5

The ChAd63 vector, encoding malaria antigens, has been safety administered to more than 900 (including 139 children and infants in malaria endemic countries) individuals and has demonstrated an excellent safety profile, with doses of up to 2 x $10^{11}$ vp ChAd63 ME-TRAP. MVA encoding various antigens, including several *P. falciparum* malaria antigens, has also been used in over 1000 individuals (including young children and infants), with an excellent safety profile. In addition MVA has been administered safely to more than 120,000 previously unvaccinated individuals as part of the smallpox eradication campaign in a field study in Germany in the 1970s [97, 98].

#### 7.8. Rationale

As described in section 7.6 the first RH5 vaccine trial in malaria-naïve adults has shown this antigen to be safe, well-tolerated and capable of inducing functional IgG antibodies, in contrast to the responses induced by a lifetime of natural malaria exposure. However the safety and capability of RH5 immunization to induce functional immune responses in adults, young children and infants residing in malaria endemic countries is unknown. This knowledge is essential to inform decisions to proceed with field studies to evaluate efficacy, or to provide more insight into the need to further improve the RH5 vaccine immunogen to focus the response on key neutralising epitopes recognised by the human B cell repertoire. This VAC070 trial will lead to: a) identification of an optimal viral-vectored RH5 dosing schedule that is safe and well-tolerated in young children and infants living in a malaria endemic country; b) characterization of the magnitude and longevity of RH5-specific antibody and cellular immune responses following immunization in a malaria-endemic population; and c) understanding of the quality and epitope-specificity of anti-RH5 IgG antibody following immunization.

Recently, immunization with the liver-stage vaccine candidate, ChAd63-MVA ME TRAP, was shown to be safe and induce ten-fold higher antibody levels in West African babies and infants compared to UK adults and West African adults and young children [99, 100]. Ten-fold higher RH5 IgG antibody responses in infants could well translate into significant protection against clinical malaria in this target population in the field. Notably, pilot deployment trials of the RTS,S/AS01 vaccine are currently focused on the 5-18 month old age group, making this now a key target age group for future assessment with a combination blood-stage vaccine component.

##### 7.9. Rationale for the use of Rabies vaccine as a control vaccine

Rabies vaccine has been chosen as the comparator because: 1) participants will benefit from receiving Rabies vaccine as rabid animals reside in the study area and 2) Rabies vaccine can be administered in a 0, 2-month schedule.

When the Rabies vaccine is administered according to the recommended vaccination schedule (days 0, 7, 21), nearly 100% of subjects attain a protective titer. High antibody titers have also been demonstrated with off-label immunization with Rabies vaccines. Among participants in England, Germany, France and Belgium who received two vaccinations one month apart, nearly 100% of the participants developed specific antibody and the geometric mean titer for the group was 10 IU/mL [101, 102], well above the protective antibody titers > 0.5 IU/mL. The proposed vaccination schedule of 0, 2-months is therefore expected to be successful in conferring protective immunity against rabies among the control participants. However all parents/guardians will be advised that if their child is bitten or scratched by a potentially rabid animal, they must seek medical attention immediately.

#### 8. OBJECTIVES AND OUTCOME MEASURES

This is a dose-escalation, age de-escalation randomised double-blind controlled Phase Ib trial to assess the safety, tolerability and immunogenicity of ChAd63-RH5 administered with MVA-RH5 in a heterologous prime-boost regimen. Adults (18-35 years), young children (1-6 years) and infants (6-11 months) will be enrolled in the study. Safety data will be collected for each of the vaccination regimens. The humoral and cellular immune responses generated by each of these regimens will be assessed.

| 8.1. **Objectives** | 8.2. **Outcome Measures** | 8.3. **Time point(s) of evaluation** |
|---|---|---|
| **Primary Objective**<br><br>To determine safety and tolerability of ChAd63-RH5 and MVA-RH5 given intramuscularly at a 2 month interval in adults (18-35 years), children (1-6 years) and infants (6-11 months) residing in a malaria endemic country. | • Solicited symptoms after vaccination.<br>• Unsolicited symptoms after each vaccination.<br>• Serious adverse events during the study period. | 7-day surveillance after each vaccination.<br><br>28-day surveillance after each vaccination.<br><br>Surveillance from first dose to end of study. |
| **Secondary Objectives**<br><br>To evaluate the magnitude of antibody and cellular immune responses to PfRH5 in adults, children and infants residing in a malaria endemic country.<br><br>To evaluate the quality of antibody and cellular immune responses to RH5 in adults, children and infants residing in a malaria endemic country.<br><br>To evaluate the longevity of antibody and cellular immune responses to fRH5 in adults, children and infants residing in a malaria endemic country. | • Antti-RH5 antibody concentration by ELISA.<br>• Growth inhibition activity of sera from vaccinees on a panel of Pf parasites.<br>• Avidity of anti-RH5 antibodies by ELISA and SPR and/or other assays to be defined.<br>• Cellular immune responses to the RH5 by ELISpot assay and/or Intracellular Cytokine Staining (ICS) and/or other assays to be defined. | At baseline, and days 14 (adults only), 28, 56, 70, 84, 112, 140 and 168 after the first vaccination. |

### 9. TRIAL DESIGN

#### 9.1. Summary of trial design

- Experimental design: Phase Ib, double blind, age de-escalation dose-escalation, randomized (2:1 ratio), controlled trial.
- Healthy adults (18-35 years), young children (1-6 years) and infants (6-11 months) will be screened; those determined to be eligible, based on the inclusion and exclusion criteria, will be enrolled in the study.
- Route of administration: all vaccines will be administered by the intramuscular route to the left deltoid.
- Each participant will be observed for at least 1 hour after vaccination to evaluate and treat any acute adverse events (AEs).
- There will be 7-day follow-up period for solicited AEs post-vaccination: Day 0, 2 and 7 evaluations will be carried out by the study clinician at the study centre and day 1, 3, 4, 5 and 6 evaluations will be carried out by a trained community health worker in the participant’s home, after each vaccination.
- There will be a 28-day (day of vaccination and 28 subsequent days) follow-up after each vaccine dose for reporting unsolicited symptoms.
- Serious adverse events (SAEs) will be recorded throughout the study period. Prior to vaccination, any SAEs due directly to study procedures will be captured. All SAEs will be captured beginning with the administration of the priming dose of ChAd63 RH5 and ending 4 months after the booster dose with MVA RH5.
- Antibodies to RH5_FL will be determined at baseline and 14, 28, 56, 63, 84, 112, 140 and 168 days after ChAd63 RH5 in all participants.
- Cellular immune responses to RH5 will be evaluated at baseline and 14 (adults only), 28, 56, 63, 84 and 168 days after ChAd63 RH5 in all participants.
- The duration of involvement in the study from enrolment will be approximately 6 months. The vaccination phase of the study takes 9 weeks and the post-vaccination follow-up lasts for 4 months after the last dose.
- The vaccine research teams will ensure that insecticide treated bed net is provided to all the study participants and training about its use are conducted.
- Data collection: conventional Case Report Form (CRF) followed by transcription into the OpenClinica Electronic Data Capture system.

#### 9.2. Trial Centre

The study will take place at the IHI Clinical Trial Facility (CTF) located in Bagamoyo town. The facility has clinical consultation rooms, a pharmacy, vaccination room, data management room and an observation area for study participants. There is a resuscitation room that is equipped with oxygen, suction, defibrillator and resuscitation kits. The facility is supported by Bagamoyo Research and Training Centre (BRTC) laboratory located within the grounds of Bagamoyo District Hospital (BDH). BRTC laboratory provides routine haematology, biochemistry and microbiology services. The laboratory can also conduct a wide range of immunology and biomedical analyses.

The clinical trial facility has experienced trained staff including pharmacist, physicians and highly experienced nurses. The facility has Quality Assurance department responsible for ensuring trials are conducted according to protocol and relevant SOP and data collected is of high quality and integrity. Study physician and nurses are ACLS and PALS certified and are involved in regular re-training activities involving simulations.

All participants who require outpatient treatment will be encourage to come to CTF during the day. All cases that require inpatient care will be seen at BDH. A clinician will be accessible 24 hours a day by phone to provide medical care to participants as needed outside the scheduled study visits. BDH is a public hospital providing secondary level health care services. The District Hospital has one paediatric ward and a general ward. There is facility for basic radiology and ultrasound. Routine paediatric surgery is carried out at the District Hospital. Referral to tertiary level facilities is rarely required, but, in cases where such referrals are required participants will be sent to Muhimbili National Hospital in Dar es Salaam. All SAEs will be managed in the inpatient wards in BDH.

##### 9.3. Study Population and Malaria Epidemiology

Participants will be recruited from around Bagamoyo town in Bagamoyo district. Bagamoyo district is one of the 6 districts of the Pwani region of Tanzania (area 8,462.63 $km^2$). According to the 2012 Tanzania National Census, the population of the Bagamoyo District was 343,412. The population of Bagamoyo is highly mixed due to migration and settlement of different ethnic groups. The main economic activities include agriculture, fishing, trade and commerce, and tourism. Bagamoyo has a humid tropical climate with seasonal average temperature ranging from 13˚C to 30˚C with humidity as high as 98%. Rainfall ranges between 800-1200 mm per annum. The short rain season starts from October to December while the long rain season starts from March to May. The driest months are June to September when monthly rainfall is generally less than 50 mm per month.

Malaria remains a public health problem in the district. The district prevalence of malaria among children aged 2 months to 9 years in 2012 was 13% with higher prevalence reported in the western side [66]. The prevalence is much lower in Bagamoyo town. Transmission of malaria tends to be highest between March and May.

#### 9.4. HIV Prevalence

The prevalence of HIV in Tanzania is estimated to be about 5.9% [103]. The prevalence among healthy children in the community is thought to be below 1%. A Prevention of Mother to Child Transmission of HIV program has been operational in Bagamoyo since 2010 and the Ministry of Health anti-viral program is operational. All patients must take part in extensive counselling before treatment. Highly Active Antiretroviral Therapy (HAART) is readily available to all HIV positive people at selected government hospitals.

#### 9.5. Vaccination Services

The United Republic of Tanzania provides immunization services countrywide both on routine and non-routine basis. Immunization services are coordinated by the Immunization and Vaccine Development (IVD) services. This service is available on public and private facilities free of charge. As high as 80% of a total of 5,650 health facilities in the country provides immunization services. Tanzanian policy on immunization calls for support for routine immunization to accelerate the control of vaccine preventable diseases. The following vaccines are provided BCG, OPV, DTP-HepB-Hib (pentavalent), pneumococcal, rotavirus, Tetanus Toxoid, vitamin A, Measles Rubella and HPV. The immunization schedule in mainland Tanzania is shown in Table 1.

Table 1 Immunization Schedule in Tanzania.

| Antigen | Time of administration |
|---|---|
| BCG, OPV 0 | At birth or first contact |
| OPV1, DTP-HepB-Hib1,PCV 1, Rota 1 | 6 Weeks of age |
| OPV2, DTP-HepB-Hib2, PCV 2, Rota 2 | 10 Weeks of age |
| OPV3, DTP-HepB-Hib3 , PCV3, IPV | 14 Weeks of age |
| MR 1 | 9 Months of age |
| MR 2 | 18 Months of age |
| HPV 1 | 9 years |
| HPV 2 | 6 months after 1st dose |

The Immunization and Vaccine Development (IVD) services are provided through four main strategies. The primary strategy is through routine immunization services at health facilities. The secondary strategies include outreach services to remote communities and mop-up campaigns aimed at selected districts to capture defaulters and reach out to children missed in routine services. Finally there are occasional national vaccination campaigns which are implemented to reach large populations in a given period, as a supplementary activity to the routine immunization in order to increase the immunity in the community.

With the exception of the measles vaccine, the infant participants (6-11 months) would have received all immunizations by the time of enrolment. However, it is possible that a few defaulters may be encountered during screening. Vaccination with the IP will be delayed for 28 days after vaccination with measles vaccine and 14 days after non-live vaccines.

#### 9.6. Rationale for Trial Design

*Administration Schedules*

Heterologous prime boost approach using ChAd63-MVA vectors has been shown to be effective in inducing potent T cell responses and antibody responses to the specific expressed antigens. Recently, a Phase Ia clinical trial of RH5 in malaria-naïve UK adults has primed participants with ChAd63 RH5 followed by a boost of MVA RH5 eight weeks later. The regimen induced functional antibodies in all subjects which blocked parasite invasion of erythrocytes *in vitro*, with functional activity associated with antibody levels. For this reason, and to provide comparability with the UK study, we will use a similar administration schedule.

A recent study in West Africa has also shown that ChAd63-MVA vectors induced ten-fold higher levels of antibody in infants compared to adults in West Africa and the UK. It is therefore expected that the approach will result in higher levels of antibodies to RH5, which would translate into better growth inhibition.

*Route & Dose*

All vaccines will be administered intramuscularly in the left deltoid. Intramuscular administration of antigens using ChAd63-MVA platform has been shown to be safe and well tolerated and this is a more practical form of administration in the field. The study will determine the ideal dose based on the safety data that will be collected. Dose escalation will occur only when the safety data from previous lower dose has been reviewed by the independent safety monitors and the approval given to proceed.

ChAd63 expressing RH5 has been given to 24 healthy adults as part of a Phase Ia dose escalation study of the ChAd63 RH5 and MVA RH5 vaccines, VAC057, conducted in Oxford and Southampton. The doses used in the study were $5 \times 10^9$ vp and $5 \times 10^{10}$ vp. Similarly MVA RH5 has been administered to sixteen individuals as part of the Phase Ia study VAC057 and the doses used were $1X10^{8}$ and $2x10^{8}$ pfu. Data from these studies can also be used to predict the expected adverse event profile following vaccination with MVA RH5 in this study. The safety profile of the vaccines was similar to those seen in previous studies with the same viral vector.

A dose of $5x10^{10}$ vp ChAd63 and $2x10^{8}$ pfu MVA expressing RH5 will be used in adults. This choice is based on the good safety and tolerability data from malaria naïve adults in the UK. Furthermore, there are substantial data showing good safety and tolerability in malaria naïve and malaria exposed adults, children and infants who received the same doses of ChAd63 and MVA expressing other malaria antigens [37, 38, 99, 104].

Because this is the first time PfRH5 is administered in young children and infants, we will start with lower doses of ChAd63 and MVA expressing RH5 and increase the dose to the final dose of $5 \times 10^{10}$ vp ChAd63 and $2 \times 10^{8}$ pfu MVA once the safety data from the lower dose have been reviewed. It is therefore important to notice that the dose of $5 \times 10^{10}$ vp ChAd63 and $2 \times 10^{8}$ pfu MVA will only be administered if lower doses are found to be safe.

Administration of equal doses of MVA and ChAd63 in adults and children is based on safety and immunogenicity data from previous studies. Unlike medications, use of similar doses in adults and children for the vaccine is not an uncommon phenomenon due to differences in mechanisms of action[105]. For instance the prophylactic dose for the yellow fever vaccine is 0.5ml in both adults and children (>9 months)[106, 107]. Safety and reactogenicity information is more critical in selection of the vaccine doses. We have established a robust design to carefully assess the safety and reactogenicity of lower doses before proceeding to higher doses (section 9.7).

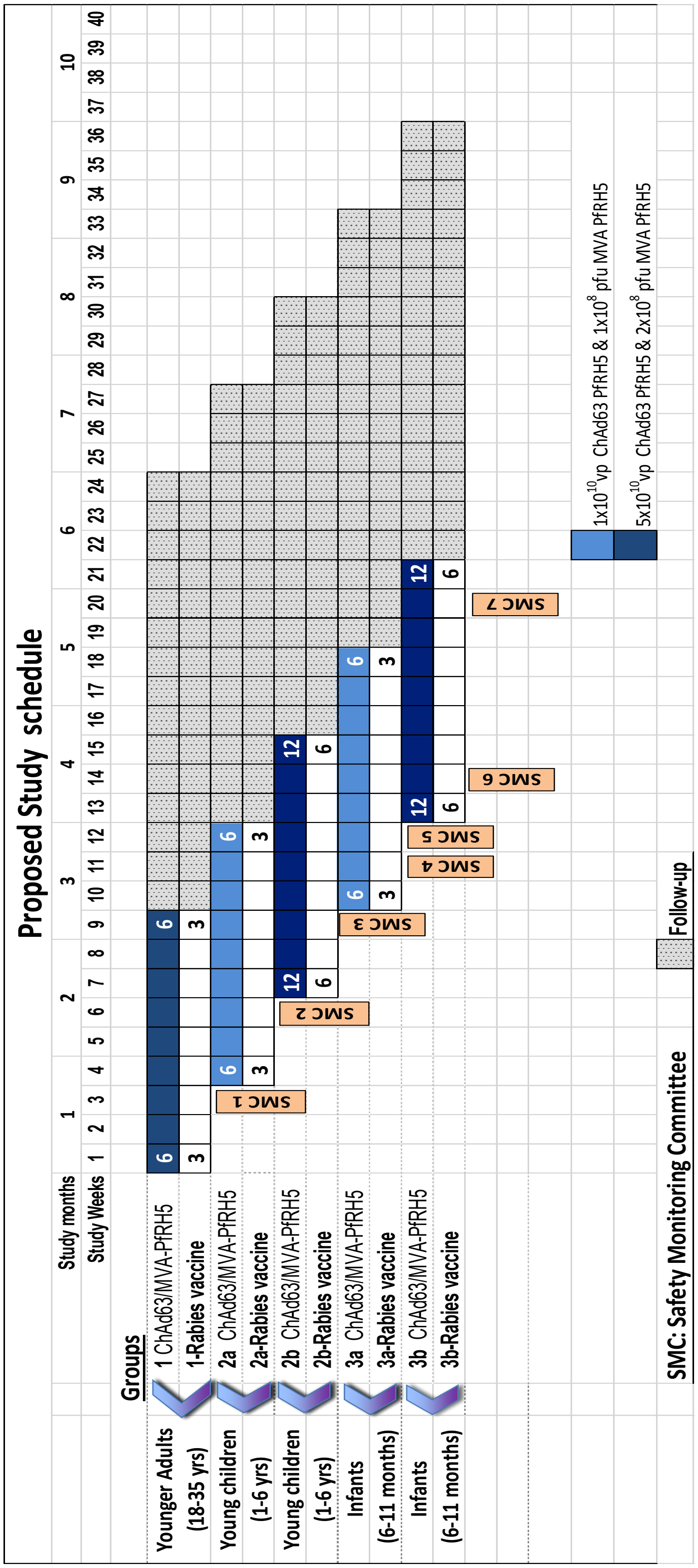

*Figure 7 : Study groups and schedule*

#### 9.7. Safety oversight during age de-escalation and dose-escalation

To assure safety, vaccinations will start in adults and will progress to young children and infants using staggered start dates at approximately 3 weekly intervals. In young children, and infants we will prime with 1 x $10^{10}$ vp ChAd63 RH5 and boost with 1 x $10^{8}$ pfu MVA RH5 before escalating to 5 x $10^{10}$ vp ChAd63 RH5 priming and 2 x $10^{8}$ pfu MVA RH5 boosting (Figure 7).

The approval to proceed to a higher dose within an age-group as well as age de-escalate to young children and infants will be sought from the independent SMC after the review of the safety data from the preceding dose and vaccination groups.

Progression to the higher dose or continuation of vaccination within the group may take place as long as none of the following specified criteria are met:

- Two (calculated as 22% of 9) or more participants in either Group 1, 2a or 3a experience the same grade 3 solicited local adverse event beginning within 2 days after vaccination (day of vaccination and one subsequent day) and persisting at Grade 3 for > 72 hrs.
- Four (calculated as 22% of 18) or more participants in either Group 2b or 3b experience the same grade 3 solicited local adverse event beginning within 2 days after vaccination (day of vaccination and one subsequent day) and persisting at Grade 3 for > 72 hrs.
- Two (calculated as 22% of 9) or more participants in either Group 1, 2a or 3a experience the same grade 3 solicited systemic adverse event beginning within 2 days after vaccination (day of vaccination and one subsequent day) and persisting at Grade 3 for > 48 hrs.
- Four (calculated as 22% of 18) or more participants in either Group 2b or 3b experience the same grade 3 solicited systemic adverse event beginning within 2 days after vaccination (day of vaccination and one subsequent day) and persisting at Grade 3 for > 48 hrs.
- Two (calculated as 22% of 9) or more participants in either Group 1, 2a or 3a experience the same grade 3 unsolicited adverse event beginning within 2 days after vaccination (day of vaccination and one subsequent day) and persisting at Grade 3 for > 48 hrs that is deemed possibly, probably or definitely related to study product administration.
- Four (calculated as 22% of 18) or more participants in either Group 2b or 3b experience the same grade 3 unsolicited adverse event beginning within 2 days

after vaccination (day of vaccination and one subsequent day) and persisting at Grade 3 for > 48 hrs that is deemed possibly, probably or definitely related to study product administration.

- One participant experiences an SAE deemed possibly, probably or definitely related to study product administration.

If any of these criteria are met, unblinding will be done to find out if the participants experiencing a grade 3 adverse event or the participant experiencing an SAE received RH5 as opposed to Rabies Vaccine. If it is found the participant(s) received RH5, all vaccinations will be put on temporary hold until guidance from the SMC is obtained to proceed with vaccinations.

The SMC will be convened by the study Sponsor and will be provided with a formal safety report to review, and they will provide recommendation with regard to proceeding with vaccination. The two age de-escalations into younger children and infants will take place approximately 3 weeks after the adults' and younger children's first vaccinations respectively, generation of safety report and its review by the SMC.

Details of the age de-escalation/ escalation scheme are shown in Figure 7 and also described here:

(**SMC Review 1**) After vaccinations have been completed in Group 1, safety and tolerability data up until a minimum of one week after vaccination with 2 $x10^{10}$ vp ChAd63 RH5 will be assembled into a formal safety report and reviewed by the SMC. If recommended by the SMC and approved by the Sponsor's representative (CI), the trial will continue by immunizing the younger children (1-6 years).

(**SMC Review 2**) Safety and tolerability data up until a minimum of one week after the completion of vaccination of younger children with $1x10^{10}$ vp ChAd63 RH5 will be assembled into a formal safety report and reviewed by the SMC. If recommended by the SMC and approved by the CI, the trial will continue by immunizing young children (1-6 years) with the $5x10^{10}$ vp ChAd63 RH5.

(**SMC Review 3**) Safety and tolerability data up until a minimum of one week after the completion of vaccination of younger children with $5x10^{10}$ vp ChAd63 will be assembled into a formal safety report and reviewed by the SMC. If recommended by the SMC and approved by the CI, the trial will continue by immunizing infants (6-11 months) with 1 x $10^{10}$ vp ChAd63 RH5.

(**SMC Review 4**) Safety and tolerability data up until a minimum of one week after the completion of vaccination of adults with 2 x $10^{8}$ pfu MVA RH5 will be assembled into a formal safety report and reviewed by the SMC. If recommended by the SMC and approved by the CI, the trial will continue by immunizing young children (1-6 years) with 1 x $10^8$ pfu MVA RH5.

(**SMC Review 5**) Safety and tolerability data up until a minimum of one week after the completion of vaccination of infants (6-11 months) with 1 x $10^{10}$ vp ChAd63 RH5 will be assembled into a formal safety report and reviewed by the SMC. If recommended by the SMC and approved by the CI, the trial will continue by immunizing infants (6-11 months) with 5 x $10^{10}$ vp ChAd63.

(**SMC Review 6**) Safety and tolerability data up until a minimum of one week after the completion of vaccination of young children (1-6 years) with 1 x $10^8$ pfu MVA RH5 will be assembled into a formal safety report and reviewed by the SMC. If recommended by the SMC and approved by the CI, the trial will continue by immunizing young children (1-6 years) with 2 x $10^8$ pfu MVA PfRH5.

(**SMC Review 7**) Safety and tolerability data up until a minimum of one week after the completion of vaccination of infants (6-11 months) with 1 x $10^8$ pfu MVA RH5 will be assembled into a formal safety report and reviewed by the SMC. If recommended by the SMC and approved by the CI, the trial will continue by immunizing infants (6-11 months) with 2 x $10^8$ pfu MVA RH5.

*<u>Sentinel Participants</u>*

As an additional safety measure, three sentinel participants from each group will be inoculated two days before the remaining participants are vaccinated with ChAd63-RH5 and MVA-RH5. Two will receive RH5 and one Rabies vaccine. The remaining participants will be vaccinated if neither of the following criteria is met;

- Two participants experience at least one grade 3 solicited local adverse event beginning within 2 days after vaccination (day of vaccination and one subsequent day) and persisting at Grade 3 for > 72 hrs.
- Two participants experience at least one grade 3 solicited systemic adverse event beginning within 2 days after vaccination (day of vaccination and one subsequent day) and persisting at Grade 3 for > 48 hrs.
- Two participants experience at least one grade 3 unsolicited adverse event beginning within 2 days after vaccination (day of vaccination and one subsequent day) and persisting at Grade 3 for > 48 hrs that is deemed possibly, probably or definitely related to study product administration.

- One participant experiences an SAE deemed possibly, probably or definitely related to study product administration.

If either of these criteria are met, and before an SMC review is called, it will be ascertained whether both participants experiencing a grade 3 adverse event or the one participant experiencing an SAE did receive RH5. If the participant(s) received RH5, the SMC will review the case and advise the CI and the Investigator if they should continue with vaccinations.

### 10. PARTICIPANT IDENTIFICATION

#### 10.1. Sensitization

After approval from ethics committees, local authorities will be informed about the study in order to receive permission to approach the community. A series of meetings will then be held to explain the study to the potential participants in the community. These meetings will consist sensitization meetings for general information and pre-screening meetings for more detailed information. Advertisements will be done using locally acceptable procedures to inform and invite potential participants and parents/guardians to the sensitization meetings. Volunteers that previously participated in an epidemiological study conducted at IHI clinical trial facility using similar sensitization procedures and have consented to be contacted for possible participation in a study like this as per the initial informed consent, may also be invited to the sensitization meetings. The local administrative leaders will be invited to be present for these meetings. The need and the difficulties of developing a vaccine against malaria will be discussed in a culturally appropriate fashion, as well as an outline of the proposed trial, including the rationale, the background data available, the study objectives and the screening and consent procedures. Particular attention will be paid to study procedures, immunization and blood collection. To avoid misunderstandings the purpose of blood collection and the associated risks will be explained in a culturally sensitive fashion. Opportunity will be provided for the community members to ask questions and get responses from the Investigators.

Potentially interested participants including the parents/guardians will be asked to register their names at the end of the meetings with the study staff. Potential participants will be invited to attend a pre-screening meeting at CTF.

At the pre-screening meeting the PI and trained study clinicians and nurses will provide detailed information about the study using the PIS in a group and individual sessions. A copy of the PIS will also be provided before commencing these meetings. The PI will ensure potential participants or parents/guardians fully understand the risks and benefits associated with participation in the study.

It will be stressed that RH5 is an experimental vaccine and cannot be guaranteed to provide protection and that therefore it will still be necessary to seek treatment for possible malaria even after vaccination and with the use of available preventive measures including insecticide treated bed nets. It will also be made clear that the study will require HIV testing of the participants. It will be explained that eventually the plan is to offer immunization to everyone including those testing HIV positive, but that normal practice in intervention evaluation is to begin with those who are HIV negative until the safety profile is better defined. It will be made clear that screening on enrolment will include a range of diseases (not just HIV), so exclusion from participating in the trial does not mean being HIV positive and the confidentiality of those participants infected with HIV will be protected. It will be stressed that this is the beginning of a long process of RH5 vaccine development, which will be accelerated by the conduct of similar trials in Africa.

Opportunity will be provided for the potential participants to ask questions and get responses from the PI and study physicians. Those who are still interested to take part will then be invited for a formal screening visit where an informed consent form will be signed or thumb-printed. A copy of the PIS will be given to the potential participants to take home and read it further including consulting their spouses or relatives.

##### 10.2. Trial Participants

The study will recruit healthy young adults (18-35 years), young children (1-6 years) and infants (6-11 months) residing in Bagamoyo town, Tanzania. The participants will be enrolled into three groups corresponding to the ages as shown in Figure 7.

##### 10.3. Inclusion Criteria

Only participants who meet all the inclusion criteria will be enrolled into the trial;

- Group 1: Healthy male or female adults aged 18-35 years at the time of enrolment with signed consent.
- Group 1 (Female only participants): Must be non-pregnant (as demonstrated by a negative urine pregnancy test), and provide consent of their willingness to take Depo-Provera contraceptive during the study and safety follow-up period.
- Groups 2a & 2b: Healthy male or female young children aged 1-6 years at the time of enrolment with signed consent obtained from parents or guardians.
- Groups 3a & 3b: Healthy male or female infants aged 6-11 months at the time of enrolment with signed consent obtained from parents or guardians.
- Planned long-term (at least 9 months from the date of recruitment) or permanent residence in Bagamoyo town.
- Adults with a Body Mass Index (BMI) 18 to 30 $Kg/m^2$; or young children and infants with Z-score of weight-for-age within ±2SD.

##### 10.4. Exclusion Criteria

The participant may not enter the trial if ANY of the following apply:

- Clinically significant congenital abnormalities as judged by the PI or other delegated individual.
- Clinically significant history of skin disorder (psoriasis, contact dermatitis etc.), allergy, cardiovascular disease, respiratory disease, endocrine disorder, liver disease, renal disease, gastrointestinal disease and neurological illness as judged by the PI or other delegated individual.

- Any confirmed or suspected immunosuppressive or immunodeficient state, including HIV infection; asplenia; recurrent, severe infections and chronic (more than 14 days) immunosuppressant medication within the past 6 months (inhaled and topical steroids are allowed).
- History of cancer (except basal cell carcinoma of the skin and cervical carcinoma in situ).
- Weight for age z-scores below 2 standard deviations of normal for age.
- History of allergic disease or reactions likely to be exacerbated by any component of the vaccines, e.g. egg products, Kathon, neomycin, betapropiolactone.
- Any history of anaphylaxis in relation to vaccination.
- Clinically significant laboratory abnormality as judged by the PI or other delegated individual.
- Blood transfusion within one month of enrolment.
- History of vaccination with previous experimental malaria vaccines.
- Administration of immunoglobulins and/or any blood products within the three months preceding the planned administration of the vaccine candidate.
- Participation in another research study involving receipt of an investigational product in the 30 days preceding enrolment, or planned use during the study period.
- Seropositive for hepatitis B surface antigen (HBsAg) or hepatitis C (HCV IgG).
- Any other finding which in the opinion of the PI or other delegated individual would increase the risk of an adverse outcome from participation in the trial.
- Likelihood of travel away from the study area.
- Positive malaria by blood smear at screening.
- Female participant who is pregnant, lactating or planning pregnancy during the course of the trial.
- Scheduled elective surgery or other procedures requiring general anaesthesia during the trial.
- Any other significant disease, disorder or situation which, in the opinion of the Investigator, may either put the participants at risk because of participation in the trial, or may influence the result of the trial, or the participant’s ability to participate in the trial.

#### 10.5. Contraindications to subsequent vaccination

*Indications for deferral of vaccination*

These events constitute contraindications to administration of the IP at that point in time and will result in deferral of vaccination to another time within the protocol specified window, or withdrawal at the discretion of the Investigator. AEs should be followed-up according to the instructions in Section 13.

- Acute disease at the time of administration of the IP (acute disease is defined as the presence of a moderate or severe illness with or without fever). All vaccines can be administered to persons with a minor illness such as diarrhoea or mild upper respiratory infection without fever, i.e. axillary temperature < 37.5°C.
- Axillary temperature of ≥ 37.5°C.
- Abnormal laboratory parameters that have been determined to be clinically significant.
- Diphtheria, Tetanus, Pertussis (whole-cell), *Hemophilus influenzae* type B, Measles, or Polio vaccination within 14 days of schedule to receive the IP.
- On request of the participant.
- On advice of the safety monitor.
- On advice of the Investigators.

*Permanent Contraindications for further vaccination*

The following AEs constitute absolute contraindications to further administration of the IP. If any of these AEs occur during the study, the participant must not receive additional doses of vaccine, but will continue with the full safety monitoring procedure (if the participant has already received a vaccination) as per protocol:

- Acute allergic reaction, significant IgE-mediated event or anaphylactic shock following the administration of vaccine investigational product.
- Any confirmed or suspected immunosuppressive or immunodeficient condition, including human immunodeficiency virus (HIV) infection.
- The participant experiences an SAE that is determined to be related to the study product.
- The participant starts taking a concomitant medication for a chronic illness.
- Pregnancy.
- On request of the participant.
- On advice of the Independent SMC.
- On advice of the Investigators.
- On advice of the IRB, regulatory authority or SMC.

*Withdrawal of study participant*

Participants have a right to withdraw from the study at any time and are not obliged to give his or her reasons for doing so. The Investigator may withdraw the participant at any time in the interests of the participant’s health and well-being. In addition the participant may withdraw/be withdrawn for any of the following reasons:

- Any serious adverse event (SAE) deemed to be related to the IP.

- Any adverse event (AE) that, according to the clinical judgment of the Investigator, is considered a contraindication to proceeding with the study procedures.
- The use of concomitant, chronic medication active on the immune system (steroids, immunosuppressive agents) or potentially affecting ChAd63 MVA RH5.
- Participant completely lost to follow-up.
- Poor compliance with study procedures, such that completeness or integrity of data are jeopardized.
- Administrative decision by the Investigator.
- Ineligibility (either arising during the study or retrospectively, having been overlooked at screening).
- Significant protocol deviation.

If a participant withdraws from the study, blood samples collected before withdrawal will be used/ stored to address the safety and immunogenicity objectives of the current protocol unless the participant specifically requests otherwise. All participants can withdraw their permission to store the samples for future studies at any time."

*Replacement of Individual Participants after Withdrawal*

If for any reason a participant cannot receive the first vaccination, they can be replaced by a backup participant that has been screened and found to be eligible. This can only happen on the day of ChAd63 RH5 vaccination for all the groups. However, if the participant is withdrawn after receiving ChAd63 RH5, there will be no replacement.

##### 10.6. **Pregnancy**

Should a participant become pregnant during the trial, she will not be vaccinated but will be followed up as other participants and in addition will be followed until pregnancy outcome, with the participant's permission. We will not routinely perform venepuncture on such participants.

The reason for withdrawal will be recorded in the CRF. If the participant is withdrawn due to an adverse event, the Investigator will arrange for follow-up visits or telephone calls until the adverse event has resolved or stabilised.

### 11. TRIAL PROCEDURES

#### 11.1. Informed Consent

The participant (Group 1) and parent/guardian of the participant (Groups 2 and 3) must personally sign and date the latest approved version of the Informed Consent form before any trial specific procedures are performed.

Written versions of the Participant Information Sheet (PIS) and Informed Consent Form (ICF) will be presented to the participants detailing no less than: the exact nature of the trial; what it will involve for the participant; the implications and constraints of the protocol; the known side effects and any risks involved in taking part. It will be clearly stated that the participant is free to withdraw from the trial at any time for any reason without prejudice to future care, without affecting their legal rights and with no obligation to give the reason for withdrawal.

All participants (Group 1) and parents/guardians of the participants (Groups 2 and 3) will be required to demonstrate their understanding of the study by answering a questionnaire which will consist of 10 true/false statements that will be instituted by study personnel. Participants must respond correctly to all 10 questions. They will have a maximum of two attempts after which those who fail to correctly answer all questions will be excluded. Those who correctly respond to all 10 questions will be allowed to sign the ICF.

The participant will be allowed as much time as wished to consider the information, and the opportunity to question the Investigator, or other independent parties, to decide whether they will participate in the trial. Written Informed Consent will then be obtained by means of participant-dated signature or thumb-print and dated signature of the person who presented and obtained the Informed Consent. If the participant is illiterate a literate witness will be present during the consent process and verify the process by signing and dating the ICF. Consenting will be done by trained study staff including the clinicians and nurses. A copy of the signed ICF will be given to the participant. The original signed form will be retained at the trial site.

#### 11.1. Screening and Eligibility Assessment

Screening procedures will only start once a participant has provided a written informed consent and continue until all 63 participants and at least 2 back-up participants have been enrolled into the study. Two screening visits will be conducted for each participant.

*Screening Visit 1*

Only participants who meet eligibility criteria and have signed/thumb-printed the informed consent will undergo a full screening procedure. At screening visit 1, study staff will go through the PIS for the second time and describe the study in detail. This will be done in a group session where potential participants will be encouraged to ask questions. After the group session each participant will have a private session with study staff where additional clarification will be provided and time for more questions provided.

Participants will be assigned with a study ID on a first come first serve basis. A photo will also be taken to allow preparation of Study ID cards which will be given after the first vaccination. In the interim all participants will receive temporary study ID forms.

The following procedures will then take place:

- Taking medical history of a participant.
- Complete physical examination including vital signs, height, weight and blood pressure.
- Venous blood sample collection for the following analyses:
    - Haematology: Full Blood Count (CBC) which includes: red blood cells, platelets, white blood cell count, lymphocyte count, haemoglobin, mean corpuscular volume and mean corpuscular haemoglobin concentration.
    - Biochemistry: serum creatinine and ALT.
    - Serology: HIV, HCV and HBV tests.
    - Thick blood smear and parasite genotyping.
    - Baseline humoral immune responses (antibodies to RH5);
    - Baseline cellular immune responses to RH5;
- Collection of urine for the following analyses:
    - Dipstick for blood, glucose and protein.
    - Pregnancy test (for women in Group 1 only).
- Checking of inclusion and exclusion criteria

Once all specific visit procedures are completed, participants will be allowed to go home and asked to return for Screening Visit 2.

The screening laboratory results will be reviewed by study clinicians before participants are seen at screening visit 2. These procedures will be documented in the case report forms (CRFs) and relevant clinical notes, which will be kept in the individual study participant’s file. After collection of all screening information, the study team will review and assess the eligibility of each participant. All participants will be contacted and informed of the specific date to come to CTF for Screening Visit 2.

*Screening Visit 2*

During Screening Visit 2, participants will be informed of their screening results and receive HIV and hepatitis post-test counselling. Study clinicians will inform them if they are eligible for enrolment into the study. The principal investigator (PI) will have the final decision on the eligibility of a participant. Only those laboratory results with clinical relevance will be provided to the participant. Immunology results will be summarized and presented as part of final feedback to participants at the end of the study. Participants excluded from this trial because of clinically significant medical conditions may be managed initially by study clinicians and/or referred to the routine health care system for further evaluation and treatment as necessary. In the event that a participant tests positive for HIV, HBV, or HCV, he or she will be referred to the appropriate public health facility for further counselling and treatment. Participants (and their parents/guardians if applicable) will be provided with ITNs at this visit and will be encouraged to use them to reduce the risk of natural malaria infection.

Recruitment will continue until all the required number of participants per Group has fulfilled all inclusion criteria and none of the exclusion criteria. Should the participant or the parents of participants change their mind and decline to participate prior to immunization, additional participants will be screened the required number of participants per group is enrolled. A list of eligible participants per group will be generated and used to identify the participants eligible for immunization. Screening tests will be completed within 30 days prior to immunization.

##### 11.2. Baseline Assessments

Before vaccination all participants with undergo baseline assessment. This will include collection of safety and immunology samples. Once all final eligibility criteria are reviewed including safety laboratory results, eligible participants will be vaccinated. The window period as shown in the appendix 1 will be used in case a participant cannot be available on a scheduled date. Pre-vaccination laboratory assessments will be done within 24 hours prior to vaccination. However only clinical assessments and vital signs collected on the day of vaccination will be recorded in the pre-vaccination CRF.

##### 11.3. Randomisation procedure

Generation of a randomization list will be done by an independent statistician from IHI. The randomization list will contain sequential codes (Treatment numbers) linking a study ID to a vaccine assignment. Study ID will be assigned to participants in the order in which they are enrolled in the trial. Access to the randomization list will be exclusively limited to the study pharmacist and the independent statistician(s) at IHI. These individuals will be unblinded and will not be involved in the evaluation of the study participants.

Participants will be assigned to groups based on their age. The various groups, characterized by age and their doses, will be enrolled sequentially. Randomization into vaccine or control will take place within each group according to a 2:1 ratio.

Participants will receive Study ID cards with Subject Numbers at the screening visit 1. At first vaccination visit, after verification of eligibility criteria, subjects will be allocated a Treatment Number in the order they present for vaccination with the lowest Treatment Number available from the randomization list. The correspondence between the Subject Number and the Treatment Number will be noted down in the CRF and subsequently entered in the OpenClinica Electronic Data Capture (EDC) system.

##### 11.4. Blinding

Data pertaining to ChAd63 RH5/MVA RH5 or rabies vaccine will be collected in a double blinded manner. Double blinded means that the vaccine recipient or their parent(s)/guardian(s) as well as those responsible for the evaluation of safety and immunogenicity endpoints will all be unaware which treatment, ChAd63/MVA RH5 or rabies vaccine, was administered to a particular subject. The only study staff aware of the vaccine assignment will be those responsible for the storage and preparation of vaccines; these staff will play no other role in the study.

Measures will be taken to keep participants and clinical Investigators (including the PI) and all other staff involved in measuring study outcomes blinded to treatment allocation. Although the final constituted test article will be similar to rabies vaccine in appearance, the two will come under distinct packages. Therefore, blinding of the individuals preparing the study vaccine (such as the study pharmacist and syringe preparation team) will not be possible. Since the test article and comparison vaccines can be distinguished by their packaging and labels, the vaccine preparation area and the immunizing area will be physically separated. The vaccine preparer, a pharmacist, will be exclusively dedicated to vaccine preparation.

The Local Independent Safety Monitor will be provided sealed code-break envelopes for each participant and may unblind himself to the vaccine allocation of individual study participants if deemed urgently necessary for medical and/or ethical reasons (for example, in the case of a participant in urgent need of rabies immunization). The Local Independent Safety Monitor will also keep one set of the master randomization list in a sealed envelope, in the event that emergency unblinding is required.

##### 11.5. Detailed scheduled study visits

Day 0: (Vaccination with ChAd63 RH5 or rabies vaccine)

Participants will first be identified using their Study ID cards and vital signs and body weight will be measured.

Blood sample (if not already done within the last 48 hours) will be collected for the following analyses:

- haematology (full blood count);
- biochemistry (including creatinine and ALT);
- Immunology:
    - humoral immune responses to RH5;
    - cellular immune responses to RH5
    - transcriptome analysis;

Medical history and focused physical examination (informed by complaints) will be conducted by study clinicians who will also check for inclusion/exclusion criteria, contraindications to vaccination and elimination criteria. Safety laboratory results will be reviewed by study clinician before vaccination and checked by the PI (or designee). If participant is confirmed to be eligible they will be randomized to receive either ChAd63 RH5 or rabies vaccine according to the site randomization SOP.

ChAd63 RH5 or rabies vaccine will then be administered intramuscularly in the left deltoid by trained study staff according to the site vaccination SOP. Each participant will be assessed for at least 1 hour after vaccination to evaluate and treat any acute adverse events. During this time the following will be recorded;

- Post vaccination vital signs
- Post-vaccination solicited signs/symptoms.
- Post-vaccination unsolicited adverse events.
- Post-vaccination SAEs.
- Any concomitant medication.

Days 1, 3, 4, 5 and 6 (Post-Vaccination with ChAd63 RH5)

Each participant will be visited at home on day 1, 3, 4, 5 and 6 by a community health worker (CHW) for assessment and recording of any solicited and unsolicited AEs. If necessary the participant will continue to be seen regularly until the AEs have resolved or stabilised. The CHW will be blinded to the group allocation of the subject. During the home visit, the following will be assessed and recorded in the CRF;

- Axillary body temperature.
- Local and general solicited adverse events.
- SAEs experienced by the participant since the last visit.
- Unsolicited adverse events experienced by the participant since the last visit.
- Concomitant medication taken.

Day 2 (-1/+1) days; Post-Vaccination with ChAd63 RH5)

Participants will be seen at CTF where medical history, temperature monitoring and physical examination will be performed and recorded in the CRF. AEs will be recorded in the CRF.

Day 7 (-1/+3 days; Post-Vaccination with ChAd63 RH5)

Participants will be seen at CTF where medical history, temperature monitoring and physical examination will be performed and recorded in the CRF. AEs will be recorded in the CRF.

Blood sample will be collected for the analysis of:

- haematology (full blood count);
- biochemistry (including creatinine and ALT);

Day 14 (-1/+3 days; Post-Vaccination with ChAd63 RH5)

Participants will be seen at CTF where medical history, temperature monitoring and physical examination will be performed and recorded in the CRF. AEs will be recorded in the CRF.

Blood sample will be collected for the analysis of:

- haematology (full blood count);
- biochemistry (including creatinine and ALT);
- Immunology:
    - humoral immune responses to RH5;
    - cellular immune responses to RH5 (for adults only) ;
    - transcriptome analysis;

Day 28 (-7/+2 days; Post-Vaccination with ChAd63 RH5)

Participants will be seen at CTF where medical history, temperature monitoring and physical examination will be performed and recorded in the CRF. AEs will be recorded in the CRF.

Blood sample will be collected for the analysis of:

- haematology (full blood count);
- biochemistry (including creatinine and ALT);
- Immunology:
    - humoral immune responses to RH5;
    - cellular immune responses to RH5 ;
    - transcriptome analysis;

Day 56 (Vaccination with MVA RH5 or rabies vaccine) (± 7days); Post-Vaccination with ChAd63 RH5)

Participants will first be identified using their Study ID cards and vital signs and body weight will be measured. Medical history and focused physical examination (informed by complaints) will be conducted by study clinicians who will also check for contraindications to vaccination and elimination criteria.

Blood sample will be collected for the pre-vaccination assessment of (if not already done within the last 48 hours):

- haematology (full blood count);
- biochemistry (including creatinine and ALT);
- Immunology:
    - humoral immune responses to RH5;
    - cellular immune responses to RH5 ;
    - transcriptome analysis;
- symptomatic malaria parasitaemia (only if participant is symptomatic)
- asymptomatic malaria parasitaemia (retrospectively);

- malaria parasite genotyping (retrospectively);

Safety laboratory results will be reviewed by the study clinician before vaccination and checked by the PI (or designee).

MVA RH5 or rabies vaccine will then be administered intramuscularly in the left deltoid by trained study staff according to the site vaccination SOP. Each participant will be assessed for at least 1 hour after vaccination to evaluate and treat any acute adverse events. During this time the following will be recorded;

- Post vaccination vital signs
- Post-vaccination solicited signs/symptoms.
- Post-vaccination unsolicited adverse events.
- Post-vaccination SAEs.
- Any concomitant medication.

Days 57, 59, 60, 61 and 62 (Post-Vaccination with MVA RH5)

Each participant will be visited at home daily on days 57, 59, 60, 61 and 62 by a community health worker (CHW) for assessment and recording of any solicited and unsolicited AEs. If necessary the participant will continue to be seen regularly until the AEs have resolved or stabilised. CHW will be blinded to the group allocation of the subject. During the home visit, the following will be assessed and recorded in the CRF;

- Axillary body temperature.
- Local and general solicited adverse events.
- SAEs experienced by the vaccinee since the last visit.
- Unsolicited adverse events experienced by the vaccinee since the last visit.
- Concomitant medication taken.

Day 58 (-1/+1 days; Post-Vaccination with ChAd63 RH5)

Participants will be seen at CTF where medical history, temperature monitoring and physical examination will be performed and recorded in the CRF. AEs will be recorded in the CRF.

Day 63 (-1/+3 days; Post-Vaccination with ChAd63 RH5)

Participants will be seen at CTF where medical history, temperature monitoring and physical examination will be performed and recorded in the CRF. AEs will be recorded in the CRF.

Blood will be collected for analysis of:

- haematology (full blood count);
- biochemistry (including creatinine and ALT);
- symptomatic malaria parasitaemia (only if participant is symptomatic);
- asymptomatic malaria parasitaemia (retrospectively);

- malaria parasite genotyping;
- Immunology:
    - humoral immune responses to RH5;
    - cellular immune responses to RH5 ;
    - transcriptome analysis;

Day 70 (-1/+3 days; Post-Vaccination with ChAd63 RH5)

Participants will be seen at CTF where medical history, temperature monitoring and physical examination will be performed and recorded in the CRF. AEs will be recorded in the CRF.

Blood will be collected for analysis of:

- haematology (full blood count);
- biochemistry (including creatinine and ALT);
- symptomatic malaria parasitaemia (only if participant is symptomatic);
- asymptomatic malaria parasitaemia (retrospectively);
- malaria parasite genotyping;

Day 84 (-7/+2 days; Post-Vaccination with ChAd63 RH5)

Participants will be seen at CTF where medical history, temperature monitoring and physical examination will be performed and recorded in the CRF. AEs will be recorded in the CRF.

Blood sample will be collected for the analysis of:

- haematology (full blood count);
- biochemistry (including creatinine and ALT);
- Immunology:
    - serology (antibodies to RH5);
    - cellular immune responses to RH5;
    - transcriptome analysis;
- symptomatic malaria parasitaemia (only if participant is symptomatic);
- asymptomatic malaria parasitaemia (retrospectively);
- malaria parasite genotyping (retrospectively);

Day 112 (-1/+7 days; Post-Vaccination with ChAd63 RH5)

Participants will be seen at CTF where medical history, temperature monitoring and physical examination will be performed and recorded in the CRF. AEs will be recorded in the CRF.

Blood sample will be collected for the analysis of:

- Symptomatic malaria parasitaemia (only if participant is symptomatic)
- Asymptomatic malaria parasitaemia;
- Malaria parasite genotyping.
- Immunology:
    - humoral immune responses to RH5;

Day 140 (±7 days; Post-Vaccination with ChAd63 RH5)

Medical history, temperature monitoring and physical examination will be performed and recorded in the CRF. AEs will be recorded in the CRF.

Blood sample will be collected for the analysis of:

- haematology (full blood count);
- biochemistry (including creatinine and ALT);
- symptomatic malaria parasitaemia (only if participant is symptomatic);
- asymptomatic malaria parasitaemia;
- malaria parasite genotyping;
- Immunology:
    - humoral immune responses to RH5;

Day 168 (± 7days; Post-Vaccination with ChAd63 RH5)

Medical history, temperature monitoring and physical examination will be performed and recorded in the CRF. AEs will be recorded in the CRF.

Blood sample will be collected for the analysis of:

- haematology (full blood count);
- biochemistry (including creatinine and ALT);
- Immunology:
    - humoral immune responses to RH5;
    - cellular immune responses to RH5;
    - transcriptome analysis;
- symptomatic malaria parasitaemia (only if participant is symptomatic);
- asymptomatic malaria parasitaemia;
- malaria parasite genotyping;

##### 11.6. **Unscheduled Visits:**

Participants will be advised to seek first line health care at CTF where they were enrolled during working hours and at BDH outside the working hours. The CTF and BDH will be staffed by clinically-qualified health workers who are on call 24 hours a day, 7 days a week. If needed participants will be referred, and if urgently needed, transported to Muhimbili National Hospital. In the event the unscheduled visit occurs in another health facility then whenever possible, clinical notes related to the management of that event will be retrieved.

Participants will also be informed that community health workers will be readily available to facilitate access to care during the course of the trial. CHW will have access to a study clinician for consultation by mobile phone 24 hours a day and 7 days a week. Reimbursements for transport to CTF or BDH will be provided when participants present for unscheduled visits.

#### 11.7. Sample Handling

Samples collected during the study will be used to address the study objectives and obtain a better understanding of vaccine immune responses following immunization with RH5. Samples may be used to make monoclonal antibodies for analysis related to this vaccine. It is possible that these antibodies may be developed into a product in the future. The participant may withdraw permission for future use of specimens at any time. If a participant withdraws his or her permission for future use of specimens, the Investigator or designee will destroy all known remaining specimens and report this destruction to the participant and the IRB. This decision will not affect the participant’s participation in this protocol or any protocols supported by Ifakara Health Institute.

All samples stored will be labelled with the participant’s study identification (ID) number, which cannot identify the study participant directly but is linkable to other research databases (e.g., questionnaires, clinical assessments, logbooks) generated by the main study. The subject identification log linking the study participant ID number to the name of the participant will be maintained with access limited to authorized research team members. In the event of samples being requested in the future, only the PIs or study coordinator(s) will have access to the log linking the study participant to the samples.

At the completion of the clinical study protocol and the primary and secondary immunology studies described in this protocol, remaining samples will either be destroyed or stored indefinitely (Section 17.9).

In more detail: for samples held at IHI in Tanzania, samples from participants whose parents/guardians or themselves did not provide permission to store them will be discarded after the primary and secondary analyses described in this protocol have been completed. Samples will also be destroyed if the study participants withdraw their permission to store the samples for future studies during the trial. If there is no withdrawal all remaining samples in Tanzania will be stored for future use. Any additional immunological analyses on these samples will be limited to malaria vaccine development unless specific permission for additional studies is obtained from the relevant IRBs. Transfer to another protocol will require approval from the IRB. In the future, other investigators (both at IHI and outside) may wish to study these samples and/or data. In that case, IRB approval must be sought prior to any sharing of samples.

Any clinical information shared about the sample with or without participant identifiers would similarly require prior IRB approval.

The samples exported to UK will be stored at the Jenner Institute, University of Oxford and used for analyses as outlined in the protocol during the study period. However, once the ethical approval on the protocol expires (1 year after the last subject last visit) the samples will be either destroyed or will be transferred to the Oxford Vaccine Centre (OVC) Biobank (only in the cases where consent to store the samples indefinitely has been provided by the participant). The OVC Biobank will then need to give approval for any samples to be extracted and used (so applications directly to the OVC Biobank will need to be made to access these samples).

Blood samples (serum, plasma, whole blood and PBMC) will be stored at IHI BRTC Laboratory in Bagamoyo, Tanzania and some will be shipped to the following collaborative laboratories;

- Jenner Institute laboratories, Centre for Clinical Vaccinology & Tropical Medicine, University of Oxford, Churchill Hospital, Old Road, Oxford, OX3 7LJ, United Kingdom.
- NIH/NIAID Laboratory of Malaria and Vector Research, Malaria Immunology Section, GIA Reference Centre, 12735 Twinbrook Parkway, Twinbrook III, Room 3W-13, Rockville, MD 20852, USA.

At BRTC laboratory, freezer and refrigerator temperatures are recorded twice a day (in the morning and evening), through in-built system and thermometers. The temperature of the freezers will be recorded on temperature logs; in addition, temperature is also continuously recorded by a data logger.

#### 11.8. Laboratory Evaluations

*Safety Blood Assessments*

Blood samples will be collected at specific time-points for haematological and biochemistry tests. The time points and the specific tests are shown in Appendix 1. Protocol specific tests will include the following;

- Complete Blood Count:
  Red blood count, haemoglobin, platelets, white blood count with differential (neutrophil, lymphocyte and eosinophil) counts.
- Biochemistry parameters:
- *At screening:* ALT and creatinine and glucose (random).
- *During subsequent scheduled visits:* ALT and creatinine.

During unscheduled visits, laboratory parameters to be tested will depend on the relevance as determined by the study clinician.

*Urinalysis*:

Urine sample will be collected at screening for urinalysis.

*Malaria Microscopy:*

The WHO malaria microscopy method will be adopted and used to identify (thick blood smear) and quantify (thin blood smear) malaria parasites in participants. Microscopy will be done at screening (before ChAd63 RH5), and monthly thereafter to monitor concurrent malaria infections during the study, and whenever participant presents with malaria symptoms.

*Serology*

Blood samples for HIV, HBV and HCV serology will be done at the screening visit 1 and all individuals infected with any one of these viruses will be excluded from the study.

*Urine Sample Pregnancy Test*

For female adult participants (18-35 years), a urine pregnancy test will be performed at first screening and before each vaccination. Pregnant women will be excluded from participation. If pregnancy is diagnosed after vaccination, the participant will be excluded from further vaccination but will be followed up for safety until the end of the study. If possible, the outcome of the pregnancy will also be determined, including the health of the new-born.

##### 11.9. Rationale for blood volume collected

As part of the process to address the research question, blood samples will be collected to assess both the safety and immunogenicity of RH5 candidate malaria vaccine. It is however imperative to prioritise the safety and wellbeing of the study participants during the study as stipulated in the Declaration of Helsinki. In order to minimize risk to the study participants, the amount of blood volume that will be collected has been estimated using existing guidelines which recommend age-specific volume limits based on total blood volume, haemoglobin level and general health status of the donors [108]. The blood volume limits as recommended by WHO range from 1-5% of the total volume within 24 hours and up to 10% of total blood volume over 8 weeks[109]. A mean of 4% of TBV has been selected as the maximum limit in this trial. This is equivalent to around 3.2mL/Kg. A mean weight for each age group was estimated using the maximum and minimum normal weight limit at –2SD of WHO growth charts and a standard 70kg for young adults. Table 2 show the estimated maximum allowable blood volume per visit and the maximum amount of the blood volume that will be collected.

Table 2: Maximum allowable blood volumes to be collected

| Age group | Mean weight (Kg) | Maximum allowable blood volume per visit | Maximum blood volume to be collected per visit |
|---|---|---|---|
| Infants (6-11 months) | 6.5 | 21mL | 12 mL |
| Young children (1-6 years) | 9 | 29mL | 22mL |
| Adults (18-35 years) | 70 | 140mL | 102mL |

For infants, a lower volume will be taken to minimize the parents/guardian anxiety. Only health participants will be included in the study. This will ensure all participants have normal haemoglobin levels.

#### 11.10. Research Assays to Address Secondary endpoints

*Humoral Immunity*

The blood volumes for the immunological outcomes and time points at which samples will be collected will depend on the group of the participants as shown in Appendix 1. Serum samples collected will be assessed by ELISA for antibodies to RH5 antigen.

Growth inhibition activity (GIA) against a panel of *Pf* isolates and lines to determine antibody function against merozoite invasion of erythrocytes will be performed. Serum IgG subclasses and avidity will be assessed by ELISA, and linear epitopes mapped by peptide arrays. Single-cell analysis of RH5-specific B cells will allow for production of human monoclonal antibodies to investigate epitope specificities. Surface plasmon resonance will be used to assess antibody affinity to RH5 and quantify the concentration of RH5 specific IgG.

*Cellular Immunity*

The blood volumes for cellular immunity and time points at which samples will be collected will differ between the different age groups as shown in Appendix 1. PBMC will be stored immediately after separation at -70ºC and then transferred into liquid nitrogen vapour phase. Stored samples will be used after the study end to address if subsets of T cells are activated by vaccination and show vaccine specific responses.

PBMC will be assessed for vaccine induced cellular immunity by the following assays:

- Intracellular cytokine staining by flow cytometry after stimulation with RH5 peptides.
- *Ex-vivo* ELISPOT after stimulation with RH5 peptides to identify numbers of vaccine induced $CD4^+$ and $CD8^+$ T cells that may be associated with qualitative or quantitative antibody responses. ELISPOT assays may also be used to assess antibody-secreting cell (ASC) and/or memory B cell (mBC) responses to RH5.

All analyses will be conducted at Bagamoyo Research and Training Centre with the exception of GIA and B cell epitope analyses. These will be conducted at The Jenner Institute laboratory in the UK.

### 12. INVESTIGATIONAL PRODUCT (IP)

#### 12.1. IP Description

*ChAd63 RH5*

ChAd63 RH5 was manufactured under Good Manufacturing Practice conditions by Advent in Rome, Italy and shipped to Clinical Biomanufacturing Facility (CBF), Churchill Hospital in Oxford, UK for labelling and release. ChAd63 RH5 is supplied as a sterile 0.65 mL liquid in 2.0 mL glass vials. The dose of ChAd63 RH5 used in this CT is $1 \times 10^{10}$ and $5 \times 10^{10}$ vp, administered intramuscularly.

*MVA RH5*

MVA RH5 was manufactured under Good Manufacturing Practice conditions by Impfstoffwerke DessauTornau (IDT), Germany. MVA RH5 is supplied as a sterile 0.55 mL liquid in 2.0 mL clear glass injection vials. The dose of MVA RH5 to be used in this study is $1 \times 10^8$ and $2 \times 10^8$ pfu, administered intramuscularly.

Final batch certification of the vaccines and associated labelling is performed by the Qualified Person (QP) at the University of Oxford’s Clinical Biomanufacturing Facility (CBF) in accordance with Annex 13 and the CBFs internal SOPs.

#### 12.2. Storage of IP

The vaccine vials are stored between –70°C and –90°C in a locked freezer at the University of Oxford, Churchill Hospital. The vaccines will be shipped from Oxford on dry ice, and then stored in a –70°C freezer at the BRTC laboratory until required. Vaccine accountability, storage, shipment and handling will be in accordance with relevant local standard operating procedures (SOPs) and forms. All movement of vaccines will be documented in vaccine accountability logs according to local site SOPs.

Vaccines will be stored in –70°C freezer at BRTC and will be transported to CTF on dry ice on the morning of use. They will be thawed at CTF and kept in a cold box, and used within 4 hours of thawing.

The IP will be handled according to the relevant SOPs. ChAd63 RH5 and MVA RH5 are genetically modified organisms. In order to minimise dissemination of the recombinant vectored vaccine viruses into the environment, the inoculation site will be covered with a dressing after vaccination. This should absorb any virus that may leak out through the needle track, and will be removed from the injection site after 30 minutes. Vaccine administrators and destructors will follow precautions for the safe handling of GMOs (including the use of eye protection and gloves).

#### 12.3. Packaging and Labelling

The Jenner Institute will supply ChAd63 RH5 and MVA RH5 vaccines to the study site. Control vaccine (rabies vaccine) will be sourced from within Tanzania. An example of labels for ChAd63 RH5 and MVA RH5 is shown in Figure xxx. This is only an example and general format, but the final labels are subject to change.

| | |
|---|---|
| CLINICAL TRIAL: XXX<br>**MVA RH5 VACCINE XX mL**<br>Vial contains XX x $10^8$ pfu/mL<br>Vial No: _______For Intramuscular Injection<br>Batch number: XXXXX Store at -70?C<br>Volunteer number _____<br>Expiry Date: ____________________ | **FOR CLINICAL TRIAL USE ONLY**<br>Sponsor: University of Oxford<br>CCVTM, Old Road, Oxford. OX3 7LJ<br>Tel: 01865 857382<br>Fax: 01865 857471 |

| | |
|---|---|
| CLINICAL TRIAL: XXX<br>**AdCh63 RH5 VACCINE XX mL**<br>Vial contains XX x $10^{11}$ vp/mL<br>Vial No: _______For Intramuscular Injection<br>Batch number: XXXXX Store at -70?C<br>Volunteer number _____<br>Expiry Date: ____________________ | **FOR CLINICAL TRIAL USE ONLY**<br>Sponsor: University of Oxford<br>CCVTM, Old Road, Oxford. OX3 7LJ<br>Tel: 01865 857382<br>Fax: 01865 857471 |

*Figure 8: Example of Vaccine labels*

#### 12.4. Accountability of the Investigational Product

At all times the figures on supplied, used and remaining vaccine doses should match. At the end of the study, it must be possible to reconcile delivery records with those of used and unused stocks. An explanation must be given of any discrepancies. The study pharmacist will ensure accurate records are maintained with regard to the date the vaccine is received, manufacture date, lot numbers, quantity received and disposition of the vaccine according to the relevant SOP. The Sponsor (Oxford University) will also maintain a copy of the records held at the site including Study ID numbers, date and time the IP has been administered, lot number and signature of the person administering the IP. All information that could unblind the observer (clinical team) will be kept in a confidential place by the pharmacist. The clinical team will not have access to the vaccine preparation room during vaccination to avoid the potential for unblinding. All unused vaccines will be returned to Jenner Institute at the end of the trial, or destroyed if requested.

#### 12.5. Administration of the IP

ChAd63 RH5, MVA RH5 and rabies vaccine will all be administered intramuscularly into the left deltoid area according to study specific SOP. Each participant will be monitored for one hour (or longer if necessary) after each vaccination. Resuscitation equipment and medication will be available in the clinic site. Staff trained in Advanced Cardiac Life Support and Paediatric Advanced Life Support will be present at all times.

#### 12.6. Concomitant Medication

Collection of concomitant medications will be done at each visit throughout the immunization period until 30 days after the MVA RH5 vaccination. Participants will be assessed by the study clinician and medication provided in the event of adverse events. The treatment will follow the Tanzania Ministry of Health, World Health Organization and/or international guidelines.

### 13. SAFETY REPORTING

#### 13.1. Definitions

*Adverse Event*

Any untoward medical occurrence in a patient or clinical investigation subject administered a pharmaceutical product and which does not necessarily have a causal relationship with this treatment. An adverse event (AE) can therefore be any unfavourable and unintended sign (including an abnormal laboratory finding), symptom, or disease temporally associated with the use of a medicinal (investigational) product, whether or not related to the medicinal (investigational) product.

Anticipated day-to-day fluctuations of pre-existing conditions that do not represent a clinically significant exacerbation will not be considered adverse events.

Discrete episodes of chronic conditions occurring during a study period will be reported as adverse events in order to assess changes in frequency or severity.

Adverse events will be documented in terms of a medical diagnosis. When this is not possible, the adverse event will be documented in terms of signs and symptoms observed by the Investigator at each study visit.

Pre-existing conditions or signs and/or symptoms (including any which are not recognised at study entry but are recognised during the study period) present in a subject prior to the start of the study will be recorded on the Medical History form within the subject's CRF.

*Adverse Reaction (AR)*

An AR is any untoward or unintended response to an IP. This means that a causal relationship between the IP and an AE is at least a reasonable possibility, i.e., the relationship cannot be ruled out. All cases judged by the reporting medical Investigator as having a reasonable suspected causal relationship to an IP (i.e. possibly, probably or definitely related to an IP) will qualify as adverse reactions.

*Unexpected Adverse Reaction*

An adverse reaction, the nature or severity of which is not consistent with the applicable product information (e.g., IB for an unapproved IP).

*Serious Adverse Event (SAE)*

A serious adverse event (experience) or reaction is any untoward medical occurrence that at any dose:

- results in death,

- is life-threatening, Note: the term "life-threatening" in the definition of "serious" refers to an event in which the patient was at risk of death at the time of the event; it does not refer to an event which hypothetically might have caused death if it were more severe.
- requires inpatient hospitalisation or prolongation of existing hospitalisation,
- results in persistent or significant disability/incapacity, or
- results in a congenital anomaly/birth defect;

*Serious Adverse Reaction (SAR)*

An adverse event (expected or unexpected) that is both serious and, in the opinion of the reporting Investigator or Sponsors, believed to be possibly, probably or definitely due to an IP or any other study treatments, based on the information provided.

*Suspected Unexpected Serious Adverse Reaction (SUSAR)*

A serious adverse reaction, the nature and severity of which is not consistent with the information about the medicinal product in question set out in the IB or Summary of Product Characteristics (SmPC).

Medical and scientific judgment should be exercised in deciding whether expedited reporting is appropriate in other situations, such as important medical events that may not be immediately life-threatening or result in death or hospitalisation but may jeopardise the patient or may require intervention to prevent one of the other outcomes listed in the definition above. These should also usually be considered serious.

##### 13.2. **Procedures for Recording Adverse Events**

*Recording adverse events*

Investigators will evaluate all adverse events observed or reported by the participant or their parents/guardians at each scheduled or unscheduled visit. New adverse events will be recorded in the Adverse Event form within the participant's CRF. Solicited adverse events will be recorded on separate pages of the CRF. The nature of each event, date of onset, outcome, intensity and relationship to vaccination will be established.

Any corrective treatment will be recorded in the CRF as concomitant medication. For solicited adverse events participants or their parents/guardians will be asked direct questions related to the pre-listed symptoms. For unsolicited adverse events, participants or their parents/guardians will be asked an open-ended question such as: "Have you felt different in any way since receiving the vaccine or since the last visit?" The investigator will record only those adverse events having occurred within the time frames defined above.

Adverse events already documented in the CRF, i.e. at a previous assessment and designated as 'ongoing' will be reviewed at subsequent visits, as necessary. If these events have resolved, the documentation in the CRF will be completed, including the date that the adverse event resolved. If an adverse event changes in frequency or intensity during a study period, the record will be updated to reflect the maximum intensity or describe the frequency.

*Solicited Systemic AEs*

For all participants, a prescribed list of systemic AEs will be solicited on the day of vaccination through to 7 days post vaccination. During these periods all participants will be examined for elevated body temperature and allergic reaction (rash, urticaria, pruritus, and/or oedema). Additional AEs occurring in adults (Group 1) will be solicited by history from a list of systemic AEs (nausea, subjective fever, arthralgia, chills, malaise, myalgia, and headache) while the guardians of infants (Group 3) and young children (Group 2) will be asked about observed history of fever, vomiting, diarrhoea, reduced oral intake, and reduced activities (see **Table 3**).

*Solicited Local AEs:*

Pain, limitation of arm movement, pruritus, bruising/ extravasated blood, erythema, swelling, tenderness and induration will be solicited from all participants after each injection. For infants and young children answers to pain, tenderness and pruritus may not be able to be obtained. Local AEs will be solicited on the day of vaccination and 7 days following each vaccination (see **Table 3**).

*Unsolicited AEs*

Unsolicited AEs will be conducted up until day 28 after each vaccination. A syndromic classification will be used for unsolicited AEs, e.g., cough, nasal congestion, sore throat should be combined into upper respiratory tract infection. Thus the term for the unifying diagnosis is recorded as the AE, not each individual sign or symptom whenever applicable.

***Table 3: Solicited Adverse Events***

| | | | | |
|---|---|---|---|---|
| Local Solicited AEs *(at injection site)* | • Pain<br>• Limitation of arm movement | | 0 | No pain |
| | | | 1 | Painful on touch, no restriction on movement of limb (baby cries when limb is touched but moves the limb around without restriction) |
| | | | 2 | Painful when arm is moved (baby cries when limb moved) |
| | | | 3 | Unable to use the limb due to pain |
| | • Pruritus | | 0 | No symptom/sign |
| | | | 1 | Awareness of a symptom, but the symptom is easily tolerated and causes no or minimal interference with usual activity. |
| | | | 2 | Discomfort enough to cause greater than minimal interference with usual activity. |
| | | | 3 | Incapacitating; symptoms causing inability to perform usual activities; requires medical intervention |
| | Infants & children | • Erythema<br>• Swelling<br>• Induration | 0 | 0 |
| | | | 1 | <20 mm |
| | | | 2 | 20-50 mm |
| | | | 3 | >50 mm and/or necrosis or exfoliative dermatitis |
| | Adults | • Erythema<br>• Swelling<br>• Induration | 0 | 0 |
| | | | 1 | <50 mm |
| | | | 2 | 50 – 100 mm |
| | | | 3 | >100 mm and/or necrosis or exfoliative dermatitis |
| Systemic Solicited AEs | • Fever (objective) | | | |
| | | | 1 | 37.6°C – 38.4°C |
| | | | 2 | 38.5°C – 39°C |
| | | | 3 | >39.0°C |
| | Adults | • Allergic reaction *(rash, urticaria, pruritis, oedema)*<br>• Headache<br>• Nausea<br>• Subjective Fever<br>• Fatigue/Malaise<br>• Chills<br>• Myalgia<br>• Arthralgia | 0 | No symptom/sign |
| | | | 1 | Awareness of a symptom, but the symptom is easily tolerated and causes no or minimal interference with usual activity. |
| | Infants and young children | • Allergic reaction *(rash, urticaria, pruritis, oedema)*<br>• Subjective fever<br>• Vomiting<br>• Diarrhoea<br>• Reduced activities<br>• Reduced oral intake | 2 | Discomfort enough to cause greater than minimal interference with usual activity. |
| | | | 3 | Incapacitating; symptoms causing inability to perform usual activities; requires absenteeism or bed rest |

#### *Follow-up of Adverse Events*

All AEs will be followed until resolution of the signs or symptoms or laboratory changes occurs, or until a non-study related causality is assigned.

At the study end, if participants have moderate or severe on-going adverse events not considered to be related to the study vaccine, they will be advised to consult a government health facility. If considered to be related to the study vaccine, a follow-up visit will be arranged to manage the problem and to determine the severity and duration of the event. If appropriate, specialist review will be arranged by Investigators.

Any serious adverse event possibly related to the vaccine and occurring after termination of the clinical trial should be reported by the Investigator according to the procedure described below.

*Grading the severity of adverse events*

For adverse events other than local swelling, redness/discoloration and pain/limitation of limb movement, for which the severity scales are detailed above, AEs will be graded according to the relevant grading tables[110, 111]. These internationally recognized grading tables classify adverse events into one of four grades, ranging from mild to potentially life-threatening. The grading tables have indications for each of over sixty clinical parameters and forty laboratory parameters for grading adult and paediatric AEs. The table also includes general guidelines for estimating the grade of parameters not explicitly listed. Each grade is described broadly below:

- Grade 1 (mild): awareness of a symptom, but the symptom is easily tolerated and causes no or minimal interference with usual activity.
- Grade 2 (moderate): discomfort enough to cause greater than minimal interference with usual activity.
- Grade 3 (severe): incapacitating; symptoms causing inability to perform usual activities; requires absenteeism or bed rest.
- Grade 4 (potentially life-threatening): symptoms causing inability to perform basic self-care functions OR medical or operative intervention is indicated to prevent permanent impairment, persistent disability or death.

Laboratory tests will also be graded based on the relevant toxicity grading scales relevant to healthy HIV negative volunteers with modification to suit local or regional normal values. In addition to the referenced toxicity grading scales above, any other relevant toxicity grading scale may be used to supplement and accomplish proper assessment. Toxicity grades and reference scales which will be used to grade severity of laboratory solicited AEs for this trial will be described in a relevant study SOPs. The abnormal values will be categorized as either clinically significant or non-clinically significant based on the study physician’s medical judgement. Abnormal laboratory assessments that are judged by the Investigator to be serious will be recorded as SAEs.

##### 13.3. **Causality**

The causal relationship between the AE and the product will be evaluated by the PI. This interpretation will be based on the type of event, the relationship of the event to the time of vaccine administration, and the known biology of vaccine therapy. The following are guidelines for assessing the causal relationship:

No relationship:

- No temporal relationship to study product; and
- Alternate aetiology (clinical state, environmental or other interventions); and
- Does not follow known pattern of response to study product

Unlikely relationship

- Unlikely temporal relationship to study product; and
- Alternate aetiology likely (clinical state, environmental or other interventions); and
- Does not follow known typical or plausible pattern of response to study product

Possible relationship:

- Reasonable temporal relationship to study product; or
- Event not readily produced by clinical state, environmental or other interventions; or
- Similar pattern of response to that seen with other vaccines

Probable relationship:

- Reasonable temporal relationship to study product; and
- Event not readily produced by clinical state, environment, or other interventions or
- Known pattern of response seen with other vaccines

Definite relationship:

- Reasonable temporal relationship to study product; and

- Event not readily produced by clinical state, environment, or other interventions; and
- Known pattern of response seen with other vaccines

When a regulatory authority requests distinct classification of AEs into either related or unrelated, without intermediate categories, only "not related" will be regarded as "unrelated" and "unlikely related", "possibly related," "probably related," and "definitely related" will be combined as "related".

#### 13.4. Reporting Procedures for Serious Adverse Events

In order to comply with current regulations on serious adverse event reporting to regulatory authorities, the event will be documented accurately and notification deadlines respected. SAEs will be reported on the standard SAE forms to members of the study team immediately when the investigators become aware of their occurrence. Copies of all reports will be forwarded by email for review to the Chief Investigator (as the Sponsor's representative) within 24 hours of the Investigator being aware of the suspected SAE. The ISMs forming SMC will be notified immediately by the PI if SAEs are deemed possibly, probably or definitely related to study interventions; in such a situation, PI will notify the SMC immediately within 24 hours of the PI being aware of their occurrence.

SAEs will not normally be reported immediately to the OxTREC unless there is a clinically important increase in occurrence rate, an unexpected outcome, or a new event that is likely to affect safety of trial volunteers, at the discretion of the Chief Investigator and/or SMC. Sponsor will notify The Oxford Tropical Research Ethics Committee (OxTREC) of all SAE that are deemed possibly, probably or definitely related to study interventions and SUSARs within specified time-frame as required by the regulations.

All SAEs will be summarized and reported to IHI IRB and The National Health Research Ethics Sub-Committee (NatHREC) in the required specified time-frames. The PI, on behalf of the Sponsor, will also report all SAEs to the Tanzanian Food and Drug Administration within specified time-frame as required by the regulations.

Additional or follow-up information (outcome, precise description of medical history, results of the investigation, copy of hospitalisation report, etc.) relating to the initial SAE report will also be reported to the Sponsor and SMC within 24 hours of receipt of such information.

*Reporting Procedures for SUSARS*

The Chief Investigator (on behalf of The Sponsor) will report all SUSARs to the ethical committee(s) and Tanzanian regulatory authorities within required timelines. The Chief Investigator will also inform all Investigators concerned of relevant information about SUSARs that could adversely affect the safety of participants.

All SUSARs and deaths occurring during the study will be reported to the Sponsor. For all deaths, available autopsy reports and relevant medical reports will be made available for reporting to the relevant authorities.

##### 13.5. Safety monitoring Committee

For this study, an independent SMC will be appointed by the UOXF (Sponsor) to follow up the safety of participants as described above and in section 9.7. The SMC will be composed of 3 members one of whom one will be the Local Safety Monitor (LSM). A local Safety Monitor will be a qualified Tanzanian Paediatrician. All the members will be experienced clinicians qualified to evaluate safety data from clinical studies. The SMC will be responsible for reviewing the safety results during age de-escalation/dose escalation and to provide clearance to progress to the next vaccinations They will also take part in the assessment of adverse events (if requested) and any recommendation regarding halting further vaccination. The SMC will liaise closely with the PI throughout the course, mutually relaying relevant safety information. Safety monitoring charter will be developed that will states their responsibility and provide guidance on the procedures for reviewing the safety data.

In addition to the scheduled reviews, SMC may be contacted for advice and independent review by the Sponsor, CI or PI, as in the following situations:

- AEs in the sentinel participants trigger criteria for an ad hoc SMC review
- Following any SAE deemed to be possibly, probably, or definitely related to the study vaccine.
- Any other situation where the PI, CI or Sponsor thinks independent advice or review is important.

SMC will support the review of safety during scheduled and ad hoc reviews. Only SAE deemed possibly, probably or definitely related to study interventions will be reported to the SMC by the PI. If necessary Sponsor may notify SMC of unrelated SAE in case they need their opinion of the SAE. If required, the SMC may provide a separate written report to document his/her assessment of the SAE or other concerning AE. The occurrence of a SAE possibly related to RH5 will lead to suspension of the trial pending further review of the event by the SMC, as well as the IHI IRB, Oxford Tropical Research Ethics Committee (OxTREC), National Health Research Ethics Sub-Committee (NatHREC) and TFDA.

#### 14. STATISTICAL ANALYSIS CONSIDERATION

Data entry will be performed on site at the BRTC. All final analyses will be performed by the PI with the support from the Sponsor and IHI statisticians.

##### 14.1. Safety Cohort

The 'Safety Cohort' will consist of all participants who have received at least ChAd63 RH5 for whom any data on safety are available. The presentation of safety data will explore separately the adverse experiences among participants who have received both ChAd63 RH5 and MVA RH5 and among those who have received ChAd63 RH5 only.

##### 14.2. Immunogenicity Cohort

The 'Immunogenicity Cohort' will include all evaluable participants (i.e., those meeting all eligibility criteria, and who have received at least ChAd63 RH5 vaccination for whom data concerning immunogenicity endpoint measures are available). This will include participants for whom assay results are available for anti-RH5 antibodies after vaccination.

##### 14.3. Sample Size Considerations

The trial is designed to only detect very large differences in the incidence of local and generalized adverse events between the vaccination groups. This is done to balance the chance to detect any possible untoward reactions against the desire to limit the number of volunteers involved for safety purposes. The sample size of 9-18 per group is reasonable given that studies of ChAd63 MVA with ME-TRAP used between 6 and 30 participant per group in a study that found the vaccine to be safe and well tolerated [99].

##### 14.4. Final Analysis Plan

A final research analysis plan will be prepared by unblended investigator in advance to guide the statistical analysis and reporting of the study. The analysis plan will be agreed upon by the PI and Sponsor prior to locking of the database for the final analysis. The analysis plan will provide information of the populations that will be analyzed and the safety and immunogenicity parameters that will be evaluated as well as specific statistical methods that will be applied. The plan will be applied to the data and sample collected until day 168 post first vaccination for each group. .

##### 14.5. Analysis of Demographics

Demographic characteristics (age, gender) and baseline covariates of each vaccination group and entire study cohort will be tabulated. The mean age (plus range and standard deviation) and gender proportion of the enrolled participants, as a whole, and per vaccination groups, will be calculated.

#### 14.6. Analysis of Safety

Data analysis will consist primarily of descriptive summaries for vaccination groups. The overall percentage of volunteers with at least one local adverse event (solicited or unsolicited) and the percentage with at least one general adverse event (solicited and unsolicited) during the surveillance period after each vaccination will be tabulated. Comparisons between study groups will be made based on a two-sided Fisher's Exact Tests.

Due to the small number of volunteers in this study, all volunteers receiving the same dose of a given vaccine will be pooled for analysis.

#### 14.7. Analysis of Clinical Laboratory Parameters

Haematological and biochemical laboratory parameters will be measured at specific time-points. Clinically relevant abnormal values will be tabulated and a trend analysis could be performed if deemed necessary. Where appropriate highly skewed data will be log-transformed and presented as geometric means with 95% confidence intervals.

#### 14.8. Analysis of immunological endpoints

The primary analysis will be based on the immunogenicity cohort. The primary immunogenicity endpoint for humoral immune responses will be the concentration of anti-RH5 serum antibodies, their percentage GIA *in vitro* using purified IgG and avidity. For cellular immune responses frequency of T-cells specific to RH5 will be identified as $CD4^+$ or $CD8^+$ T-cells expressing cytokines (e.g. IFN-γ) upon *in vitro* stimulation with RH5 antigen and/or RH5-specific ASC or mBC B cell responses.

Descriptive summaries and plot distributions of cellular and humoral immune responses over time for each participant and vaccination group will be presented. The results will be presented both as raw data and as log-transformed data. Between and within groups comparisons will be performed by non-parametric Wilcoxon Rank Sum tests and Wilcoxon Signed Rank test respectively.

### 15. DATA HANDLING AND RECORD KEEPING

#### 15.1. Data handling and record keeping

The PI will be the data manager with responsibility for delegating the receiving, entering, cleaning, querying, analysing and storing all data that accrues from the study. Trained study staff will enter the data into the participants' CRFs, which will be in a paper format. These data will include clinical and laboratory safety data. All source documents and laboratory reports will be reviewed by the clinical team and data entry staff, who will ensure that they are accurate and complete. Adverse events must be graded, assessed for severity and causality, and reviewed by the PI or designee.

The Investigators will maintain appropriate medical and research records for this trial in compliance with ICH E6 GCP and regulatory and institutional requirements for the protection of confidentiality of participants. The Investigators will permit authorized representatives of the Sponsor, regulatory agencies and the monitors to examine clinical records for the purposes of quality assurance reviews, audits and evaluation of the study safety and progress.

Data and sample collected will be provided to the Sponsor to allow study related documentation and immunological analyses where these analyses cannot be done in Tanzania. Data and sample shipment agreements will be established between the Sponsor and the IHI before samples and data are transferred to the Sponsor.

Study documents will be retained for a minimum of 20 years after the end of the clinical trial. These documents will be retained for a longer period, however, if required by local regulations. No records will be destroyed without the written consent of Sponsor and it is the responsibility of Sponsor to inform the investigator when these documents no longer need to be retained.

#### 15.2. Access to Data

Direct access will be granted to authorised representatives from the Sponsor, host institution and the ethical and regulatory authorities to permit trial-related monitoring, audits and inspections and evaluation of the study safety, progress, and data validity. All documents will be stored safely in a locked filing cabinet where only study Investigators will have access to the keys.

#### 15.3. Source document and Case Report Form

All protocol-required information will be collected in CRFs designed by the investigator. All source documents will be filed in the CRF. Source documents are original documents, data, and records from which the volunteer's CRF data are obtained. For this study these will include, but are not limited to; volunteer consent form, blood results, clinician's medical notes, laboratory records, and relevant correspondences. In the majority of cases, CRF entries will be considered source data as the CRF is the site of the original recording (i.e. there is no other written or electronic record of data). In this study collected information in the CRF will include, but is not limited to medical history, medication records, vital signs, physical examination records, urine assessments, blood, urine analysis results, adverse event data and details of vaccinations. If participants fell ill and receive medical treatment, medical notes and investigational results will also be considered as source documents. All source data and participant CRFs will be stored in a secure place accessible to only authorized study staff.

Data entry into the 21 CFR Part 11-compliant Electronic Data Capture system will be done by study staff. The data system will include password protection and internal quality checks, such as automatic range checks, to identify data that appear inconsistent, incomplete, or inaccurate. Clinical data will be entered directly from the source documents. Immunology data will be generated from analyses of blood samples and will be stored in Excel format for sharing and archiving, and analysed using statistical packages (Prism, and/or STATA). The name and any other identifying detail will NOT be included in any trial data electronic file. On all trial-specific documents, other than the signed consent, the participant will be referred to by the trial Study ID number, not by name.

### 16. QUALITY ASSURANCE PROCEDURES

The trial will be conducted in accordance with the current approved protocol, GCP, relevant regulations and standard operating procedures.

#### 16.1. Monitoring

Clinical trial monitoring will be conducted by IHI QA team according to applicable local SOP to ensure that GCP standards and regulatory guidelines are being followed. Pre-trial monitoring visits will be made to the site, including the clinical laboratory. All records will be made available to monitors, including regulatory files, CRFs and other source documents, QA/QC documentation, SOPs, etc.

A detailed monitoring plan, subject to approval by the sponsor, will be developed by IHI QA team. The monitoring plan will include the number of participant charts to be reviewed, which/what proportion of data fields and what will be monitored, and who will be responsible for conducting the monitoring visits, and who will be responsible for ensuring that monitoring findings are addressed.

#### 16.2. Investigator procedures

Approved site-specific SOPs will be used at all clinical and laboratory sites.

#### 16.3. Modification to protocol

No amendments to this protocol will be made without consultation with, and agreement of, the Sponsor. Any amendments to the trial that appear necessary during the course of the trial must be discussed by the Investigator and Sponsor concurrently. If agreement is reached concerning the need for an amendment, it will be produced in writing by the PI and will be made a formal part of the protocol following ethical and regulatory approval.

An administrative change to the protocol is one that modifies administrative and logistical aspects of a protocol but does not affect the subjects' safety, the objectives of the trial and its progress. An administrative change does not require IRB or regulatory approval.

The Investigator is responsible for ensuring that changes to an approved trial, during the period for which regulatory and IRB approval has already been given, are not initiated without regulatory and IRB review and approval except to eliminate apparent immediate hazards to the subject.

#### 16.4. Protocol deviation

A protocol deviation is any noncompliance with the clinical trial protocol, GCP, or SOP requirements. The noncompliance may be either on the part of the participant, the investigator, or the study site staff. As a result of deviations, corrective actions are to be developed by the site and implemented promptly.

It is the responsibility of the site to use continuous vigilance to identify and report deviations within 5 working days of identification of the protocol deviation, or within 5 working days of the planned protocol-required activity. All deviations must be promptly reported to the sponsor.

Any deviations that impact subject safety, or that alter the risk: benefit analysis or the scientific integrity of the study is regarded as major deviation and will be reported to the Sponsor within 24 hours of the PI or study personnel becoming aware of the deviation. Major deviation will be reported to the Tanzanian regulatory authorities by the PI within 7 days of the Sponsor becoming aware of the deviation.

All deviations from the protocol must be addressed in study participant source documents. A completed copy of the Protocol Deviation Form must be maintained in the trial master file, as well as in the participant's source document. Protocol deviations will be reported to the IRB's per their guidelines.

##### 16.5. Audit & inspection

The QA designated site staff will conduct internal audits to check that the trial is being conducted, data recorded, analysed and accurately reported according to the protocol, sponsor's SOPs and in compliance with ICH GCP. The audits will also include laboratory activities according to an agreed audit schedule.

The Sponsor may carry out audit to ensure compliance with the protocol, GCP and appropriate regulations. GCP inspections may also be undertaken by the regulatory authority to ensure compliance with protocol and national regulations. The sponsor will assist in any inspections.

##### 16.6. Trial Progress

The progress of the trial will be overseen by the Principal Investigator

##### 16.7. Study results feedback

Study outcomes will be shared with study participants after the final safety report is completed. In addition local authorities will be informed of the end of the study. Final safety report will be submitted to all IRBs and regulatory authority.

### 17. ETHICAL AND REGULATORY CONSIDERATIONS

#### 17.1. Declaration of Helsinki

The Investigator will ensure that this trial is conducted in accordance with the principles of the Declaration of Helsinki 2008.

#### 17.2. Guidelines for Good Clinical Practice

The Investigator will ensure that this trial is conducted in accordance with Medicine for Human use (clinical trials) Regulations 2004 and its amendments and with the ICH guidelines for GCP (CPMP/ICH/135/95) July 1996. The trial will also comply with the European Communities (Clinical Trials on Medicinal Products for Human Use) Regulations, 2004 [S.I. 190 of 2004].

#### 17.3. Approvals

The protocol, informed consent form, participant information sheet and any proposed advertising material will be submitted to IHI IRB, National Health Research Ethics Sub-Committee (NatHREC) and Oxford Tropical Research Ethics Committee (OxTREC) for written approval. In addition approval from Tanzania Drug and Food Authority (TFDA), a regulatory authority, will be sought before the study starts. No data collection will start without approval from ethics committees and regulatory authority.

The Investigator will submit and, where necessary, obtain approval from the above parties for all substantial amendments to the original approved documents.

#### 17.4. Reporting

The PI shall submit once a year throughout the clinical trial, or on request, an Annual Progress Report to the IHI IRB, National Health Research Ethics Sub-Committee (NatHREC), Oxford Tropical Research Ethics Committee (OxTREC) and Sponsor. In addition, an End of Trial notification and final report will be submitted to the IHI IRB, National Health Research Ethics Sub-Committee (NatHREC), Oxford Tropical Research Ethics Committee (OxTREC) and Sponsor.

#### 17.5. Participant Confidentiality

The trial staff will ensure that the participants' anonymity is maintained. The participants will be identified only by a participant study ID on all trial documents and any electronic database, with the exception of the CRF, where participant initials may be added. All documents will be stored securely and only accessible by trial staff and authorised personnel, including representatives of the REC and regulatory authorities. The trial will comply with the Data Protection Act, 1998, which requires data to be anonymised as soon as it is practical to do so.

Photographs taken of vaccination sites (if required, with the volunteer's written, informed consent) will not include the volunteer's face and will be identified by the date, trial code and subject's unique identifier. Once developed, photographs will be stored as confidential records, as above. This material may be shown to other professional staff, used for educational purposes, or included in a scientific publication.

##### 17.6. Potential Benefits to participants

Participants will benefit from receiving a free consultation and information about their general health status at screening. This information will help participants get medical attention as soon as possible to avoid potential complications. In addition participants in the control group will receive the rabies vaccine which will protect them against rabies in case they are bitten/scratched by a rabid animal. A stock of rabies vaccine will also be kept to assist participants who received IP in case they are bitten/scratched by a rabid animal. During the trial, all participants will receive information about their health status. Medical costs for acute illnesses whether or not related to the IP or study procedure during the study period will be covered by the study to the limit of the allocated funds. When experiencing illness, enrolled participants will receive care at Bagamoyo District Hospital. If the situation arises in which medical care for a complication unrelated to the study procedure/product exceeds the limits of the budget allocated for clinical care, the participant or guardian will be responsible for funding this care through the normal channels provided by the Ministry of Health and Social Welfare of United Republic of Tanzania.

##### 17.7. Potential Risks and Burden to Participants

*Vaccination:*

Risks for participants are related to exposure to ChAd63 and MVA vectors and RH5 antigens expressed by these vectors. Potential expected risks from vaccination, which include local and systemic reactions are specific to each IP and are described in section 13. It is important to note that ChAd63 RH5 & MVA RH5 have not previously been administered to young children and infants but have been safely administered in UK adults. ChAd63 and MVA expressing ME-TRAP have been safety administered in infants and young children [99]. Therefore, although the AE profile can be estimated from previous use of these vectors, the reactogenicity may vary from that seen previously with ChAd63 and MVA encoding ME-TRAP. For this reason, vaccinees will be enrolled in a staggered format to allow early identification of any concerning reactogenicity before the majority of individuals have been vaccinated. Any vaccine can cause allergic reactions including anaphylaxis. Therefore vaccination will take place in the presence Advanced Life Support trained physicians and where equipment and drugs for managing anaphylaxis and any other severe adverse reaction are available.

*Phlebotomy:*

It is expected that participants will experience pain of a needle during vaccination and blood drawing at different time-points. The maximum volume of blood drawn over the study period will be based on age and weight and is not expected to compromise the health of participants. There may be minor bruising, local tenderness or pre-syncopal symptoms associated with venepuncture.

##### 17.8. **Incentives**

The study will cover all the costs related to participant's participation in the trial including clinic visits or hospitalization. A reasonable time reimbursement of 15000 Tanzanian shillings will be provided during scheduled visits. In addition participants will receive a meal during the scheduled visits. In case of expected or unexpected medical complications, medical treatment at CTF and BDH will be available at no cost to the participant. Participants who come for unscheduled visits will be reimbursed for their transport fare. In the event it is required, additional insurance coverage for the costs of medical treatment for acute illnesses or adverse events related to investigational product will be covered by liability insurance held by Oxford University.

##### 17.9. **Future use of stored samples**

If residual sera and cells are available following the serological and CMI assays described in this protocol, additional immunological and *in vitro* studies may be performed at the Bagamoyo Research and Training Centre laboratory in Tanzania on those samples for which permission was expressly granted for storing the samples for future studies at the time of informed consent. These assays may include ability of participant sera to interfere with *in vitro* parasite growth. Additional research questions to be asked for cells include antigen-specific cytokine induction as measured by ELISPOT, flow cytometry, or both. Samples from participants whose parents/guardians or themselves did not provide permission to store samples will be discarded after the analyses described in the current protocol have been completed. Study participants will have the right to withdraw their permission for further use of their samples at any time during and after the study.

The samples that are sent to UK will be stored at the Jenner Institute and used for analyses as outlined in the protocol. At the end of the study (defined as one year after the last subject last visit), samples will be either destroyed, or transferred to the Oxford Vaccine Centre (OVC) Biobank if the consent to store the sample indefinitely has been granted by the participant. Additional analyses related to malaria vaccine development can be done on these samples after receiving approval from the Biobank.

Maximizing the research use of collected samples from participants to benefit science and society is an important ethical consideration. It is important to make efficient use of participant samples in an ethical manner with respect and transparency rather than collecting new samples [112-114].

Development of a malaria vaccine is an evolving science and it is expected that the samples collected from this trial and stored for future use will be used to analyse the immune mechanisms of RH5 and other candidate malaria vaccines with potential benefit to science and global society. Development of some advanced candidate malaria vaccines such as RTS,S and ME-TRAP has been ongoing for over 40 years and some of the important insights on these antigens were made from the stored samples collected from different clinical trials and immune-epidemiological studies through collaborative research[31, 115, 116].

### 18. FINANCE AND INSURANCE

#### 18.1. Funding

The study will be funded primarily by a grant from the Medical Research Council, UK.

#### 18.2. Insurance

*Negligent Harm:* Indemnity and/or compensation for negligent harm arising specifically from an accidental injury for which Ifakara Health Institute is legally liable will be covered by the study.

*Non-Negligent Harm:* Indemnity and/or compensation for harm arising specifically from an accidental injury, and occurring as a consequence of the Research Subjects' participation in the trial for which the University is the Research Sponsor will be covered by the Ifakara Health Institute and University of Oxford (the University has a specialist insurance policy in place with Newline Underwriting Management Ltd, at Lloyd's of London, UK). In addition compensation for injury will be guided by the respective insurance policies that adhere to the national guidelines provided by the TFDA.

#### 19. PUBLICATION POLICY

The Final Safety Report will be prepared by the PI. The protocol and data derived from the trial are the shared property of the IHI, Oxford University and GSK. Any publication or presentation, abstracts and press releases related to the trial must be approved by all parties' representatives before submission of the manuscript. After publication of the results of the trial, any participating partner may publish or otherwise use its own data provided that any publication of data from the trial gives recognition to the trial group. Either partner must have the opportunity to review the proposed abstract, manuscript or presentation before submission for publication/presentation. A request for delay in publication shall only be allowed to enable the requesting party to secure its proprietary and confidential information contained in the draft publication and/or to prevent the loss of patenting opportunities. Such delay shall not be more than 3 months starting from the day of submission for approval. Any information identified as confidential must be deleted prior to submission. The authorized persons as an author of the publication(s) are those who have contributed to the protocol and/or to the analysis of the data, drafting and reviewing the communication. According to the main topic of the publication, the first author will be the greatest contributing Investigator. Data from the study may also be used as part of a thesis for a PhD, MD or Masters. Publications arising from this study will be made open access.

mt2013248 [pii]. PubMed PMID: 24166483; PubMed Central PMCID: PMC3911893.

mt2012223 [pii]. PubMed PMID: 23089736; PubMed Central PMCID: PMC3519995.
96. Stanisic DI, Gerrard J, Fink J, Griffin PM, Liu XQ, Sundac L, et al. Infectivity of Plasmodium falciparum in Malaria-Naive Individuals Is Related to Knob Expression and Cytoadherence of the Parasite. Infect Immun. 2016;84(9):2689-96. doi: 10.1128/IAI.00414-16. PubMed PMID: 27382019; PubMed Central PMCID: PMC4995910.
97. Vollmar J, Arndtz N, Eckl KM, Thomsen T, Petzold B, Mateo L, et al. Safety and immunogenicity of IMVAMUNE, a promising candidate as a third generation smallpox vaccine. Vaccine. 2006;24(12):2065-70. doi: 10.1016/j.vaccine.2005.11.022. PubMed PMID: 16337719.
98. Kennedy JS, Greenberg RN. IMVAMUNE: modified vaccinia Ankara strain as an attenuated smallpox vaccine. Expert review of vaccines. 2009;8(1):13-24. doi: 10.1586/14760584.8.1.13. PubMed PMID: 19093767.
99. Afolabi MO, Tiono AB, Adetifa UJ, Yaro JB, Drammeh A, Nebie I, et al. Safety and Immunogenicity of ChAd63 and MVA ME-TRAP in West African children and infants. Molecular therapy : the journal of the American Society of Gene Therapy. 2016. doi: 10.1038/mt.2016.83. PubMed PMID: 27109630.
100. Bliss CM, Drammeh A, Bowyer G, Sanou GS, Jagne YJ, Ouedraogo O, et al. Viral Vector Malaria Vaccines Induce High-Level T Cell and Antibody Responses in West African Children and Infants. Molecular therapy : the journal of the American Society of Gene Therapy. 2017;25(2):547-59. doi: 10.1016/j.ymthe.2016.11.003. PubMed PMID: 28153101; PubMed Central PMCID: PMCPMC5368405.
101. Ajjan N, Soulebot JP, Stellmann C, Biron G, Charbonnier C, Triau R, et al. [Results of preventive rabies vaccination with a concentrated vaccine of the PM/WI38-1503-3M rabies strain cultured on human diploid cells. Preparation of mixed antirabies-antitetanus hyperimmune immunoglobulin by plasmapheresis of blood taken from vaccinated veterinary students]. Dev Biol Stand. 1978;40:89-100. PubMed PMID: 680405.
102. Costy-Berger F. [Preventive rabies vaccination using vaccine prepared from human diploid cells]. Dev Biol Stand. 1978;40:101-4. PubMed PMID: 680381.

103. Tanzania Commission for AIDS (TACAIDS) ZACZ, National Bureau of, Statistics (NBS) OotCGSO, and ICF International,. Tanzania HIV/AIDS and Malaria Indicator Survey 2011-12. Dar es Salaam, Tanzania: TACAIDS, ZAC, NBS, OCGS, and ICF International., 2013.
104. Mensah VA, Gueye A, Ndiaye M, Edwards NJ, Wright D, Anagnostou NA, et al. Safety, Immunogenicity and Efficacy of Prime-Boost Vaccination with ChAd63 and MVA Encoding ME-TRAP against Plasmodium falciparum Infection in Adults in Senegal. PloS one. 2016;11(12):e0167951. doi: 10.1371/journal.pone.0167951. PubMed PMID: 27978537; PubMed Central PMCID: PMCPMC5158312 applications and patents on malaria vectored vaccines and immunisation regimes including the following (WO2008/122769, Adenoviral vector encoding malaria antigen; and WO 2008/122811 Novel adenovirus vectors). Egeruan Imoukhuede and Ines Petersen were employees of EVI at the time of the study, which supports the development and testing of malaria vaccines. Nicola Viebig is an employee of EVI and Odile Leroy is executive director of EVI. Authors from ReiThera (formerly Okairos) are employees of and/or shareholders in ReiThera, which is developing vectored vaccines for malaria and other diseases. Alfredo Nicosia was employed by ReiThera (formerly Okairos) at the time of the study. There are no further patents, products in development or marketed products to declare. This does not alter our adherence to all the PLOS ONE policies on sharing data and materials, as detailed online in the guide for authors.
105. Clem AS. Fundamentals of vaccine immunology. J Glob Infect Dis. 2011;3(1):73-8. doi: 10.4103/0974-777X.77299. PubMed PMID: 21572612; PubMed Central PMCID: PMCPMC3068582.
106. Medscape. Yellow fever vaccine 2017 [cited 2017 21 October 2017]. Available from: https://reference.medscape.com/drug/y-f-vax-yellow-fever-vaccine-343179.
107. WHO. International travel and health 2017 [cited 2017 21 October 2017]. Available from: http://www.who.int/ith/vaccines/yf/en/.
108. Gibson BE, Todd A, Roberts I, Pamphilon D, Rodeck C, Bolton-Maggs P, et al. Transfusion guidelines for neonates and older children. Br J Haematol. 2004;124(4):433-53. PubMed PMID: 14984493.
109. Howie SR. Blood sample volumes in child health research: review of safe limits. Bulletin of the World Health Organization. 2011;89(1):46-53. doi: 10.2471/BLT.10.080010. PubMed PMID: 21346890; PubMed Central PMCID: PMCPMC3040020.
110. Services. DoANIoAaIDNIoHUDoHaH. Division of AIDS (DAIDS) Table for Grading the Severity of Adult and Pediatric Adverse Events. Division of AIDS National Institute of Allergy and Infectious Diseases National Institutes of Health US Department of Health and Human Services., 2017 July 2017 Report No.
111. US-FDA. Guidance for Industry: Toxicity Grading Scale for Healthy Adult and Adolescent Volunteers Enrolled in Preventive Vaccine Clinical Trials 2014 [updated 15 September 2014 23 October 2017]. Available from: https://www.fda.gov/biologicsbloodvaccines/guidancecomplianceregulatoryinformation/guidances/vaccines/ucm074775.htm.
112. WHO. PROPOSED INTERNATIONAL GUIDELINES ON ETHICAL ISSUES IN MEDICAL GENETICS AND GENETIC SERVICES: Report of a WHO meeting on Ethical Issues in Medical Genetics. 1998 15-16 December 1997. Report No.: Contract No.: WHO/HGN/GL/ETH/98.1.
113. Human Tissue Act 2004 2004 [cited 2017 23 October 2017]. Available from: http://www.legislation.gov.uk/ukpga/2004/30/contents.
114. Petrini C. "Broad" consent, exceptions to consent and the question of using biological samples for research purposes different from the initial collection purpose. Soc Sci Med. 2010;70(2):217-20. doi: 10.1016/j.socscimed.2009.10.004. PubMed PMID: 19853341.
115. Ballou WR, Cahill CP. Two decades of commitment to malaria vaccine development: GlaxoSmithKline Biologicals. The American journal of tropical medicine and hygiene. 2007;77(6 Suppl):289-95. PubMed PMID: 18165505.
116. Hill AV. Pre-erythrocytic malaria vaccines: towards greater efficacy. Nat Rev Immunol. 2006;6(1):21-32. doi: 10.1038/nri1746. PubMed PMID: 16493425.

### 21. APPENDIX A: SCHEDULE OF PROCEDURES

| Group 1 (18 to 35 years) | | Screening | Vaccination and Post Vacciation Follow-up Period | | | | | | | | | | | | | | | | | | | Long Term Follow-up | | | |
|---|---|---|---|---|---|---|---|---|---|---|---|---|---|---|---|---|---|---|---|---|---|---|---|---|---|
| Study Weeks Relative to First Vaccination | | 0 | 1 | | | | | | | 2 | 3 | 5 | 8 | 8 | | | | | | 9 | 10 | 12 | 16 | 20 | 24 |
| Study Days Relative to First Vaccination | | `-30- to 0 | 0 | 1 | 2 | 3 | 4 | 5 | 6 | 7 | 14 | 28 | 56 | 57 | 58 | 59 | 60 | 61 | 62 | 63 | 70 | 84 | 112 | 140 | 168 |
| Study Visit Code | | SC** | V1* | V1+1 | V1+2 | V1+3 | V1+4 | V1+5 | V1+6 | V1+7 | V1+14 | V1+28 | V2* | V2+1 | V2+2 | V2+3 | V2+4 | V2+5 | V2+6 | V2+7 | V2+14 | V2+28 | V2+56 | V2+84 | V2+112 |
| Window Relative to each Study Visit | | | 0 | - | ±1 | - | - | - | - | -1/+3 | -1/+3 | -7/+2 | ±7d | - | ±1 | - | - | - | - | -1/+3 | -1/+3 | -7/+2 | ±7 | ±7 | ±7 |
| Home Visits | | | | X | | X | X | X | X | | | | | X | | X | X | X | X | | | | | | |
| Clinic Visits | | X | X | | X | | | | | X | X | X | X | | X | | | | | X | X | X | X | X | X |
| Vaccination (ChAd63/MVA-RH5 or Rabies vaccine) | | | X | | | | | | | | | | X | | | | | | | | | | | | |
| **Clinical Assessment** | | | | | | | | | | | | | | | | | | | | | | | | | |
| Informed Consent and/or Eligibility | | X | X | | | | | | | | | | | | | | | | | | | | | | |
| Medical History with demographic data | | X | | | | | | | | | | | | | | | | | | | | | | | |
| Enrolment | | | X | | | | | | | | | | | | | | | | | | | | | | |
| Photo taken and Temporary ID Card | | X | | | | | | | | | | | | | | | | | | | | | | | |
| Permanent ID Card | | | X | | | | | | | | | | | | | | | | | | | | | | |
| Physical Examination | •Detailed | X | | | | | | | | | | | | | | | | | | | | | | | |
| | •Focused | | X | | X | | | | | X | X | X | X | | X | | | | | X | X | X | | X | X |
| Physical Assessments | •Vital Signs | X | X | | X | | | | | X | X | X | X | | X | | | | | X | X | X | X | X | X |
| | •Weight and Height | X | X# | | | | | | | | | | X# | | | | | | | | | | | | X# |
| Adverse Events Data Collection | •Solicited | | X | X | X | X | X | X | X | X | | | X | X | X | X | X | X | X | X | | | | | |
| | •Unsolicited | | X | X | X | X | X | X | X | X | X | X | X | X | X | X | X | X | X | X | X | X | | | |
| | •SAE & New Medical Conditions | | X | X | X | X | X | X | X | X | X | X | X | X | X | X | X | X | X | X | X | X | X | X | X |
| Supply Insecticide Treated Nets (ITN) | | X | | | | | | | | | | | | | | | | | | | | | | | |
| **Laboratory Assessment:** Numbers indicate volume of blood to be collected at each timepoint | | | | | | | | | | | | | | | | | | | | | | | | | |
| Hematology | •RBC, HGB, platelets, WBC with differential (neutrophil, lymphocyte and eosinophil) counts | 0.5 | 0.5 | | | | | | | 0.5 | 0.5 | 0.5 | 0.5 | | | | | | | 0.5 | 0.5 | 0.5 | | | 0.5 |
| Biochemistry | •ALT,Creatinine | 1 | 1 | | | | | | | 1 | 1 | 1 | 1 | | | | | | | 1 | 1 | 1 | | | 1 |
| Parasitology | •Thick blood smear/Parasite Genotyping | 0.5 | | | | | | | | | | | 0.5 | | | | | | | 0.5 | 0.5 | 0.5 | 0.5 | 0.5 | 0.5 |
| Serology [HIV, HBV and HCV] | | 1 | | | | | | | | | | | | | | | | | | | | | | | |
| Immunology | | | 100 | | | | | | | | 70 | 70 | 70 | | | | | | | 100 | | 100 | 50 | 50 | 70 |
| Urine (Pregnancy test)*** | | X | X | | | | | | | | | | X | | | | | | | | | | | | |
| Urine Dipstick [Blood and Protein] | | X | | | | | | | | | | | | | | | | | | | | | | | |
| Total Blood Volume withdrawn/Visit (mL▶ | | 3.0 | 101.5 | 0.0 | 0.0 | 0.0 | 0.0 | 0.0 | 0.0 | 1.5 | 71.5 | 71.5 | 72.0 | 0.0 | 0.0 | 0.0 | 0.0 | 0.0 | 0.0 | 102.0 | 2.0 | 102.0 | 50.5 | 50.5 | 72.0 |
| Cumulative Blood Volume withdrawn (mL▶ | | 3.5 | 105.0 | 105.0 | 105.0 | 105.0 | 105.0 | 105.0 | 105.0 | 106.5 | 178.0 | 249.5 | 321.5 | 321.5 | 321.5 | 321.5 | 321.5 | 321.5 | 321.5 | 423.5 | 425.5 | 527.5 | 578.0 | 628.5 | 700.5 |

* Laboratory tests for this visit must be done within 24 hours before vaccination.

** This includes a screening visit 2 when participant will receive their results

*** Only for women

### Only Weight will be measured

| Group 2 (1 to 6 years) | | | Vaccination and Post Vacciation Follow-up Period | | | | | | | | | | | | | | | | | | | Long Term Follow-up | | | |
|---|---|---|---|---|---|---|---|---|---|---|---|---|---|---|---|---|---|---|---|---|---|---|---|---|---|
| Study Weeks Relative to First Vaccination | | 0 | 1 | | | | | | | 2 | 3 | 5 | 8 | 8 | | | | | | 9 | 10 | 12 | 16 | 20 | 24 |
| Study Days Relative to First Vaccination | | `-30- to 0 | 0 | 1 | 2 | 3 | 4 | 5 | 6 | 7 | 14 | 28 | 56 | 57 | 58 | 59 | 60 | 61 | 62 | 63 | 70 | 84 | 112 | 140 | 168 |
| Study Visit Code | | SC** | V1* | V1+1 | V1+2 | V1+3 | V1+4 | V1+5 | V1+6 | V1+7 | V1+14 | V1+28 | V2* | V2+1 | V2+2 | V2+3 | V2+4 | V2+5 | V2+6 | V2+7 | V2+14 | V2+28 | V2+56 | V2+84 | V2+112 |
| Window Relative to each Study Visit | | | 0 | - | ±1 | - | - | - | - | -1/+3 | -1/+3 | -7/+2 | ±7d | - | ±1 | - | - | - | - | -1/+3 | -1/+3 | -7/+2 | ±7 | ±7 | ±7 |
| Home Visits | | | | X | | X | X | X | X | | | | | X | | X | X | X | X | | | | | | |
| Clinic Visits | | X | X | | X | | | | | X | X | X | X | | X | | | | | X | X | X | X | X | X |
| Vaccination (ChAd63/MVA-RH5 or Rabies vaccine) | | | X | | | | | | | | | | X | | | | | | | | | | | | |
| **Clinical Assessment** | | | | | | | | | | | | | | | | | | | | | | | | | |
| Informed Consent and/or Eligibility | | X | X | | | | | | | | | | | | | | | | | | | | | | |
| Medical History with demographic data | | X | | | | | | | | | | | | | | | | | | | | | | | |
| Enrolment | | | X | | | | | | | | | | | | | | | | | | | | | | |
| Photo taken and Temporary ID Card | | X | | | | | | | | | | | | | | | | | | | | | | | |
| Permanent ID Card | | | X | | | | | | | | | | | | | | | | | | | | | | |
| Physical Examination | •Detailed | X | | | | | | | | | | | | | | | | | | | | | | | |
| | •Focused | | X | | X | | | | | X | X | X | X | | X | | | | | X | X | X | | X | X |
| Physical Assessments | •Vital Signs | X | X | | X | | | | | X | X | X | X | | X | | | | | X | X | X | X | X | X |
| | •Weight and Height | X | X# | | | | | | | | | | X# | | | | | | | | | | | | X# |
| | •MUAC | X | X | | | | | | | | | | X | | | | | | | | | | | | X |
| Adverse Events Data Collection | •Solicited | | X | X | X | X | X | X | X | X | | | X | X | X | X | X | X | X | X | | | | | |
| | •Unsolicited | | X | X | X | X | X | X | X | X | X | X | X | X | X | X | X | X | X | X | X | X | | | |
| | •SAE & New Medical Conditions | | X | X | X | X | X | X | X | X | X | X | X | X | X | X | X | X | X | X | X | X | X | X | X |
| Supply Insecticide Treated Nets (ITN) | | X | | | | | | | | | | | | | | | | | | | | | | | |
| **Laboratory Assessment:** Numbers indicate volume of blood to be collected at each timepoint | | | | | | | | | | | | | | | | | | | | | | | | | |
| Hematology | •RBC, HGB, platelets, WBC with differential (neutrophil, lymphocyte and eosinophil) counts | 0.5 | 0.5 | | | | | | | 0.5 | 0.5 | 0.5 | 0.5 | | | | | | | 0.5 | 0.5 | 0.5 | | | 0.5 |
| Biochemistry | •ALT,Creatinine | 1 | 1 | | | | | | | 1 | 1 | 1 | 1 | | | | | | | 1 | 1 | 1 | | | 1 |
| Parasitology | •Thick blood smear/Paraiste Genotyping | 0.5 | | | | | | | | | | | 0.5 | | | | | | | 0.5 | 0.5 | 0.5 | 0.5 | 0.5 | 0.5 |
| Serology [HIV, HBV and HCV] | | 1 | | | | | | | | | | | | | | | | | | | | | | | |
| Immunology | | | 20 | | | | | | | | 15 | 20 | 20 | | | | | | | 20 | | 20 | 15 | 15 | 20 |
| Urine Dipstick [Blood and Protein] | | X | | | | | | | | | | | | | | | | | | | | | | | |
| Total Blood Volume withdrawn/Visit (mL▶ | | 3.0 | 21.5 | 0.0 | 0.0 | 0.0 | 0.0 | 0.0 | 0.0 | 1.5 | 16.5 | 21.5 | 22.0 | 0.0 | 0.0 | 0.0 | 0.0 | 0.0 | 0.0 | 22.0 | 2.0 | 22.0 | 15.5 | 15.5 | 22.0 |
| Cumulative Blood Volume withdrawn (mL▶ | | 3.5 | 25.0 | 25.0 | 25.0 | 25.0 | 25.0 | 25.0 | 25.0 | 26.5 | 43.0 | 64.5 | 86.5 | 86.5 | 86.5 | 86.5 | 86.5 | 86.5 | 86.5 | 108.5 | 110.5 | 132.5 | 148.0 | 163.5 | 185.5 |

* Laboratory tests for this visit must be done within 24 hours before vaccination.

** This includes a screening visit 2 when participant will receive their results

### Only Weight will be measured

| Group 3 (6 to 11 month) | | | Vaccination and Post Vacciation Follow-up Period | | | | | | | | | | | | | | | | | | | Long Term Follow-up | | | |
|---|---|---|---|---|---|---|---|---|---|---|---|---|---|---|---|---|---|---|---|---|---|---|---|---|---|
| Study Weeks Relative to First Vaccination | | 0 | 1 | | | | | | | 2 | 3 | 5 | 8 | 8 | | | | | | 9 | 10 | 12 | 16 | 20 | 24 |
| Study Days Relative to First Vaccination | | `-30- to 0 | 0 | 1 | 2 | 3 | 4 | 5 | 6 | 7 | 14 | 28 | 56 | 57 | 58 | 59 | 60 | 61 | 62 | 63 | 70 | 84 | 112 | 140 | 168 |
| Study Visit Code | | SC** | V1* | V1+1 | V1+2 | V1+3 | V1+4 | V1+5 | V1+6 | V1+7 | V1+14 | V1+28 | V2* | V2+1 | V2+2 | V2+3 | V2+4 | V2+5 | V2+6 | V2+7 | V2+14 | V2+28 | V2+56 | V2+84 | V2+112 |
| Window Relative to each Study Visit | | | 0 | - | ±1 | - | - | - | - | -1/+3 | -1/+3 | -7/+2 | ±7d | - | ±1 | - | - | - | - | -1/+3 | -1/+3 | -7/+2 | ±7 | ±7 | ±7 |
| Home Visits | | | | X | | X | X | X | X | | | | | X | | X | X | X | X | | | | | | |
| Clinic Visits | | X | X | | X | | | | | X | X | X | X | | X | | | | | X | X | X | X | X | X |
| Vaccination (ChAd63/MVA-RH5 or Rabies vaccine) | | | X | | | | | | | | | | X | | | | | | | | | | | | |
| **Clinical Assessment** | | | | | | | | | | | | | | | | | | | | | | | | | |
| Informed Consent and/or Eligibility | | X | X | | | | | | | | | | | | | | | | | | | | | | |
| Medical History with demographic data | | X | | | | | | | | | | | | | | | | | | | | | | | |
| Enrolment | | | X | | | | | | | | | | | | | | | | | | | | | | |
| Photo taken and Temporary ID Card | | X | | | | | | | | | | | | | | | | | | | | | | | |
| Permanent ID Card | | | X | | | | | | | | | | | | | | | | | | | | | | |
| Physical Examination | •Detailed | X | | | | | | | | | | | | | | | | | | | | | | | |
| | •Focused | | X | | X | | | | | X | X | X | X | | X | | | | | X | X | X | | X | X |
| Physical Assessments | •Vital Signs | X | X | | X | | | | | X | X | X | X | | X | | | | | X | X | X | X | X | X |
| | •Weight and Height | X | X# | | | | | | | | | | X# | | | | | | | | | | | | X# |
| | •MUAC | X | X | | | | | | | | | | X | | | | | | | | | | | | X |
| Adverse Events Data Collection | •Solicited | | X | X | X | X | X | X | X | X | | | X | X | X | X | X | X | X | X | | | | | |
| | •Unsolicited | | X | X | X | X | X | X | X | X | X | X | X | X | X | X | X | X | X | X | X | X | | | |
| | •SAE & New Medical Conditions | | X | X | X | X | X | X | X | X | X | X | X | X | X | X | X | X | X | X | X | X | X | X | X |
| Supply Insecticide Treated Nets (ITN) | | X | | | | | | | | | | | | | | | | | | | | | | | |
| **Laboratory Assessment:** Numbers indicate volume of blood to be collected at each timepoint | | | | | | | | | | | | | | | | | | | | | | | | | |
| Hematology | •RBC, HGB, platelets, WBC with differential (neutrophil, lymphocyte and eosinophil) counts | 0.5 | 0.5 | | | | | | | 0.5 | 0.5 | 0.5 | 0.5 | | | | | | | 0.5 | 0.5 | 0.5 | | | 0.5 |
| Biochemistry | •ALT,Creatinine | 1 | 1 | | | | | | | 1 | 1 | 1 | 1 | | | | | | | 1 | 1 | 1 | | | 1 |
| Parasitology | •Thick blood smear/Parasite Genotying | 0.5 | | | | | | | | | | | 0.5 | | | | | | | 0.5 | 0.5 | 0.5 | 0.5 | 0.5 | 0.5 |
| Serology [HIV, HBV and HCV] | | 1 | | | | | | | | | | | | | | | | | | | | | | | |
| Immunology | | | 10 | | | | | | | | 10 | 10 | 10 | | | | | | | 10 | | 10 | 10 | 10 | 10 |
| Urine Dipstick [Blood and Protein] | | X | | | | | | | | | | | | | | | | | | | | | | | |
| Total Blood Volume withdrawn/Visit (mL► | | 3.0 | 11.5 | 0.0 | 0.0 | 0.0 | 0.0 | 0.0 | 0.0 | 1.5 | 11.5 | 11.5 | 12.0 | 0.0 | 0.0 | 0.0 | 0.0 | 0.0 | 0.0 | 12.0 | 2.0 | 12.0 | 10.5 | 10.5 | 12.0 |
| Cumulative Blood Volume withdrawn (mL► | | 3.0 | 14.5 | 14.5 | 14.5 | 14.5 | 14.5 | 14.5 | 14.5 | 16.0 | 27.5 | 39.0 | 51.0 | 51.0 | 51.0 | 51.0 | 51.0 | 51.0 | 51.0 | 63.0 | 65.0 | 77.0 | 87.5 | 98.0 | 110.0 |

* Laboratory tests for this visit must be done within 24 hours before vaccination.

** This includes a screening visit 2 when participant will receive their results

### Only Weight will be measured

#### 22. APPENDIX B: SAE REPORTING FLOW CHART

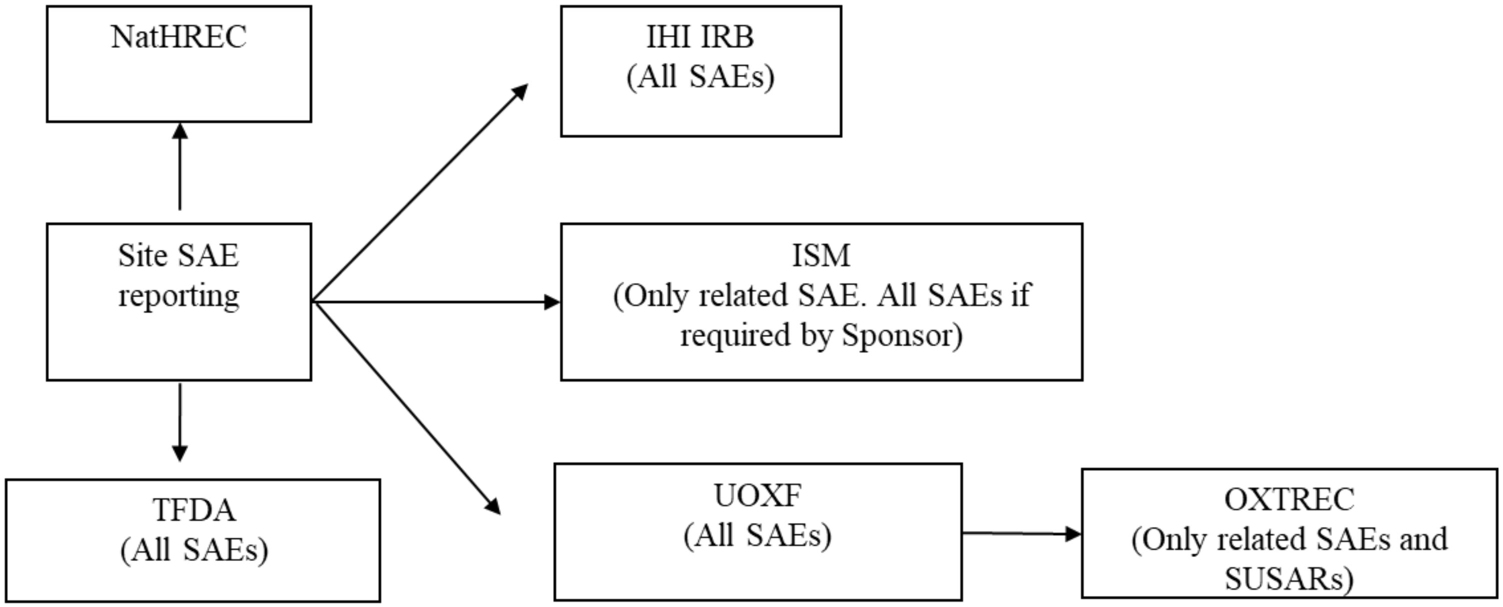

### 23. APPENDIX C: AMENDMENT HISTORY

| Amendment No. | Protocol Version No. | Date issued | Author(s) of changes | Details of Changes made |
|---|---|---|---|---|
| 1 | 1.1 | 19th October 2017 | Ally Olotu | • Addition of Sponsor and IHI logo in the first page of the protocol.<br>• Addition of the name of Paediatrician and a study physician in the investigator's team<br>• Retention of local safety monitor name in the protocol to be consistent with other protocols.<br>• Addition of the name of the project manager for the study.<br>• Section 13.5 updated: SMC consisting of 3 members will be established. All areas mentioning ISM have been updated for consistency.<br>• Section 11.7 has been revised to clarify under what situation the blood samples will be destroyed or stored indefinitely.<br>• The section 10.5 of the protocol under the subheading "Withdrawal of study participant" has been revised to provide more clarification and on what will happen to already collected data and samples.<br>• Section 9.5 describing the vaccination service in Tanzania has been revised to reflect the up to date information from the IVD program.<br>• Section 9.6 (Rationale for the trial design) has been updated to clarify the rationale for the choice of the vaccine dose.<br>• Sub-section 17.9 under ethical and regulatory considerations section titled "Future use of stored samples" has been created to give details of the future use of samples.<br>• Outline of study procedures tables have been updated to provide clarification for the number of screening visits, time points when only weight will be measured, and corrected topographical errors.<br>• Other topographical errors in the protocol have been corrected. |

| | | | | |
|---|---|---|---|---|
| 2 | 1.1 | 23rd October 2017 | Ally Olotu | • Clarification text in sections 11.7 and 17.9 with regard to future use of stored samples |
